# The genetic architecture of MRI derived human cervical spinal cord morphology reveals sensory-motor axis and biomarkers of neurological and systemic diseases

**DOI:** 10.1101/2025.08.05.25333008

**Authors:** Zhuopin Sun, Jiru Han, Zachary F Gerring, Victoria E. Jackson, Oneil G Bhalala, Ian H. Harding, Thiago J.R. Rezende, Melanie Bahlo

## Abstract

The spinal cord is critical to motor, sensory, autonomic function, and increasingly implicated in neurological disease and human health, yet its genetic architecture remains largely unexplored. We performed the first large-scale phenotyping of the upper cervical spinal cord (C1-C3) structure using brain magnetic resonance imaging from over 40,000 UK Biobank participants, extracting shape metrics including cross-sectional area, diameters, and eccentricity. We identified a total of 179 independent genome-wide significant variants, with cervical spinal cord morphology showing moderate to high SNP-based heritability (0.16 to 0.42). We also uncovered sex-specific genetic signals, highlighting potential biological sex differences in spinal cord development. In addition, spinal cord structure was associated with a wide range of neurological, metabolic, and systemic conditions, such as multiple sclerosis, neuropathies, diabetes, and attention-deficit/hyperactivity disorder. These findings establish the cervical spinal cord as a genetically informative and health relevant structure, offering new opportunities to study its role in disease mechanisms and human health.

## INTRODUCTION

The spinal cord is a key component of the human nervous system, serving as the primary communication pathway between the brain and the rest of the body outside of the head. Its complex structure and functions enable the efficient passage of ascending sensory signals, descending motor commands, and sensory-motor integration, allowing for smooth movement coordination, environmental feedback, and autonomic function^1^. Through sensory and motor tracts, the spinal cord connects to the brainstem, thalamus, basal ganglia, cerebellum, and motor-sensory cortices^2^. Structural and functional abnormalities of the spinal cord are implicated in a wide range of health conditions, including multiple sclerosis (MS)^3,4^, motor neuron diseases^5,6^, spinal cord injury^7,8^, pain^9,10^, ataxias,^11,12^ and other degenerative conditions. Studying the spinal cord is essential not only for defining neural organization and sensorimotor integration, but also for understanding pathological changes that inform clinical diagnosis, symptom manifestation, and treatment.

In recent years, growing clinical and population cohorts linking imaging-derived phenotypes (IDPs) and genetic variation have offered a powerful opportunity to examine the biological mechanisms underlying development and disease susceptibility. Previous genome-wide association studies (GWAS) using the UK Biobank (UKB) imaging and genetic data have explored a wide range of IDPs, including structural and functional brain measurements^13–18^, retinal thickness^19^, skeletal proportions,^20^ and other organ-specific traits^21^. However, while GWAS have identified hundreds of loci associated with magnetic resonance imaging (MRI) derived brain phenotypes, the genetics of the spinal cord remain largely unexplored. This gap reflects a major limitation in our understanding of the genetic basis of the full central nervous system and its relationship to health and disease.

Recent advances in neuroimaging have enabled non-invasive characterization of spinal cord anatomy and functions^22,23^. These imaging-based measures have uncovered structural and functional changes associated with neurological diseases^24,25^, injury^26,27^, and biological factors^28,29^. Genetically, a small cohort twin study^30^ suggested high heritability of cervical spinal cord structure (h^2^=0.85-0.91), and genotype-specific spinal cord damage has been reported in spinocerebellar ataxias^31^. These studies demonstrate the potential role of genetic variability in shaping spinal cord structure. However, existing findings have relied on modestly sized clinical cohorts. A key challenge has been the lack of reliable high-throughput methods to process spinal cord MRI at scale. This challenge is driven by the sensitivity of spinal cord imaging to anatomical variability, neck curvature, partial volume effects, motion, and distortion artefacts, making fully automated segmentation, labelling, and quality control difficult, especially in large scale datasets^22,32^.

To address this gap, we implemented a fully automated deep learning-based pipeline optimized for upper cervical spinal cord segmentation and labelling, which is robust to pathological variations and image quality heterogeneity. We applied this pipeline to the UKB T1-weighted brain MRI, extracting spinal cord shape metrics from over 40,000 participants. These IDPs capture the mean cross-sectional area (CSA), eccentricity, anterior–posterior (AP) and right-left (RL) diameters of the spinal cord at the C1, C3, and C3 vertebral levels. Leveraging these quantitative traits, we conducted the first GWAS of spinal cord morphology, establishing its genetic architecture and heritability, and highlighting potential links to development and disease susceptibility, identifying new and further characterizing known biomarkers. This work establishes a scalable framework for spinal cord imaging genetics and highlights the value of incorporating spinal cord into future integrative studies of human health and disease.

## RESULTS

### Spinal cord phenotypes

We applied and validated a fully automated pipeline optimized for upper cervical spinal cord segmentation and phenotyping^33^. The pipeline is illustrated in Fig. 1a and is detailed in the Methods section. In brief, raw T1-weighted brain MRI data underwent spinal cord segmentation using the Spinal Cord Toolbox^23^ and then vertebral labelling of the first three cervical spinal cord segments (C1–C3) using a pretrained nnU-Net model^34^. Twelve (4 measurements x 3 spinal levels) spinal cord IDPs were then extracted, including CSA, eccentricity, AP and RL diameters of the spinal cord at each vertebral level. From an initial cohort size of n=46,203, data was available after imaging quality control (QC) for n=45,684 participants for C1, n=44,737 for C2, and n=40,024 for C3. The distribution of spinal cord phenotypes and their average values across age are shown in Fig. S1-2.

**Fig. 1:**
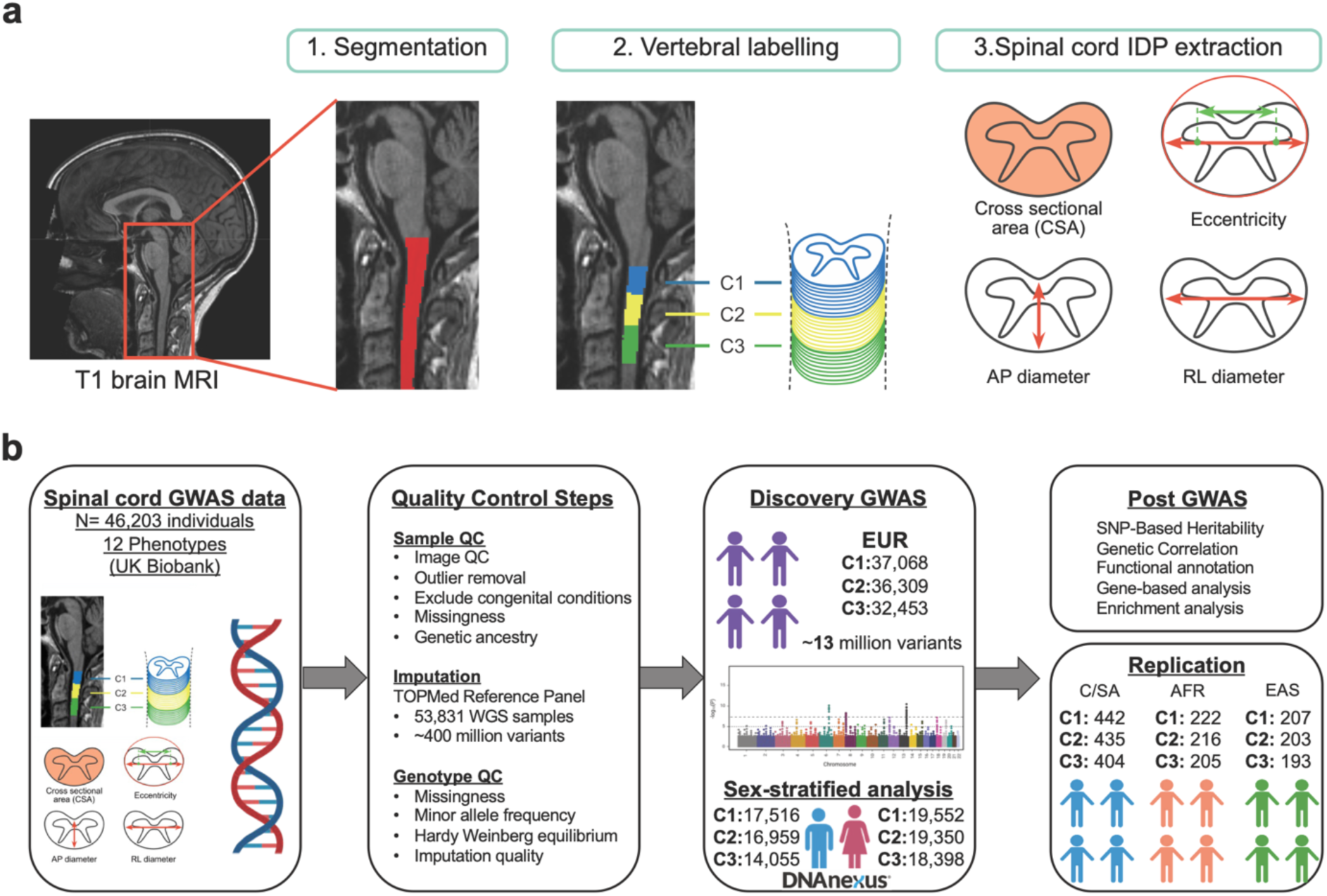
Genome-wide associations of cervical spinal cord shape metrics. **(a)** An overview of the spinal cord image processing pipeline using T1-weighted brain MRI from the UKB. Following spinal cord segmentation and vertebral labelling of C1 to C3, spinal cord shape metrics including the mean CSA, eccentricity, AP and RL diameters were derived. **(b)** Overview of the GWAS study design for spinal cord shape metrics. We conducted GWAS of 12 MRI-derived spinal cord shape phenotypes, including CSA, eccentricity, AP and RL diameters at cervical levels C1 to C3. Quality control included sample and genotype filtering, with imputation using the TOPMed reference panel. Discovery analyses were performed in individuals of European (EUR) ancestry, including sex-stratified GWAS, followed by post-GWAS analyses. Replication was carried out in C/SA, EAS, and AFR cohorts. Reproduced by kind permission of UK Biobank ©.

Follow-up data from repeat scans acquired 1-7 years (average 2.7 years) post-baseline were available for n=4374, 4270, and 3290 participants at C1, C2, and C3 respectively. Repeatability analysis indicated that eccentricity was highly stable across all levels (Spearman’s *r^2^* = 0.77-0.91) (Fig. S3), whereas CSA and diameters were more susceptible to age-related degeneration (Fig. S2). C1 phenotypes had moderate-to-high correlation (*r^2^* = 0.68-0.91) within the first four years, but had declining repeatability over longer time spans and weaker longitudinal agreement at C2 (*r^2^* = 0.62-0.89) and C3 (*r^2^* = 0.50-0.88) levels (Fig. S3). For robustness testing, spinal cord phenotypes at C2 and C3 showed high cross-method consistency across all traits (*r^2^* = 0.97-0.99) when compared to an alternative MRI atlas-based processing pipeline (Fig. S4). Detailed quality control and validation are presented in the Methods section.

### GWAS sample characteristics

Following sample and variant QC of the imputed genetic data (Fig. 1b and Methods), the discovery GWAS (mean age = 64 years, SD = 7.69 years, 52.6% female) focused on individuals of European ancestry, defined by the Pan-UK Biobank^35^. As detailed in the Methods, individuals with poor-quality imaging, those who failed standard QC, or those with congenital conditions and operative procedures affecting the cervical spine were excluded (Table S1). The study population included 37,068 individuals at the C1 spinal level, 36,309 at C2, and 32,453 at C3. In addition to sex-combined analysis, sex-stratified GWAS was also performed separately at each spinal level. For replication, we examined the independent significant SNPs identified in the European-ancestry GWAS among non-European individuals in the UKB, including those of Central and South Asian (C/SA), African (AFR), and East Asian (EAS) ancestry. Sample sizes for each group are shown in Fig. 1b.

### GWAS identifies 179 independent genome-wide significant variants

We conducted quantitative trait GWAS for all autosomes and the X chromosome for the 12 spinal cord IDPs using REGENIE^36^, adjusting for sex, age, age², genotyping batch, imaging site, intracranial volume (ICV), height, and the inverted signal-to-noise ratio in T1-weighted images. Across these traits, we identified 179 independent genome-wide significant variants using a significance threshold of *P* < 4.2 × 10⁻⁹, adjusted for testing across 12 phenotypes (Table S2). These variants were mapped to 104 unique genomic loci, including one on the X chromosome (Fig. 2). Manhattan and quantile–quantile (Q–Q) plots for each phenotype are presented in Fig. S5-8. The genetic inflation factor (*λ* = 1.04-1.09) and the LDSC intercept values (0.99-1.03) as shown in Table S3 suggest little to no inflation due to population stratification, or confounding factors. Eccentricity had the greatest number of associations, with 58 variants at C1, 46 at C2, and 35 at C3 (78 total, 13 shared across C1-C3). Other phenotypes showed fewer associations: AP diameters were associated with 22, 17, and 6 variants at C1, C2, and C3, respectively (27 total, 4 shared); RL diameters with 22, 11, and 7 variants (27 total, 5 shared); and CSA with 18, 13, and 6 variants (20 total, 6 shared).

**Fig. 2:**
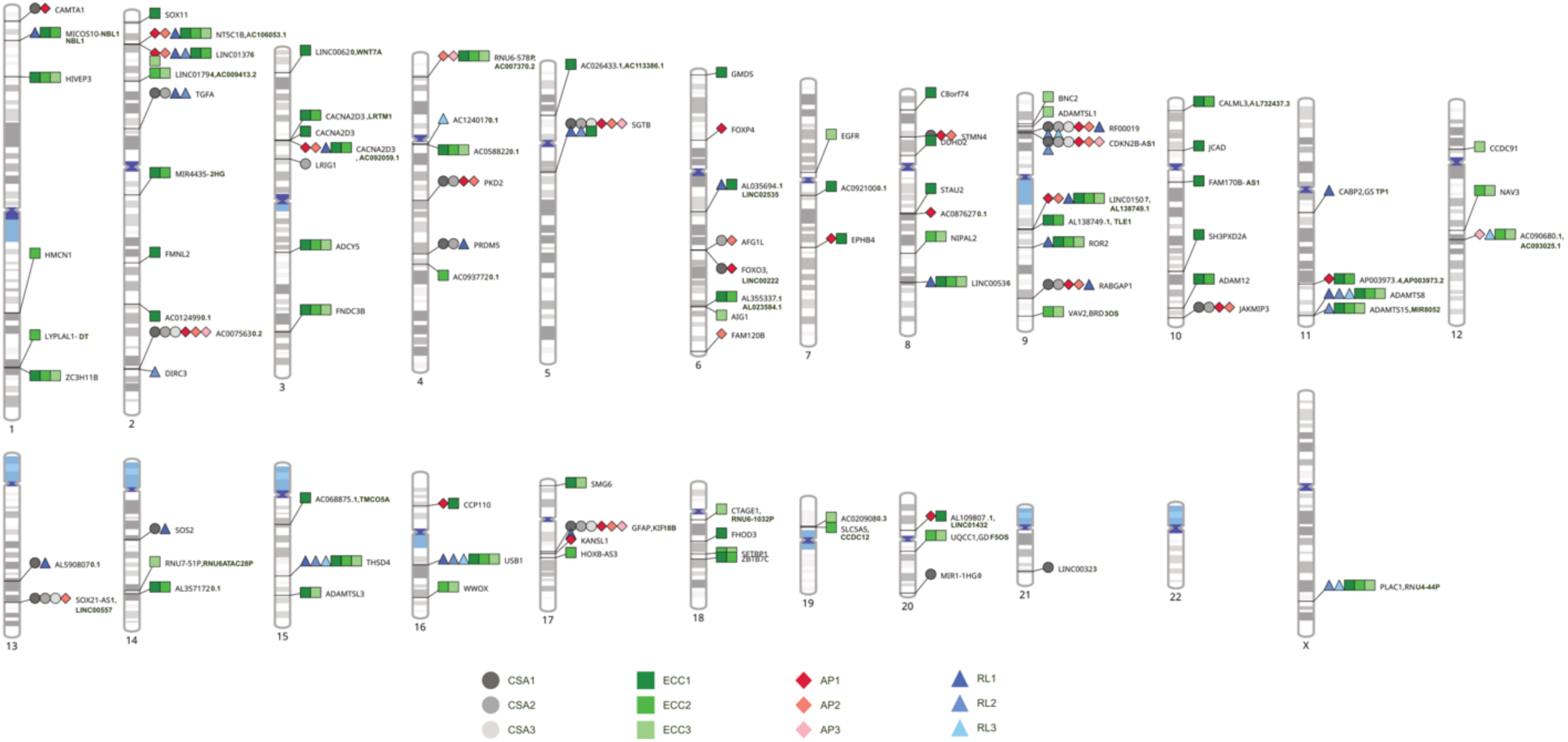
Ideogram of genomic loci associated with 12 spinal cord shape phenotypes. Shapes represent the four spinal cord shape metrics: CSA, eccentricity (ECC), anterior-posterior (AP) and right-left (RL) diameters, and are shaded by vertebral level (C1 to C3). Loci were defined by merging genome-wide independent significant variants (*P* < 4.17 × 10⁻⁹) that were in linkage disequilibrium (*r²* > 0.1) and located within 250 kb. Each locus is labelled with the nearest protein-coding gene, and loci annotated to the same gene were merged for clarity.

Among the 104 genomic loci associated with spinal cord IDPs, several were shared across multiple phenotypes and/or across spinal levels (Fig. S9). Notably, a locus on chromosome 5, spanning a gene cluster that includes *ADAMTS6*, *CENPK*, *TRAPPC13*, and *SGTB*, was associated with all four spinal cord shape metrics (CSA, eccentricity, AP and RL diameters). The most statistically significant association at this locus was observed for rs201934842, located in the 3′ UTR of *SGTB*, with C1 CSA (Minor allele frequency (MAF) =0.26, *beta* = 0.079, *P* = 2.65 × 10⁻²³). *SGTB* has previously been linked to brainstem volume and is implicated in synaptic protein homeostasis^37^. A few rare (MAF < 0.01) SNPs exhibited relatively large effect sizes (Fig. S10). For example, rs554265256 (MAF = 0.0083), located in an intronic region of *RABGAP1*, was associated with CSA, AP, and RL diameters (peak association with CSA C1: *beta* = -0.29, *P* = 6.5 × 10⁻¹⁴). The rs554265256 variant has been previously linked to iron levels in the brain according to the GWAS Catalog^38^. RABGAP1 is a GTPase-activating protein involved in a variety of key cellular and molecular processes, including vesicular trafficking and cell migration. Dysfunction of RABGAP1 is linked to neurodevelopmental syndrome^39^ and potentially neurodegenerative disease^40^.

We further cross-referenced these 179 independent genome-wide significant variants with the GWAS Catalog^41^ and found that 50 variants have been previously associated with a range of traits, spanning across 120 unique phenotypes (Table S4). The most frequently reported associations involved brain-related traits (e.g., cerebral cortex area, white matter, and cognitive performance), blood pressure measures (systolic, diastolic, and pulse pressure), and body composition (e.g., body mass index [BMI], waist-hip ratio). These findings suggest enrichment in neurological, cardiovascular, and anthropometric phenotypes.

We considered height as an important covariate in the main GWAS results as it was shown to have a significant correlation on brain volume and spinal cord^28^, and we confirmed this in the stepwise covariate selection analysis (Fig. S11). For additional sensitivity analysis, we performed GWAS for the 12 phenotypes without adjusting for height (Fig. S12–15, Table S3). SNP effect sizes showed high concordance in both direction and magnitude, with Pearson correlation coefficients ranging from 0.986 to 0.999 when comparing results with and without height adjustment (Table S5). P-values were also highly correlated (*r* = 0.944–0.996).

### Heritability and genetic correlation of spinal cord shape metrics

We estimated SNP-based heritability for the four spinal cord shape metrics across the C1, C2, and C3 spinal levels using linkage disequilibrium score regression (LDSC) applied to the GWAS summary statistics^42^. Heritability estimates ranged from 0.16 to 0.42, indicating moderate to strong genetic contribution (Fig. 3a, Table S6). Eccentricity showed the highest heritability overall, with estimate of 0.42 at C1, 0.36 at C2, and 0.31 at C3. We observed a consistent trend of decreasing heritability from the C1 to C3 levels across all four traits.

**Fig. 3:**
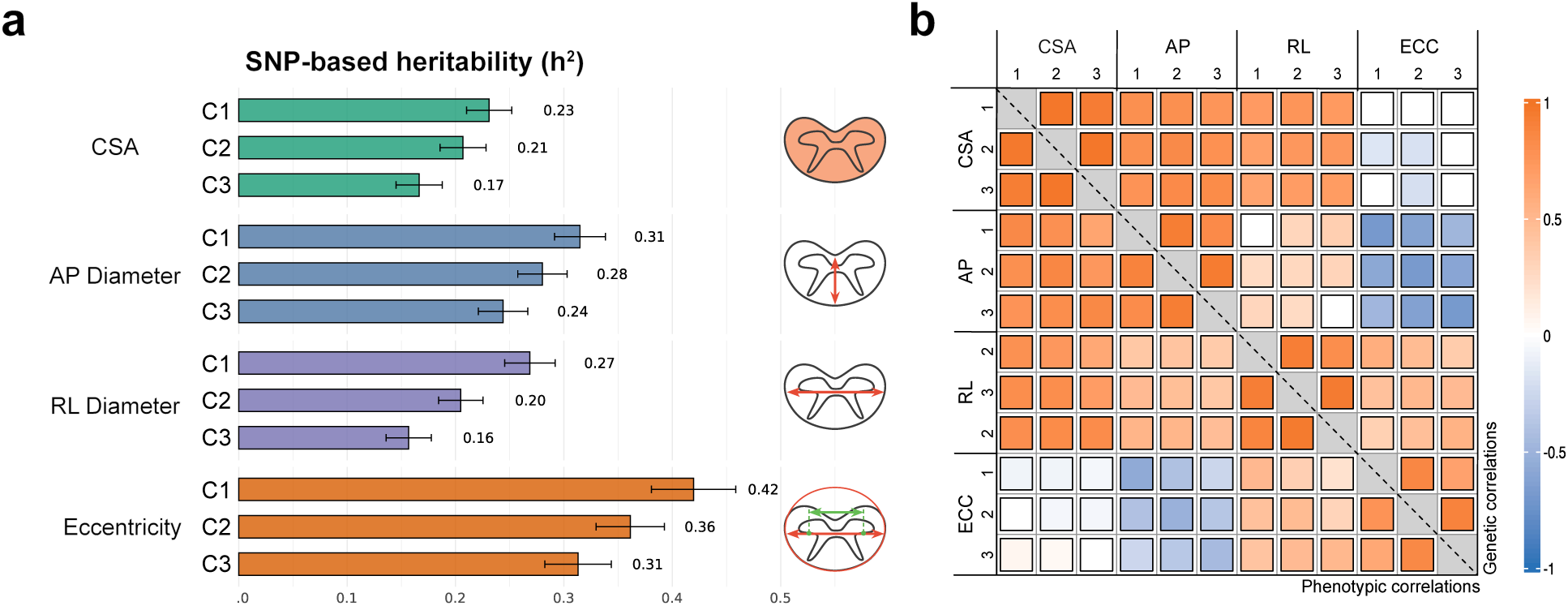
Heritability estimation and correlations of spinal cord shape metrics. **(a)** SNP-based heritability (*h^2^*) and **(b)** phenotypic (lower half of the heatmap) and genetic correlations (upper half of the heatmap) of the spinal cord shape metrics. AP: Anterior-Posterior; RL: Right-Left; CSA: cross sectional area; ECC: eccentricity.

We next examined genetic correlations among the 12 spinal cord phenotypes using LDSC to assess the degree of shared genetic architecture (Fig. 3b, Table S7). CSA showed strong genetic correlations across all spinal levels, with the highest correlation between C1 and C2 (*r*_g_ = 0.98), followed by C2–C3 (*r*_g_ = 0.98) and C1–C3 (*r*_g_ = 0.94), indicating consistent genetic influence across the upper cervical spine CSA. AP and RL diameters also exhibited strong inter-level correlations ranging from 0.93 to 0.96. In contrast, eccentricity showed moderate within-phenotype correlations (*r*_g_ = 0.87 between C1 and C2; *r*_g_ = 0.67 between C1 and C3), suggesting partially distinct genetic determinants across levels.

Between phenotypes, CSA was strongly correlated with both AP and RL diameters (*r*_g_ = 0.72-0.80), as expected given their geometric relationship. Eccentricity, derived from the ratio of RL and AP diameters, was negatively correlated with AP diameter (peak *r*_g_ = –0.69) and positively correlated with RL diameter (peak *r*_g_ = 0.58). In contrast, CSA and eccentricity were only weakly correlated, suggesting that they represent distinct traits and likely capture distinct biological dimensions. The overall pattern of spinal cord shape metrics phenotypic correlations closely mirrored the corresponding genetic correlations (Fig. 3b, Table S8).

### Sex-specific genetic associations

We then investigated potential sex differences in the genetic architecture of spinal cord shape by conducting sex-specific GWAS for each of the 12 traits. Miami plots of the sex-stratified GWAS results are shown in Fig. S16-19. Genomic inflation factors ranged from 1.02 to 1.06 in females and 1.00 to 1.05 in males, with LDSC intercepts ranging from 0.99 to 1.01 in both sexes (Table S3). SNP-based heritability estimates ranged from 0.17 to 0.38 in females and 0.15 to 0.43 in males, following a similar pattern to that observed in the sex-combined GWAS (Table S6). We further assessed the concordance between sexes and observed a strong correlation in SNP-based heritability estimates across the 12 traits (Pearson correlation coefficients *r* = 0.887, P = 0.00012; Fig. S20).

We identified two independent genetic signals (*P* < 4.17 × 10⁻⁹, Bonferroni-corrected for 12 traits) that were not detected through our sex-combined analyses. Rs150219059, an intronic SNP in *PEPD,* was significantly associated with AP diameter C2 spinal level in males (MAF = 0.045, *beta* = 0.15, *P* = 1.94 × 10⁻⁹), but not in females (MAF = 0.047, *beta* = 0.015, *P* = 0.52). Similarly, the intergenic SNP rs373400435 was associated with AP diameter C1 spinal level in males only (males: MAF = 0.0028, *beta* = 0.6, *P* = 1.22 × 10⁻⁹, females: MAF = 0.0029, *beta* = 0.057, *P* = 0.54). SNP by sex interaction analysis confirmed a significant sex-interaction for these two SNPs (Table S9, Fig. S21).

SNP-by-sex interactions were additionally examined for all independent SNPs identified through the sex-combined discovery analyses. One SNP, rs2328459, associated with C3 eccentricity, was found to have a significant interaction (*P* = 9.65 × 10⁻^6^, Benjamini-Hochberg *P* = 0.0026, Table S9). The sex-specific analyses for this SNP revealed a significant association in females (MAF = 0.54, *beta* = -0.069, *P* = 1.1 × 10⁻^11^, Fig. S21), but not males (MAF= 0.54, *beta* = -0.012, *P* = 0.31). This SNP is located within an intron of *AIG1* (Androgen Induced 1), which plays a role in fatty acid metabolism and may be regulated by androgen levels.

### Association in the non-European ancestries

We performed replication analyses of the 179 independent genome-wide significant loci identified in the European-ancestry discovery GWAS of 12 spinal cord shape metrics using independent data from three non-European cohorts from the UK Biobank: C/SA (*n* = 442-404), AFR (*n* = 222-205), and EAS (*n* = 207-193) participants across the C1 to C3 spinal levels. Of these, 178 variants were available in the C/SA cohort, 173 in the EAS cohort, and 176 in the AFR cohort after excluding SNPs with a MAF < 0.05 within each respective population cohort. No variants reached Bonferroni-corrected significance in any non-European group (*P* < 0.00028, correcting for 179 tests). Effect size estimates from the European cohort were compared with those observed in each non-European ancestry group across the 12 spinal cord shape metrics (Fig. S22-24). Directional concordance ranged from 28.6% (RL diameter C3 level in the AFR cohort) to 83.3% (AP diameter C3 level in both the EAS and C/SA), with overall higher concordance observed in C/SA and EAS compared to AFR. Spearman correlation coefficients between effect sizes in European and non-European populations were generally positive, although most were not statistically significant. The highest correlation was observed for AP diameter C3 level in the C/SA cohort (*r* = 0.94, *P* = 0.017), while the lowest was for RL diameter C3 level in the AFR cohort (*r* = –0.61, *P* = 0.17) (Fig. S22-24). These results suggest that many loci exhibit consistent effect directions across ancestries, particularly in the C/SA and EAS cohorts. However, the lack of statistical significance and the variability in correlation strength reflect the limited power of the current non-European samples.

### Gene-based findings

Candidate risk genes for each of the four morphological features of cervical spinal cord segments C1–C3 were prioritized using six gene-based mapping approaches. Positional mapping was conducted with MAGMA and mBAT-combo, while functional mapping leveraged eQTL data via TWAS (PsychENCODE and GTEx v8 expression weights) and SMR (eQTLGen and MetaBrain datasets). Across all spinal cord morphology-segment combinations, we identified 432 unique significant genes after applying a Bonferroni correction (threshold of *P* = 0.05/N genes tested for a given gene-based method). The largest number of unique gene associations was found for eccentricity (*n* = 237), followed by AP diameter (*n* = 188), CSA (*n* = 162), and RL diameter (*n* = 113). The proximity-based approaches mBAT-combo and MAGMA identified the largest number of significant genes, with 292 and 195 genes, respectively, followed by TWAS of GTEx tissues with 127 significant gene associations (Table S10-13). Of the 432 genes significantly associated with a spinal cord phenotype in at least one approach, 218 were implicated by at least two approaches and 96 by at least 3 approaches (Table S14-15).

Among the 96 genes identified by at least three mapping approaches, gene candidates for spinal cord phenotypes were further prioritized using colocalization (TWAS-COLOC) and SMR-HEIDI analyses (Fig. 4a; Table S10-13, Fig. S25). Colocalization identified TWAS associations supported by a shared causal variant between GWAS and eQTL signals (posterior probability H4 > 0.8). HEIDI was used to retain SMR associations consistent with a single causal variant influencing both gene expression and trait (P > 0.05). Of the 96 genes implicated by at least three gene-based tests, 44 had a significant TWAS association in GTEx brain tissue and showed evidence of colocalization (i.e., PP.H4 > 0.8), while 41 genes had a significant SMR association in brain tissue, 29 of which had a HEIDI *P* value > 0.05. Top among the list of high confidence genes was *SGTB,* which was implicated by five gene-based approaches across three morphological measures and showed evidence of colocalization in human brain tissue. Other high confidence genes included *TGFA* and *ROR2*, both of which were implicated by five gene-based approaches (across three spinal levels) and colocalized in human brain tissue (Fig. 4a).

**Fig. 4.**
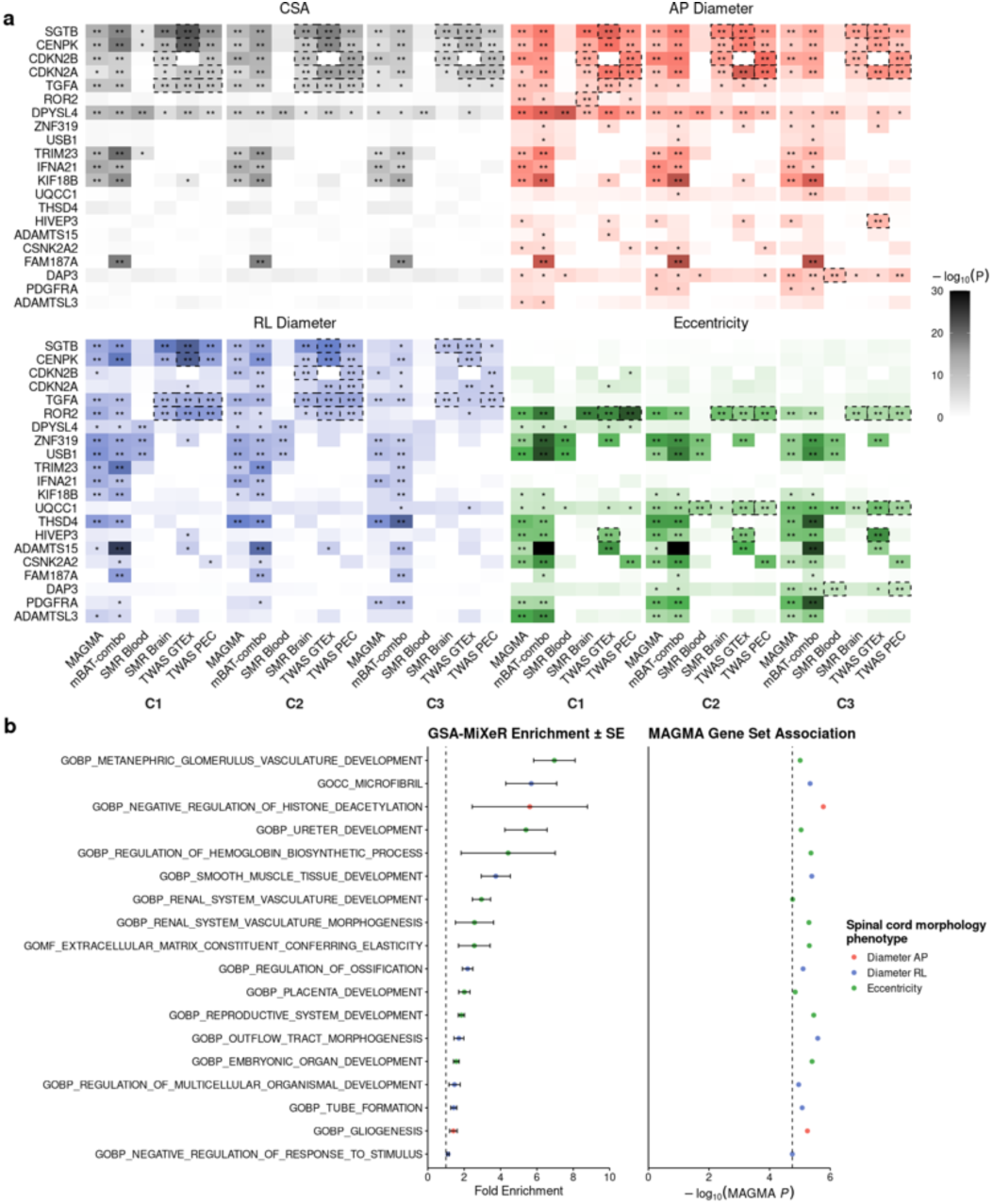
Top 20 prioritised gene-based associations for spinal cord morphology phenotypes. (**a)** For each combination of spinal cord vertebra (C1, C2, C3) and spinal cord morphology phenotype (CSA, AP diameter, RL diameter, and eccentricity), we extracted gene-level association results from six gene-based analysis methods: MAGMA, mBAT, SMR (BrainMeta and eQTLgen), and TWAS (GTEx and PsychENCODE). For each method and verbetra-morphology feature pair, we identified the gene with the minimum p-value. From these results, a set of unique gene identifiers was extracted to summarize the most significant associations across all analytic approaches and phenotype combinations. The genes are ordered by the number of times they were significant after Bonferroni correction across all verbetra-morphology feature pair combinations. * FDR < 0.05; ** Bonferroni P < 0.05; dashed boxes represent significant gene-based associations (Bonferroni P < 0.05) with evidence the same causal variant underlies the GWAS and eQTL association using the HEIDI test in SMR (HEIDI P > 0.05) or colocalisation in TWAS (COLOC PP.H4 > 0.8). **(b).** Gene sets prioritized by MAGMA, ranked by GSA-MiXeR fold enrichment. GSA-MiXeR fold enrichment estimates with standard errors; the vertical dashed line denotes enrichment threshold (fold enrichment = 1). MAGMA -log(10) P values gene set analysis; the vertical dashed line indicates the Bonferroni-corrected significance threshold. Abbreviations: GOMF, Gene Ontology Molecular Function; GOBP, Gene Ontology Biological Process; GOCC, Gene Ontology Cellular Component.

### Gene Set Enrichment Analysis

We applied GSA-MiXeR and MAGMA gene-set enrichment analysis to each spinal cord morphology phenotype (Fig. 4b, Table S16). Among the 20 significant (*P_FDR_* < 0.05) MAGMA gene sets, gene sets with the largest GSA-MiXeR enrichment included pathways related to organ development (e.g., metanephric glomerulus vasculature [GSA-MiXeR enrichment = 6.96; MAGMA *P* = 9.82 × 10⁻^6^] and smooth muscle tissue development [enrichment = 3.74; MAGMA *P* = 4.06 × 10⁻^6^]). We also identified more specific biological pathways related to spinal cord development and morphology, including microfibril pathways (enrichment = 5.69; MAGMA *P* = 4.61 × 10⁻^6^), tube formation (enrichment = 1.43; MAGMA *P* = 8.43 × 10⁻^6^), and gliogenesis (enrichment = 1.41; MAGMA *P* = 5.65 × 10⁻^6^).

### Associations with brain-image derived phenotypes

We examined genetic and phenotypic correlations between spinal cord phenotypes and brain volumes (Fig. 5, Table S17-18). Both genetic and phenotypic correlations revealed that spinal cord CSA and diameters were most strongly correlated with the brainstem, with the medulla showing the highest genetic correlations with C1 spinal levels (*r*_g_ = 0.63, 0.47, and 0.52 for CSA, AP diameter, and RL diameter, respectively), followed by the pons and midbrain with similar trends. C2 and C3 spinal levels showed slightly lower but comparable correlations to C1.

**Fig. 5.**
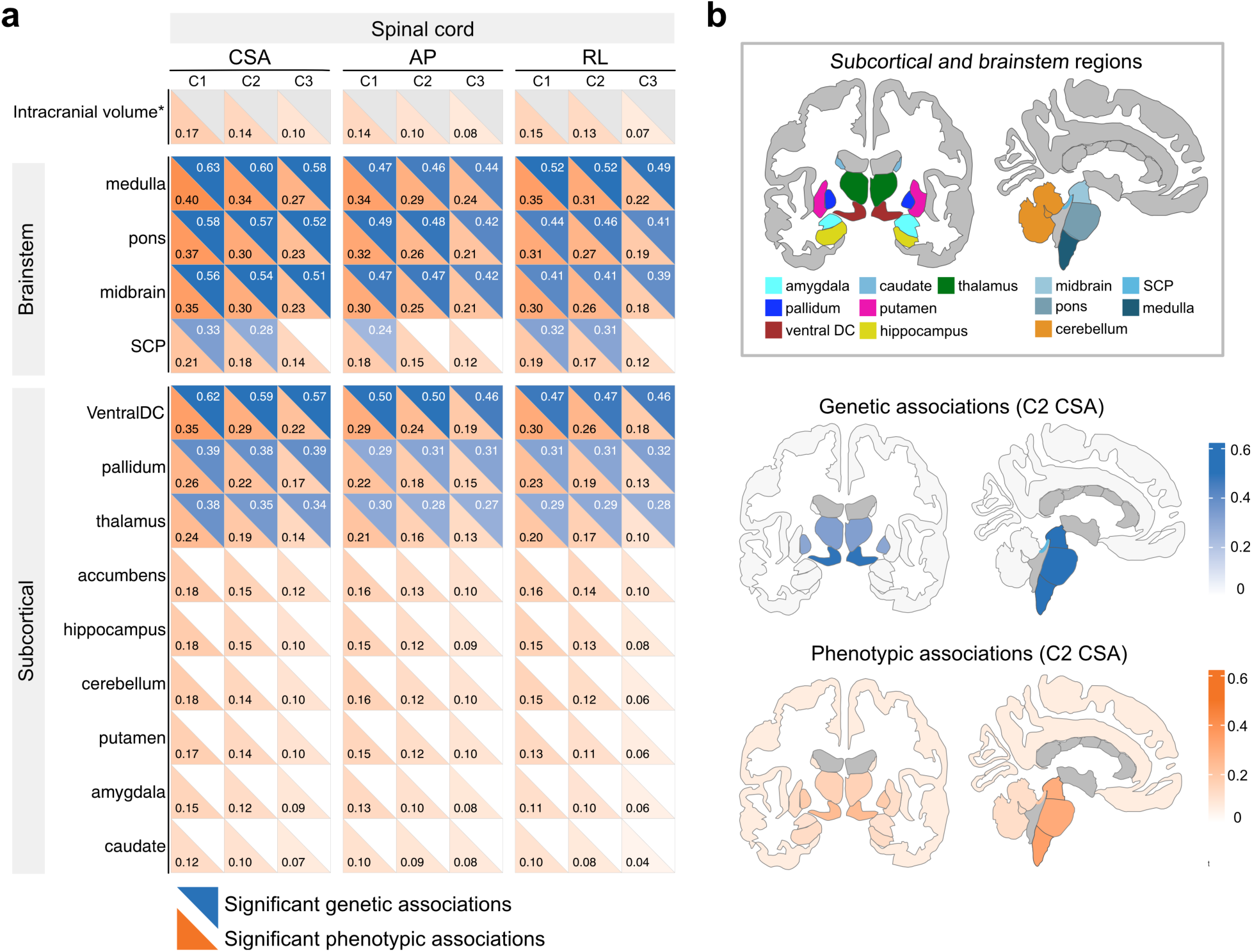
Genetic and phenotypic associations between spinal cord phenotypes and brain volumes. (**a)** Heatmap of significant associations (Bonferroni-corrected *P* < 0.05/504) sorted by effect size: upper blue triangle shows genetic correlations, lower orange triangle shows phenotypic associations. Eccentricity and cortical regions are not displayed due to no significant genetic associations. Genetic correlation for ICV is not calculated as it’s used as a covariate in GWAS. (**b)** top row: subcortical and brainstem regions in Freesurfer aseg atlas; middle: significant genetic associations for C2 CSA; bottom: phenotypic associations for C2 CSA. SCP: Superior Cerebellar Peduncle; ventral DC: ventral diencephalon. Colour intensity indicates magnitude of the beta values.

Among subcortical structures, the ventral diencephalon (*r*_g_ = 0.62, 0.50, 0.47 for CSA, AP diameter, and RL diameter), pallidum (*r*_g_ = 0.39, 0.29, 0.31), and thalamus (*r*_g_ = 0.38, 0.30, 0.29) exhibited the strongest genetic correlations with CSA and diameters at C1 spinal level. Several additional subcortical regions showed weaker significant genetic correlations with spinal cord phenotypes after false discovery rate (FDR) correction, including the cerebellum, hippocampus, accumbens, amygdala, and putamen (Fig. S26 and Table S17), but did not survive multiple testing correction. In cortical regions, the strongest phenotypic associations were observed in motor areas, particularly the bilateral anterior and posterior Brodmann area (BA) 4 (motor) regions (Fig. S26 and Table S18). Genetically, the right posterior BA4 showed the highest correlation but did not survive multiple testing correction. As expected, eccentricity, being a shape metric, did not show strong correlations with other brain volumes.

### Links to Disease and Health-Related Traits

We tested associations between spinal cord shape metrics and PhecodeX diseases from UKB health records, identifying significant phenotypic associations across 14 major disease categories (Fig. 6a, Table S19). Neurological diseases showed the greatest number of significant associations. Smaller spinal cord sizes, measured by reduced CSA and AP and RL diameters, were consistently associated with increased risk of neurological conditions. Strongest negative associations were observed in MS (peak *r*_p_ = -0.55 for C1 AP diameter), encephalitis, and both polyneuropathies and mononeuropathies. Weaker, but notable associations were also observed with nerve root disorders and sleep disorders. In some conditions, such as polyneuropathies, increased eccentricity, reflecting flatter spinal cord, was also associated with higher disease risk. Beyond neurological disorders, we observed strong and consistent negative associations between spinal cord size and metabolic conditions, including type 1 (highest *r*_p_ = -0.4 for C1 AP diameter) and type 2 diabetes (highest *r*_p_ = -0.28 for C1 AP diameter). Additional associations were identified in categories such as viral infections and musculoskeletal conditions. These findings suggest that spinal cord morphology may reflect broader systemic health and disease susceptibility.

**Fig. 6.**
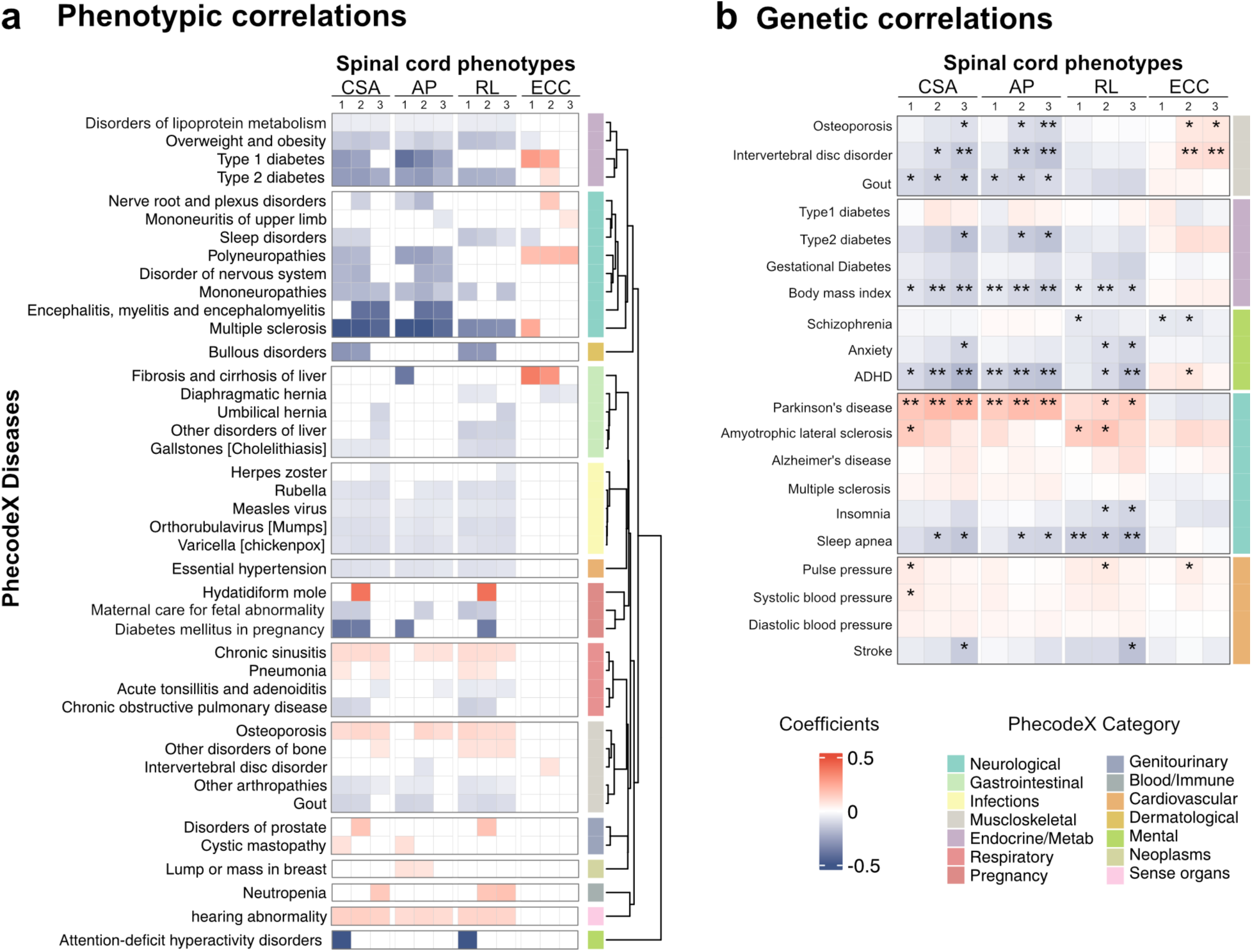
Associations between spinal cord phenotypes and health-related outcomes. **(a)** Heatmap of phenotypic associations between spinal cord shape metrics and health-related outcomes (converted to Phecode diseases) in the UKB. Only associations with FDR-adjusted *P* < 0.05 and diseases associated with multiple spinal cord phenotype are displayed. Diseases are grouped by category (legend on the right) and ordered by hierarchical clustering based on similarity in association patterns across 12 phenotypes. **(b)** Genetic associations for diseases/ traits. Asterisks indicate significance: * *P* < 0.05; **FDR-adjusted *P* < 0.05. Blue indicates negative effect size; red indicates positive effect size. CSA: cross-sectional area; AP: anterior–posterior diameter; RL: right–left diameter; ECC: eccentricity.

We replicated the analysis in the non-European ancestry UKB samples (Table S20). Due to limited sample size, many diseases lacked sufficient cases in the UKB (i.e. only 1 MS case among non-European samples). For conditions with adequate representation, we observed consistent effect directions and comparable effect sizes including for type 2 diabetes, polyneuropathies, mononeuropathies, and viral infections (*P* < 0.05, but did not survive FDR correction).

Following the exploratory phenotypic associations with health-related outcomes, we used bivariate LDSC to estimate genetic correlations between spinal cord shape metrics and 20 disease traits from well-powered GWAS studies spanning neurological, musculoskeletal, metabolic, cardiovascular, and mental health conditions (Fig. 6b, Table S21). Several traits showed significant genetic correlations with at least one spinal cord phenotype, after FDR correction (*P* < 0.05): Intervertebral disc disorder; osteoporosis; BMI; Attention-deficit/hyperactivity disorder (ADHD); Parkinson’s disease (PD); and sleep apnea. The genetic correlations with greatest magnitude were with PD and ADHD. PD showed positive genetic correlation with spinal cord sizes, measured by CSA (C2: *r*_g_ = 0.20, *P_FDR_* = 6.55 × 10⁻³), diameter AP (C2: *r*_g_ = 0.18, *P_FDR_* = 6.55 × 10⁻³). We also found ADHD had significant negative genetic correlations with spinal cord, measured by CSA (C3: *r*_g_ = –0.2, *P_FDR_* = 6 × 10⁻³), diameter AP (C3: *r*_g_ = –0.17, *P_FDR_* = 3.27 × 10⁻³), and RL metrics (C3: *r*_g_ = –0.16, *P_FDR_* = 0.03). These results suggest potential shared genetic architecture between spinal cord shape metrics and a range of disease categories.

For traits showing significant genetic correlations with spinal cord phenotypes, and with sufficient genetic instruments (≥3 variants; BMI, PD, ADHD, sleep apnea and osteoporosis), we next utilized Mendelian randomization (MR) methods to explore potential causal effects of these traits on spinal cord shape metrics. Our primary MR analyses utilized the two-stage, inverse-variance weighted (IVW) approach^43^, and identified potential causal effects of PD on CSA (peak association at C3: *beta* = 0.037, *P_FDR_* = 0.017) and AP diameter (C3: *beta* = 0.043, *P_FDR_* = 0.017), and of BMI on CSA (C2: *beta* = -0.12, *P_FDR_* = 0.001), AP diameter (C2: *beta* = -0.15, *P_FDR_* = 8.11 × 10⁻^5^), and RL diameter (C3: *beta* = -0.09, *P_FDR_* = 0.012) (Table S22, Fig. S27).

To evaluate the robustness of these results, we performed additional analyses using a series of complementary MR approaches (Table S23, Fig. S27). For BMI, similar causal estimates were obtained using weighted median^44^ and weighted mode^45^ methods; however no evidence for a causal effect was identified, through the MR-Egger^46^ and CAUSE^47^ approaches. We observed significant heterogeneity in the effects of the genetic instruments (Cochran’s Q statistic *P*-values all ≤ 3.45 × 10^-10^). Furthermore, reverse MR revealed an apparent causal effect of CSA and AP diameter on BMI, with a direction of effect opposite to that observed in the forward MR (Table S24). Together, these results suggest potential violations of MR assumptions, such as horizontal pleiotropy. For PD, no alternate approach (Weighted Median, Weighted Mode, MR-Egger, CAUSE) provided evidence of a causal effect on spinal cord phenotypes. Overall, the inconsistencies across methods suggest the original IVW results may not be valid due to violations of MR assumptions. Thus, our results do not provide robust evidence of a causal effect of PD or BMI on spinal cord phenotypes.

## DISCUSSION

This study presents the first GWAS of spinal cord morphology and the largest effort to date in extracting spinal cord shape metrics from MRI data. We identified novel genetic loci linked to spinal cord morphological traits and observed sex-specific associations. Our findings indicated that the spinal cord sizes have significant genetic overlap with brainstem and subcortical volumes including the ventral diencephalon, pallidum, and the thalamus. Leveraging health-relation outcomes, we provide evidence that the spinal cord does not just reflect neurological health but also metabolic and wide-ranging systemic disease conditions, highlighting its importance as the conduit of nerve signals and its multifactorial vulnerability.

Our approach represents a substantial advance on previous work that quantified cervical spinal cord structure in modest sample sizes, including a small subset of UKB data (n=804)^48^, by analyzing the spinal cord morphology at the individual vertebral level across C1 to C3 of over 40,000 participants. This was made possible by an artificial intelligence-enabled automated imaging analysis pipeline underpinned by a model trained on high-quality spinal cord labels of healthy and pathological cases, enhancing robustness to anatomical variations and image artifacts. This optimized pipeline was designed to handle a broader range of real-world imaging conditions without any manual intervention needed. We observed that discontinuous segmentations, often caused by signal dropout or artifacts, are corrected during vertebral labelling as the trained model preserved the expected tubular structure of the spinal cord. Neck angle and pathological variations among the older age range of UKB participants were handled reliably. Through rigorous manual QC check and validation, we showed that this pipeline significantly improved the accuracy of C1 labelling, closely reflecting known anatomical patterns, such as the shorter length of C1 compared to C2, and enabled detection of subtle morphological trends like decreasing AP and increasing RL diameters from C1 to C3 regions^49^. Furthermore, we implemented neck angle correction to account for individual curvature and improve metric estimation accuracy. We demonstrated that the concordance across C1–C3 levels, reproducibility between longitudinal scans, and correlations with structurally connected brain regions collectively offer substantial evidence of the success of this automated approach. This paves the way for future studies which will require these types of automated frameworks to enable broader application.

We established that cervical spinal cord morphology exhibits moderate to high SNP-based heritability ranging from h^2^=0.16-0.42. Previously, only a small twin study^30^ had suggested that spinal cord CSA and diameters were highly heritable (h^2^=0.85-0.91). Interestingly, we found that eccentricity, a metric reflecting circular or flattened shape, exhibited the highest heritability. Clinical observations have noted that patients with ataxia tend to have flatter spinal cords, and this metric appears to remain stable over time^12^. Our analysis of population-level data further supports the notion that eccentricity is largely developmentally determined and remains stable with aging, in contrast to size-related metrics such as CSA and diameters, which show proportional changes with aging. These size-based metrics are likely more sensitive to pathological processes, as they are more modifiable over time. The observed decline in heritability from C1 to C3 across all phenotypes may be attributed to both underlying genetic trends and decreasing sample size from C1 to C3. Notably, C1 lies closest to the brainstem, a highly heritable structure, which may explain its stronger genetic signal. Scans covering regions beyond C3 will help to further explore these patterns.

Our large-scale genetic investigation identified 179 genetic variants associated with spinal cord shape metrics. Many of these variants have been previously reported to be associated with neurological, cardiovascular, and anthropometric phenotypes. One highly associated variant, rs1126642 on chromosome 17, leads to a missense mutation within exon 5 of *GFAP*, an intermediate filament cytoskeletal gene highly expressed in glial cells, particularly astrocytes. *GFAP* is central to both brain and spinal cord development and pathology, but is also expressed in non-neural tissue^50^, highlighting a broad biological importance for this GWAS signal. Sex-specific analyses revealed both overlapping and distinct genetic influences on spinal cord shape in males and females. While most loci identified in the combined analysis were also detected in the sex-stratified GWAS, three SNPs, one of which is located in the androgen receptor gene, showed significant SNP-by-sex interaction effects, indicating potential sex-dependent genetic regulation of spinal cord morphology.

Our gene-based analysis using positional and functional gene-based mapping approaches identified a set of high-confidence genes that are likely to contribute to variation in cervical spinal cord morphology. Notably, we identified 96 genes which were supported by at least three gene-based approaches, several of which were further prioritized based on colocalization and heterogeneity thresholds, pointing to shared causal variants between gene expression and morphological traits. Among these, *SGTB*, *TGFA*, and *ROR2* emerged as top ranked and biologically plausible gene candidates. For example, *SGTB*, encoding a small tetratricopeptide repeat co-chaperone, was implicated by five gene-based approaches and showed strong evidence of colocalization in brain tissues for CSA and diameter measures. *SGTB* has a key role in shaping neurite morphology and promoting neuron survival^51^ and has been implicated in GWAS of neuroimaging phenotypes^14,37^ and white matter integrity^52^. Meanwhile, *TGFA*, a ligand of the epidermal growth factor receptor (EGFR), promotes proliferation and survival in epithelial and neural tissues during development^53,54^, and is expressed in the developing spinal cord^55^. Finally, *ROR2* is a member of the tyrosine-protein kinase receptor family with an essential role in axial skeletal development and vertebral patterning. Importantly, loss-of-function mutations in *ROR2* cause autosomal recessive Robinow syndrome^56^ characterized by vertebral segmentation defects and skeletal malformations, further highlighting its involvement in determining spinal cord morphology.

We observed moderate negative phenotypic correlations between cervical spinal cord size and several neurological conditions, including MS and neuropathies. These findings were consistent with clinical evidence that spinal cord atrophy has prognostic value for MS progression and treatment response^57,58^, and spinal cord atrophy rate may help distinguish MS subtypes^59^. We also observed negative correlations between cervical spinal cord size and mono- and polyneuropathies, consistent with spinal cord atrophy reported in both early and severe peripheral neuropathy^60,61^. This may provide some insights into the phenotypic association we found with both type 1 and type 2 diabetes in our study^62^. Neuropathies are clinically difficult to quantify and measure over time, hence spinal cord biomarkers may serve as a useful tool going forward. Additionally, phenotypical correlations with viral infections, many of which are neurotropic^63,64^, musculoskeletal disorders such as osteoporosis and gout arthropathies, and essential hypertension suggest that spinal cord morphology may serve as a potential radiological biomarker for a broad range of disease states.

Genetically, PD was previously shown to have positive overlap with basal ganglia, while ADHD was negatively linked to the brainstem and ventral diencephalon^17^. A previous study identified shared molecular genetic factors between subcortical volumes and PD, highlighting potential links to inflammation, mitophagy, vesicle-trafficking, and the hypothalamic-pituitary-adrenal pathway^65^. We extend these association with PD and ADHD to the spinal cord (C1–C3) with consistent trend and directions. However, Mendelian randomization analyses did not provide robust evidence to support a causal effect of PD or ADHD on spinal cord phenotypes. Follow-up analysis is required to further examine these relationships in depth, especially in non-European populations and clinical cohorts.

In the brain, previous studies have reported SNP-based heritability estimates ranging from 9% to 33% for intracranial and subcortical brain volumes^17,37^, while the whole brainstem has a heritability of approximately 48%^16^. Our findings suggest that spinal cord morphometry lies within a comparable range. We observed stronger genetic associations with other brain regions along the motor-sensory pathways, consistent with known neuroanatomical and imaging evidence^30,48^. Note that we used the total volume of the cerebellum to compute the genetic correlation, as GWAS of total cerebellar white matter has not been reported. Similar to brain phenotypes, we identified associations between spinal cord morphology and cardiovascular and metabolic conditions^66,67^. Among neurological disorders, MS showed the strongest phenotypic association with spinal cord morphology, although no significant genetic association was observed, similar to previously reported a lack of association between polygenic risk for MS and brain MRI^68^.

One limitation of this study is that the discovery GWAS sample consists of mainly European individuals aged 45 years and older. Previous studies indicate that spinal cord morphometry has greater dynamic range earlier in life and may thus be even more potent as a biomarker in younger age cohort studies. Another limitation is that our findings in European populations may not generalize to individuals of other ancestries. We examined whether SNPs identified in the European-ancestry GWAS of spinal cord shape metrics were also associated with the same traits in non-European UKB individuals. However, these analyses were limited in scale and substantially underpowered. Consequently, no significant associations were identified in individuals of non-European ancestries, and genome-wide discovery was not feasible in these populations.

In summary, this study presents the first large-scale GWAS to uncover novel genetic loci associated with spinal cord shape metrics, providing new insights into the genetic architecture of spinal cord. Our findings, combining imaging and genetic evidence, highlight the importance of further investigating the spinal cord’s involvement in human health.

## METHODS

### UK Biobank

The UKB is a large-scale biomedical database containing in-depth genetic, imaging, and health information from over 500,000 participants. Access to all UKB (https://www.ukbiobank.ac.uk/) data was granted on June 18th, 2019, application #36610. UKB has ethics approval from the North West Multi–centre Research Ethics Committee. This study was approved by the Walter and Eliza Hall Institute of Medical Research (WEHI), Human Research Ethics Committee (HREC reference 17/09LR).

### MRI acquisition and image sample

MRI image acquisition was done following the UKB imaging protocol (https://biobank.ctsu.ox.ac.uk/crystal/crystal/docs/brain_mri.pdf). In brief, brain imaging was acquired across four identical imaging centres in the UK. T1-weighted (T1-w) structural scans were acquired using a 3D MPRAGE protocol (voxel size = 1 × 1 × 1 mm^3^, matrix size = 208 × 256 × 256, inversion time (TI)/TR = 880/2,000 ms, in-plane acceleration = 2, total scan time = 4 min 54 s).

We used the late 2023 release of brain MRI data (downloaded prior to March 2024). The UKB standard QC^69^ pipeline was applied to exclude any images of poor quality and severe artefacts (labelled as unusable). Although T1-w images were processed using the UKB image processing pipeline, our study used the unprocessed data with the largest field of view to preserve the spinal cord region. After the initial standard QC, a total of 46,366 images and an additional 4,578 repeat scans underwent further spinal cord processing.

### Spinal cord processing pipeline

We implemented an optimized deep learning–based spinal cord processing pipeline for upper spinal cord labelling (https://github.com/art2mri/Enigma-SC), designed for large-scale and reliable spinal cord shape analysis (Fig. 1a). The pipeline begins with spinal cord segmentation using the *sct_deepseg_sc* command from the Spinal Cord Toolbox (SCT)^23^, followed by vertebral labelling of C1–C3 using a pretrained nnu-Net^34^ deep learning model specifically tuned for the upper spinal cord^33^. From the resulting segmentation masks and vertebral labels, spinal cord shape metrics are extracted. This pipeline addresses several key challenges: (1) it enables fully automatic and accurate identification of C1–C3 without the need for manual correction; (2) robustness to pathological variations, neck angle differences, and partial coverage; and (3) it significantly improves the accuracy of C1 labelling by leveraging a training set curated with high-quality C1 annotations. In addition, CSA measurements were corrected for neck curvature using AP and RL angles derived from the shape analysis.

### Spinal cord phenotype QC

To address the problem of image quality variability within the same scan and to maximize sample size, we implemented a multi-step QC procedure (detailed in Supplementary Section 11). First, T1-weighted images with poor overall quality were excluded based on the standard UKB QC flag. Next, given spinal cord phenotypes had not been extracted in large-scale, we used a conservative manual QC approach (Supplementary section 11.2). Vertebral-level-specific QC was applied by manually inspecting all vertebral labels from C1 to C3 to exclude mislabelled or poor-quality regions. Finally, we removed a small number of outlier points with large discrepancies between metrics derived from the spinal cord segmentation mask vs. the vertebral labels, typically caused by segmentation inaccuracies (Supplementary section 11.3). This process enabled rigorous exclusion of inaccurate data while preserving as much high-quality information as possible to maintain statistical power for downstream GWAS.

### Phenotyping robustness and reproducibility evaluation

We considered an alternative processing method to ensure the derived spinal cord phenotypes are not prone to processing method choices. Atlas based analysis was performed by adding an additional registration step after vertebral labelling. In brief, the spinal cord of the PAM50 atlas^70^ was straightened and aligned to the native space. Shape metrics were extracted from the registered PAM50 vertebral level images. Correlations between the atlas-based method vs. the native-space spinal cord processing pipeline used in the current study are calculated (Supplementary section 2.2).

Reproducibility evaluation was performed by calculating the correlations between repeat scans. A small subset of participants underwent repeat scan (1-7 years after the first scan). Spearman’s correlations between the first and second scans were calculated to assess their repeatability, stratified by scan intervals (Supplementary section 2.1).

### Spinal cord IDP and covariate selection

SCT outputs nine shape metrics per vertebral level: CSA, eccentricity, AP diameter, RL diameter, AP angle, RL angle, orientation, solidity, and total length. However, not all of these reflect biologically meaningful variation suitable for GWAS. We selected relevant phenotypes based on two key criteria: (1) stability across processing steps: metrics such as orientation, angles, sum length can be influenced by image straightening or rotation, making them modifiable during processing; and (2) repeatability across imaging sessions: measures that reflect transient conditions, such as head positioning during a specific scan, were excluded. Based on our robustness and repeatability analyses, we identified four phenotypes, CSA, eccentricity, AP diameter, and RL diameter, as consistently measurable and biologically interpretable, and thus selected them for GWAS to investigate their genetic architecture.

To confirm that these phenotypes captured both shared and distinct information across C1–C3, we assessed their interrelationships using linear regression models, adjusting for sex, age, height, ICV, imaging site, and genetic principal components 1–10. Principal component analysis was also conducted to explore the underlying structure of these shape metrics and to identify statistical outliers for exclusion (Supplementary Section 4).

Previous spinal cord studies suggested that age, sex, brain size and ICV were relevant covariates that were recommended to be included for normalization^28^. We performed additional stepwise regression analysis including additional potential confounding factors including scanning site and T1 signal-noise-ratio (Data-Field 25734) to verify relevant covariates to be included in the genome wide association analysis (Supplementary Section 4). We also performed genome-wide association analysis without correcting for the effect of body height as sensitivity analysis.

### Genotyping, quality control and imputation

Genotyping, QC and imputation for the UKB cohort were performed by UKB and have been described previously^71^. We used the imputed genetic dataset based on the TOPMed r2 reference panel (Data-Field: 21007), which includes 97,256 deeply sequenced human genomes and approximately 308 million genetic variants from the autosomal and X chromosomes^72^. All QC and analyses of genetic data were performed on the UKB Research Analysis Platform (RAP).

Filtering of the genetic data was undertaken using PLINK2 (v2.0.0-a.6.9)^73^. Variants were excluded if they had a MAF < 0.001 or deviated from Hardy–Weinberg equilibrium (*P* < 1 × 10⁻¹⁵). Imputation quality metrics were extracted from the VCF “helper files”, and variants identified for exclusion if they had low imputation quality, defined as R^2^ < 0.6, or empirical R^2^ < 0.6 (directly genotyped variants only). We conducted the analysis on the autosomes (chromosomes 1 to 22) and the X chromosome.

Sample QC was undertaken using metrics made available by UKB (Data Fields 22001; 22019; 22027; 22021). We excluded individuals who had discordant self-reported and genetically inferred sex, or sex chromosome aneuploidy; or were flagged by UKB prior to imputation for high heterozygosity, high missingness (>5%); or having an excessive number of relatives (≥10) in the cohort. Individuals who had withdrawn consent were also excluded. To reduce potential confounding, we further excluded individuals with conditions affecting the cervical spine, as identified by ICD-10 diagnostic codes (Data-Field 41270) and operative procedure (OPCS-4, Data-Field 41272), with detailed codes provided in Table S1.

Genetic ancestry principal components (PCs) were obtained from the Pan-UK Biobank project (https://pan.ukbb.broadinstitute.org)^35^. Individuals of European, C/SA, AFR, and EAS ancestries were selected for inclusion in the analysis. Individuals from other ancestry groups were excluded due to limited small sample sizes (<200 individuals per group).

### Genetic association analyses for spinal cord shape metrics

GWAS were conducted for the 12 spinal cord shape metrics using REGENIE (v2.1.0)^36^. For the discovery GWAS, analyses were restricted to individuals of European genetic ancestries, who passed all genetic and imaging QC. All GWAS were adjusted for age, sex, age², genotyping batch, imaging site, ICV, height, inverted signal-to-noise ratio in T1-weighted images, and the first ten genetic PCs to account for population stratification. Rank inverse normal transformation was applied to all phenotypes, and an additive genetic model was assumed. The REGENIE method involves two steps. In Step 1, a whole-genome regression model was fitted, using only directly genotyped variants with MAF > 0.01% to generate polygenic predictions for each phenotype, which were further decomposed into 23 chromosome-specific predictions using a leave-one-chromosome-out (LOCO) scheme. In Step 2, imputed UKB genotype data were tested for association with the phenotype conditional upon the predictions from Step 1. For analyses of the X chromosome, hemizygous males were coded as homozygous. Quantile–quantile (Q–Q) plots, genomic inflation factor (λ), and LDSC^42^ regression intercept were used to evaluate potential systematic bias in the association results.

### Identification of significant independent SNPs

Significant independent SNPs were identified using the stepwise model selection procedure implemented in GCTA-COJO (v1.94.4)^74^, with LD estimated from the same European-ancestry discovery cohort. A 10 megabase (Mb) window was used to define regions of complete LD, and genome-wide significance was set at the conventional threshold of *P* < 5 × 10⁻⁸.

To confirm all variants identified by GCTA-COJO were truly independent signals, we further performed conditional analyses using REGENIE^36^, retaining all parameters and Step 1 models from the unconditioned analysis. In Step 2, we conditioned on the variant with the lowest P-value within a 1 Mb window to assess whether the association for each independent signal remained genome-wide significant (*P* < 5 × 10⁻⁸).

### Reporting of significant SNPs and loci

To account for multiple testing across the 12 spinal cord shape metrics, a Bonferroni-corrected threshold of *P* < 4.17 × 10⁻⁹ (5 × 10⁻⁸/12) was applied for final reporting and downstream follow-up of SNPs. All significant independent SNPs were annotated using Functional Annotation of Variants - Online Resource (FAVOR)^75^, with the variant function, and (nearest) gene, based on GENCODE annotations^76^.

To examine the overlap of findings across the 12 traits, we grouped independent SNPs into loci based on their proximity and linkage disequilibrium. Specifically, SNPs were considered part of the same locus if they were within 250 kilobase (kb) of each other and had a pairwise linkage disequilibrium of r² > 0.1.

To determine whether our independent SNPs had been previously reported, we downloaded the GWAS Catalog association summary statistics (accessed 22 July 2025) and cross-referenced them with the GWAS Catalog. To improve the integration of GWAS Catalog phenotype categories, we used the EFO Ontology Trait Mappings (https://www.ebi.ac.uk/gwas/api/search/downloads/trait_mappings) to align the traits with their corresponding ontology terms and parent terms.

### SNP-based heritability

We used LDSC (v.1.0.1)^42^ to estimate SNP-based heritability (*h^2^*) for each spinal cord shape metrics using GWAS summary data. Precomputed LD scores from the 1000 Genomes Project European reference population^77^. were used as the reference panel, and analyses were restricted to HapMap3 SNPs^78^.

### Sex-specific GWASs and Sex interaction analyses

To investigate sex differences in the genetic architecture of spinal cord shape metrics, we conducted sex-specific GWAS using REGENIE, and identified independent SNPs using GCTA-COJO as in the sex-combined discovery analyses. We defined sex-specific signals as independent SNPs meeting *P* < 4.17 × 10⁻⁹ in one of the sex-specific analyses, but not in the corresponding sex-combined analysis. To confirm these sex-specific signals, we verified that the top SNP was not in LD (r² > 0.1) with any independent SNP identified through the sex-combined GWAS.

For all independent, significant SNPs identified through either the sex-combined, or sex-specific analyses, we sought evidence of SNP-by-sex interaction, by repeating REGENIE’s step 2, with the addition of the interaction flag. REGENIE’s gene by environment interaction model uses a Wald test with sandwich estimators of variance for variants whose minor allele count (MAC) > 1,000, with a heteroskedastic linear model used for variants of lower frequencies^79^. We defined evidence of a SNP-by-sex interaction based on the interaction term p-value meeting a Benjamini-Hochberg false discovery rate (FDR) adjusted threshold of 0.05.

### Associations in non-European ancestries

We accessed whether the independent significant SNPs identified in the European-ancestry GWAS of spinal cord shape metrics were also associated with the same traits in individuals of C/SA, AFR, and EAS ancestry from the UKB. Association analyses were conducted separately for each ancestry group using REGENIE, following the same procedures as in the discovery analysis. To focus on common variants, we excluded SNPs with a MAF < 0.05 within each respective population cohort.

Replication was defined as achieving significance at a Bonferroni-corrected threshold, adjusted for the number of loci tested (*P* < 0.05/179). We then compared effect sizes between the European and non-European groups by calculating Spearman’s Rank Correlation coefficients and the proportion of SNPs with concordant effect directions. A two-sided binomial test assessed whether observed concordance differed from that expected under random directionality.

### Gene-based tests

GWAS summary statistics for each of the 12 spinal cord traits were lifted over from genome build hg38 to hg19 using the UCSC LiftOver tool with the appropriate chain files.

## MAGMA

MAGMA (Multi-marker Analysis of GenoMic Annotation) was used to identify risk genes near genome-wide significant loci for the 12 spinal cord shape phenotypes. This approach assigns variants to genes based on their physical proximity to a defined set of genic coordinates. We mapped variants to protein coding genes obtained from the MAGMA website (https://cncr.nl/research/magma/). Genic coordinates extended 10kb upstream and downstream to capture regulatory variation. After mapping variants to genes, gene-based P-values were calculated using the SNP-wise mean model. We used samples from the 1000 Genomes Project (Phase 3) as the LD reference panel.

### mBAT-combo

We performed a gene-based analysis utilizing the multivariate Set-Based Association Test (mBAT-combo) within GCTA version 1.94.1. This method enhances the detection power for multi-SNP associations, especially in the presence of masking effects, by combining mBAT and fastBAT test statistics through a Cauchy combination approach. This allows for the integration of different test statistics without prior knowledge of their correlation structure, thereby maximizing overall power regardless of masking effects at specific loci. We used the European subsample (*n* = 503) from Phase 3 of the 1000 Genomes Project as the LD reference panel, applying the fastBAT default LD cut-off of 0.9. A gene list of 19,899 protein-coding genes was utilized to map base pair positions according to genome build hg19.

### Transcriptome-wide association study

We performed a transcriptome-wide association study (TWAS) using the FUSION method, which integrates GWAS summary statistics with gene expression prediction models to identify associations between gene expression and the 12 spinal cord phenotypes. We utilized brain gene expression weights from the PsychENCODE project, which includes expression data from the dorsolateral prefrontal cortex. We incorporated LD information from the 1000 Genomes Project Phase 3 to estimate the genetic component of gene expression. This genetic component was then tested for association with each spinal cord phenotype using GWAS summary data. To correct for multiple testing, we applied the Bonferroni correction (i.e., 0.05/number of tests performed). We used the COLOC R package^80,81^, implemented using FUSION, to identify TWAS associations (i.e., trait-gene expression associations) that are likely to share a causal SNP. Colocalisation calculates posterior probabilities (PP) to assess whether individual lead SNPs within a significant TWAS locus are: (i) independent signals (PP3) (e.g., two causal SNPs in LD, one affecting transcription and the other affecting spinal cord morphology), or (ii) shared signals (PP4) (e.g., a single causal SNP affecting both transcription and spinal cord morphology).

### Summary-based Mendelian Randomisation

We used Summary data-based Mendelian Randomization (SMR) to integrate GWAS and expression quantitative trait loci (eQTL) data from human brain and blood samples. This method leverages summary-level data in a Mendelian Randomisation statistical framework to test for pleiotropic associations between gene expression and GWAS data. We used whole blood expression data from the eQTLGen consortium and brain expression data from the MetaBrain resource^82^. To assess the presence of linkage or pleiotropy, we performed the Heterogeneity in Dependent Instruments (HEIDI) test alongside SMR, evaluating effect size heterogeneity between the GWAS and eQTL summary statistics. The HEIDI test is designed to distinguish between pleiotropy and linkage by assessing heterogeneity in the association signals. A HEIDI P-value threshold of 0.05 was used to determine the likelihood of linkage influencing the observed association.

### Gene set enrichment analysis with GSA-MiXeR

Gene set analysis was conducted using GSA-MiXeR to quantify the enrichment of genetic signals across 12 spinal cord phenotypes within gene ontology (GO) categories. GWAS summary statistics were analyzed with GSA-MiXeR using default parameters as described by Frei et al.^83^. The method employs a competitive approach to gene set analysis, estimating fold enrichment of partitioned heritability through maximum likelihood optimization, accounting for LD, MAF, and polygenicity. We used the FDR (*P* < 0.05) to control for multiple comparisons in MAGMA analyses. Gene ontology sets were prioritized based on their fold enrichment, allowing assessment of biologically relevant gene sets associated with each spinal cord phenotype.

### Genetic correlations with other brain phenotypes and diseases

We used cross-trait LDSC to estimate genetic correlations within spinal cord shape metrics and between these metrics and a range of brain IDPs, including measures from the cortical, subcortical, and brainstem regions, as well as various diseases. LDSC accounts for sample overlap and population stratification, providing robust estimates of shared genetic architecture. Phenotypes were selected based on prior evidence of relevance to spinal cord structure, observed phenotypic associations, and data availability. GWAS summary statistics used in these analyses are listed in the Table S25. Multiple testing correction for all cross-trait analyses was performed using the Benjamini–Hochberg procedure to control the FDR (*P* < 0.05).

### Mendelian randomization

For the traits which showed significant genetic correlations with spinal cord metrics, and had a sufficient number (β3) of associated genetic variants to act as instruments, we conducted Mendelian randomization (MR) to explore potential causal effects on spinal cord shape metrics. MR is a form of instrumental variable analysis whereby genetic variants (“instruments”) are used to examine the causal relationship between one trait (“exposure”) and another (“outcome”). There are three core assumptions central to MR: 1) the genetic instruments must be significantly associated with exposure of interest; 2) no confounding should exist between the genetic instruments and the outcome; and 3) the genetic instruments must not be associated with the outcome, via any pathway, other than through the exposure (i.e. no horizontal pleiotropy).

We utilised a two-sample MR approach, which is intended to leverage GWAS summary statistics from non-overlapping samples for exposure and outcome. For all exposure traits (i.e. the non-spinal cord phenotypes), we used the same published GWAS summary statistics as in the genetic correlation analyses (Table S25). We note the GWAS for these traits were large meta-analyses which, with the exception of ADHD, included UKB samples. This has the potential to lead to bias estimates from two-stage MR methods, depending on the level of overlap^84,85^.

Our primary MR analyses were performed using the inverse-variance weighted (IVW) method^43^, as implemented in the TwoSampleMR R package^86,87^. Genetic instruments for each exposure were obtained using LD-clumping, using a 10,000 kb window, a clumping P-value threshold of 5 × 10⁻⁸, an r² cut-off of 0.001, and LD estimated from 1000 Genomes European samples. When extracting instruments from the outcome (spinal cord phenotypes) GWAS summary statistics, proxies were not considered, in place of unavailable SNPs. Results passing the Benjamini–Hochberg procedure to control the FDR at *P* < 0.05 were considered significant.

### Mendelian randomization sensitivity analyses

In addition to the standard IVW approach, we repeated the MR analyses using a series of other methods and approaches, allowing us to evaluate the validity of the genetic instruments and assess potential violations of the core MR assumptions.

First we repeated the MR analyses using the MR-Egger^46^, weighted median^44^, and weighted mode^45^ methods, all implemented in the TwoSampleMR R package^86,87^. These methods provide consistent estimates, even where not all of the genetic instruments are valid. We examined the MR-Egger intercept to test for directional pleiotropy; a non-zero intercept suggests the presence of directional horizontal pleiotropy, which violates assumption 3, and could bias the MR estimate. We also assessed heterogeneity of instrument effects using Cochran’s Q statistic; significant heterogeneity may also be indicative of horizontal pleiotropic effects.

As an alternative approach, we implemented the Causal Analysis Using Summary Effect estimates (CAUSE)^47^ model (v1.2.0, https://github.com/jean997/cause). CAUSE accounts for shared genetic effects between traits and provides estimates robust to both correlated and uncorrelated horizontal pleiotropy. CAUSE can also account for shared genetic effects that might arise from sample overlap. Briefly, in step 1 we estimated the nuisance parameters, using the default settings, and 1 million randomly selected genome-wide variants. In step 2 (estimation of causal effects), we used filtered and pruned variants (*r²* < 0.001 within 10 Mb windows) based on two P-value thresholds: *P* < 0.001 and *P* < 5 × 10⁻⁸.

Finally, to investigate potential bidirectional relationships, we conducted reverse MR analyses. In these analyses, we used genetic instruments for spinal cord phenotypes as the exposure, with our original exposure traits as the outcome. Spinal cord genetic instruments were selected as those identified in the COJO analysis at a significance threshold of *P* < 5 × 10⁻⁸/12. Reverse MR was carried out for inverse variance weighting, MR-Egger, weighted median, and weighted mode methods.

### Phenotypic correlations with other brain phenotypes and health-related outcomes

A total of 41 volumes were selected including four brainstem subregions: pons, medulla, midbrain, superior cerebellar peduncle (SCP) (Category 191); 9 subcortical regions: caudate, cerebellum (white matter [WM] + grey matter [GW]), hippocampus, pallidum, putamen, thalamus, ventral diencephalon, amygdala, accumbens (Category 191); 28 cortical BA volumes in the left and right hemisphere (Category 195). Phenotypic association with ICV was also calculated as a reference. Inverse normalization transformation was applied to spinal cord shape metrics, then linear regression model was used to calculate the associations between spinal cord IDPs and other brain volumes, correcting for the effects of sex, age, and genetic principal components 1–10.

ICD-10 outcomes were extracted from the UKB (Category 1712), incorporating data from multiple sources: primary care records, hospital inpatient admissions, death registries, and self-reported medical conditions. These ICD-10 codes were then mapped to the corresponding Phecode Map X (https://phewascatalog.org/phewas/#phex). Because a single ICD-10 code can map to multiple PhecodeX entries, we selected the most appropriate PhecodeX based on the following criteria: (1) preserving the ICD-10 category, and (2) best aligning with the specificity of the original ICD-10 code (Table S26). This mapping process yielded 778 unique PhecodeX disease codes from 1,165 ICD-10 codes for subsequent association analysis.

The extracted data were converted into binary format, where 1 indicated the presence of a diagnosis and 0 indicated absence. Logistic regression models were used to assess associations between spinal cord phenotypes and each PhecodeX condition. Covariates included sex, age, height, ICV, imaging site, and genetic PCs 1–10. A total of 9,336 tests were performed across 12 spinal cord phenotypes. An FDR corrected threshold of *P* < 0.05 was considered significant.

## Data Availability

All genetics, health related information, and imaging data described in this paper are publicly available to registered researchers through the UKB data-access protocol. Additional information about registration for access to the data are available at: https://www.ukbiobank.ac.uk/enable-your-research/apply-for-access.
GWAS summary statistics will be submitted to the GWAS catalogue upon publication.
The supplementary table is available for downloading at https://osf.io/9s6gk/.

https://osf.io/9s6gk/

## Acknowledgement

This work was also made possible through the Victorian State Government Operational Infrastructure Support and Australian Government National Health and Medical Research Council Independent Research Institute Infrastructure Support Scheme. The data used in the study are obtained from UKB Resource (https://www.ukbiobank.ac.uk/) under Application Number 36610. We would like to thank the participants and their families, without whom these studies would not have been possible. We also like to thank the WEHI AI initiative and the technical support provided by WEHI’s research computing team.

## Funding

ZFG is supported by an Australian National Health and Medical Research Council (NHMRC) EL1 Investigator Grant (2034743).

MB was supported by an Australian National Health and Medical Research Council (NHMRC) Investigator Grant (GNT1195236).

## Competing interests

The authors declare that they have no competing interests.

## Data availability statement

All genetics, health related information, and imaging data described in this paper are publicly available to registered researchers through the UKB data-access protocol. Additional information about registration for access to the data are available at: https://www.ukbiobank.ac.uk/enable-your-research/apply-for-access.

GWAS summary statistics will be submitted to the GWAS catalogue upon publication. The supplementary table is available for downloading at https://osf.io/9s6gk/.

## Code availability

The spinal cord image pre-processing pipeline is publicly available at: https://github.com/art2mri/Enigma-SC

No custom code was used in this study. Publicly available software tools were used to perform genetic analyses and are referenced throughout the paper.

## Notes

### Competing Interest Statement

The authors have declared no competing interest.

## References

1. Kandel, E. R., Koester, J. D., Mack, S. H. & Siegelbaum, S. A. Sensory-Motor Integration in the Spinal Cord. in Principles of Neural Science, 6e (McGraw Hill, New York, NY, 2021).

2. Purves, D., Augustine, G. J. & Fitzpatrick, D. Sinauer Associates. (Sunderland (MA, 2001).

3. Tartaglino, L. M. et al. Multiple sclerosis in the spinal cord: MR appearance and correlation with clinical parameters. Radiology 195, 725–732 (1995).

4. Lycklama, G. et al. Spinal-cord MRI in multiple sclerosis. Lancet Neurol. 2, 555–562 (2003).

5. Bede, P. et al. Spinal cord markers in ALS: diagnostic and biomarker considerations. Amyotroph. Lateral Scler. 13, 407–415 (2012).

6. Querin, G. et al. Multimodal spinal cord MRI offers accurate diagnostic classification in ALS. J. Neurol. Neurosurg. Psychiatry 89, 1220–1221 (2018).

7. Freund, P. et al. MRI in traumatic spinal cord injury: from clinical assessment to neuroimaging biomarkers. Lancet Neurol. 18, 1123–1135 (2019).

8. Marquez de la Plata, C. D., et al. Diffusion tensor imaging biomarkers for traumatic axonal injury: analysis of three analytic methods. J. Int. Neuropsychol. Soc. 17, 24–35 (2011).

9. Basbaum, A. History of spinal cord “pain” pathways including the pathways not taken. Front. Pain Res. (Lausanne) 3, 910954 (2022).

10. Todd, A. J. Neuronal circuitry for pain processing in the dorsal horn. Nat. Rev. Neurosci. 11, 823–836 (2010).

11. Harding, I. H. et al. Brain structure and degeneration staging in friedreich ataxia: Magnetic resonance imaging volumetrics from the ENIGMA-ataxia working group. Ann. Neurol. 90, 570–583 (2021).

12. Rezende, T. J. R. et al. Progressive Spinal Cord Degeneration in Friedreich’s Ataxia: Results from ENIGMA-Ataxia. Mov. Disord. 38, 45–56 (2023).

13. Elliott, L. T. et al. Genome-wide association studies of brain imaging phenotypes in UK Biobank. Nature 562, 210–216 (2018).

14. Smith, S. M. et al. An expanded set of genome-wide association studies of brain imaging phenotypes in UK Biobank. Nat. Neurosci. 24, 737–745 (2021).

15. Zhao, B. et al. Large-scale GWAS reveals genetic architecture of brain white matter microstructure and genetic overlap with cognitive and mental health traits (n = 17,706). Mol. Psychiatry 26, 3943–3955 (2021).

16. Elvsåshagen, T. et al. The genetic architecture of human brainstem structures and their involvement in common brain disorders. Nat. Commun. 11, 4016 (2020).

17. García-Marín, L. M. et al. Genomic analysis of intracranial and subcortical brain volumes yields polygenic scores accounting for variation across ancestries. Nat. Genet. 1–12 (2024) doi:10.1038/s41588-024-01951-z.

18. Chambers, T. et al. Genetic common variants associated with cerebellar volume and their overlap with mental disorders: a study on 33,265 individuals from the UK-Biobank. Mol. Psychiatry 27, 2282–2290 (2022).

19. Jackson, V. E. et al. Multi-omic spatial effects on high-resolution AI-derived retinal thickness. Nat. Commun. 16, 1317 (2025).

20. Kun, E. et al. The genetic architecture and evolution of the human skeletal form. Science 381, eadf8009 (2023).

21. Wen, J. et al. The genetic architecture of biological age in nine human organ systems. *Nat*. Aging 4, 1290–1307 (2024).

22. Valošek, J. & Cohen-Adad, J. Reproducible Spinal Cord Quantitative MRI Analysis with the Spinal Cord Toolbox. Magn. Reson. Med. Sci. (2024) doi:10.2463/mrms.rev.2023-0159.

23. De Leener, B. et al. SCT: Spinal Cord Toolbox, an open-source software for processing spinal cord MRI data. Neuroimage 145, 24–43 (2017).

24. Paquin, M.-Ê. et al. Spinal cord gray matter atrophy in amyotrophic lateral sclerosis. AJNR Am. J. Neuroradiol. 39, 184–192 (2018).

25. Karbasforoushan, H., Cohen-Adad, J. & Dewald, J. P. A. Brainstem and spinal cord MRI identifies altered sensorimotor pathways post-stroke. Nat. Commun. 10, 3524 (2019).

26. Parizel, P. M. et al. Trauma of the spine and spinal cord: imaging strategies. Eur. Spine J. 19 **Suppl 1**, S8–17 (2010).

27. Hubertus, V. et al. In vivo imaging in experimental spinal cord injury - Techniques and trends. Brain Spine 2, 100859 (2022).

28. Labounek, R. et al. Body size and intracranial volume interact with the structure of the central nervous system: A multi-center in vivo neuroimaging study. Imaging Neuroscience (2025) doi:10.1162/imag_a_00559.

29. Engl, C. et al. Brain size and white matter content of cerebrospinal tracts determine the upper cervical cord area: evidence from structural brain MRI. Neuroradiology 55, 963–970 (2013).

30. Solstrand Dahlberg, L., Viessmann, O. & Linnman, C. Heritability of cervical spinal cord structure. Neurol. Genet. 6, e401 (2020).

31. Rezende, T. J. R. et al. Genotype-specific spinal cord damage in spinocerebellar ataxias: an ENIGMA-Ataxia study. J. Neurol. Neurosurg. Psychiatry 95, 682–690 (2024).

32. Stroman, P. W. et al. The current state-of-the-art of spinal cord imaging: methods. Neuroimage 84, 1070–1081 (2014).

33. Enigma-SC pipeline. https://github.com/art2mri/Enigma-SC

34. Isensee, F., Jaeger, P. F., Kohl, S. A. A., Petersen, J. & Maier-Hein, K. H. nnU-Net: a self-configuring method for deep learning-based biomedical image segmentation. Nat. Methods 18, 203–211 (2021).

35. Karczewski, K. J. et al. Pan-UK Biobank GWAS improves discovery, analysis of genetic architecture, and resolution into ancestry-enriched effects. bioRxiv (2024) doi:10.1101/2024.03.13.24303864.

36. Mbatchou, J. et al. Computationally efficient whole-genome regression for quantitative and binary traits. Nat. Genet. 53, 1097–1103 (2021).

37. Satizabal, C. L. et al. Genetic architecture of subcortical brain structures in 38,851 individuals. Nat. Genet. 51, 1624–1636 (2019).

38. Buniello, A. et al. The NHGRI-EBI GWAS Catalog of published genome-wide association studies, targeted arrays and summary statistics 2019. Nucleic Acids Res. 47, D1005–D1012 (2019).

39. Biallelic loss-of-function variants in RABGAP1 cause a novel neurodevelopmental syndrome Oh. Rachel Youjin et al. Genetics in Medicin.

40. Eden, J., et al. RABGAP1 acts as a sensor to facilitate sorting and processing of amyloid precursor protein. bioRxiv (2024) doi:10.1101/2024.07.11.602925.

41. Sollis, E. et al. The NHGRI-EBI GWAS Catalog: knowledgebase and deposition resource. Nucleic Acids Res. 51, D977–D985 (2023).

42. Bulik-Sullivan, B. K. et al. LD Score regression distinguishes confounding from polygenicity in genome-wide association studies. Nat. Genet. 47, 291–295 (2015).

43. Burgess, S., Butterworth, A. & Thompson, S. G. Mendelian randomization analysis with multiple genetic variants using summarized data. Genet. Epidemiol. 37, 658–665 (2013).

44. Bowden, J., Davey Smith, G., Haycock, P. C. & Burgess, S. Consistent estimation in Mendelian randomization with some invalid instruments using a weighted median estimator. Genet. Epidemiol. 40, 304–314 (2016).

45. Hartwig, F. P., Davey Smith, G. & Bowden, J. Robust inference in summary data Mendelian randomization via the zero modal pleiotropy assumption. Int. J. Epidemiol. 46, 1985–1998 (2017).

46. Bowden, J., Davey Smith, G. & Burgess, S. Mendelian randomization with invalid instruments: effect estimation and bias detection through Egger regression. Int. J. Epidemiol. 44, 512–525 (2015).

47. Morrison, J., Knoblauch, N., Marcus, J. H., Stephens, M. & He, X. Mendelian randomization accounting for correlated and uncorrelated pleiotropic effects using genome-wide summary statistics. Nat. Genet. 52, 740–747 (2020).

48. Bédard, S. & Cohen-Adad, J. Automatic measure and normalization of spinal cord cross-sectional area using the pontomedullary junction. Front. Neuroimaging 1, 1031253 (2022).

49. Frostell, A., Hakim, R., Thelin, E. P., Mattsson, P. & Svensson, M. A review of the segmental diameter of the healthy human spinal cord. Front. Neurol. 7, 238 (2016).

50. Abdelhak, A. et al. Blood GFAP as an emerging biomarker in brain and spinal cord disorders. Nat. Rev. Neurol. 18, 158–172 (2022).

51. Vuong, T. A. et al. SGTb regulates a surface localization of a guidance receptor BOC to promote neurite outgrowth. Cell. Signal. 55, 100–108 (2019).

52. Traylor, M., Malik, R., Gesierich, B. & Dichgans, M. The BS variant of C4 protects against age-related loss of white matter microstructural integrity. Brain 145, 295–304 (2022).

53. Junier, M. P. What role(s) for TGFalpha in the central nervous system? Prog. Neurobiol. 62, 443–473 (2000).

54. Karki, P., Johnson, J., Jr, Son, D.-S., Aschner, M. & Lee, E. Transcriptional regulation of human transforming growth factor-α in astrocytes. Mol. Neurobiol. 54, 964–976 (2017).

55. Boillée, S., Cadusseau, J., Coulpier, M., Grannec, G. & Junier, M. P. Transforming growth factor alpha: a promoter of motoneuron survival of potential biological relevance. J. Neurosci. 21, 7079–7088 (2001).

56. Afzal, A. R. & Jeffery, S. One gene, two phenotypes: ROR2 mutations in autosomal recessive Robinow syndrome and autosomal dominant brachydactyly type B. Hum. Mutat. 22, 1–11 (2003).

57. Sastre-Garriga, J. et al. MAGNIMS consensus recommendations on the use of brain and spinal cord atrophy measures in clinical practice. Nat. Rev. Neurol. 16, 171–182 (2020).

58. Keegan, B. M. et al. Spinal cord evaluation in multiple sclerosis: clinical and radiological associations, present and future. Brain Commun. 6, fcae395 (2024).

59. Casserly, C. et al. Spinal cord atrophy in multiple sclerosis: A systematic review and meta-analysis. J. Neuroimaging 28, 556–586 (2018).

60. Eaton, S. E. et al. Spinal-cord involvement in diabetic peripheral neuropathy. Lancet 358, 35–36 (2001).

61. Al-Nasser, B. Early involvement of spinal cord in diabetic peripheral neuropathy may influence patient outcome after neuraxial anesthesia. Journal of anesthesia vol. 26 951–952 (2012).

62. Feldman, E. L. et al. Diabetic neuropathy. Nat. Rev. Dis. Primers 5, 41 (2019).

63. Ludlow, M. et al. Neurotropic virus infections as the cause of immediate and delayed neuropathology. Acta Neuropathol. 131, 159–184 (2016).

64. Verma, A. K. & Perlman, S. Unraveling the complexities of neurotropic virus infection and immune evasion. Microbiol. Mol. Biol. Rev. e0018523 (2025) doi:10.1128/mmbr.00185-23.

65. García-Marín, L. M. et al. Shared molecular genetic factors influence subcortical brain morphometry and Parkinson’s disease risk. NPJ Parkinsons Dis. 9, 73 (2023).

66. Shang, X. et al. Association of a wide range of individual chronic diseases and their multimorbidity with brain volumes in the UK Biobank: A cross-sectional study. EClinicalMedicine 47, 101413 (2022).

67. Accelerated MRI-Predicted Brain Ageing and Its Associations with Cardiometabolic and Brain Disorders. Scientific Reports (2020).

68. Jacobs, B. M., Watson, C., Marshall, C., Noyce, A. & Dobson, R. No evidence for association between polygenic risk of multiple sclerosis and MRI phenotypes in ∼30,000 healthy adult UK Biobank participants. Multiple sclerosis (Houndmills, Basingstoke, England) vol. 28 1656–1657 (2022).

69. Alfaro-Almagro, F. et al. Image processing and Quality Control for the first 10,000 brain imaging datasets from UK Biobank. Neuroimage 166, 400–424 (2018).

70. De Leener, B. et al. PAM50: Unbiased multimodal template of the brainstem and spinal cord aligned with the ICBM152 space. Neuroimage 165, 170–179 (2018).

71. Bycroft, C. et al. The UK Biobank resource with deep phenotyping and genomic data. Nature 562, 203–209 (2018).

72. Taliun, D. et al. Sequencing of 53,831 diverse genomes from the NHLBI TOPMed Program. Nature 590, 290–299 (2021).

73. Chang, C. C. et al. Second-generation PLINK: rising to the challenge of larger and richer datasets. Gigascience 4, 7 (2015).

74. Yang, J., Lee, S. H., Goddard, M. E. & Visscher, P. M. GCTA: a tool for genome-wide complex trait analysis. Am. J. Hum. Genet. 88, 76–82 (2011).

75. Zhou, H. et al. FAVOR: functional annotation of variants online resource and annotator for variation across the human genome. Nucleic Acids Res. 51, D1300–D1311 (2023).

76. Frankish, A. et al. GENCODE: reference annotation for the human and mouse genomes in 2023. Nucleic Acids Res. 51, D942–D949 (2023).

77. 1000 Genomes Project Consortium, et al. A Global Reference for Human Genetic Variation.

78. International HapMap 3 Consortium et al. Integrating common and rare genetic variation in diverse human populations. Nature 467, 52–58 (2010).

79. MacKinnon, J. G. & White, H. Some heteroskedasticity-consistent covariance matrix estimators with improved finite sample properties. J. Econom. 29, 305–325 (1985).

80. Giambartolomei, C. et al. Bayesian test for colocalisation between pairs of genetic association studies using summary statistics. PLoS Genet. 10, e1004383 (2014).

81. Wallace, C. Eliciting priors and relaxing the single causal variant assumption in colocalisation analyses. PLoS Genet. 16, e1008720 (2020).

82. de Klein, N. et al. Brain expression quantitative trait locus and network analyses reveal downstream effects and putative drivers for brain-related diseases. Nat. Genet. 55, 377–388 (2023).

83. Frei, O. et al. Improved functional mapping of complex trait heritability with GSA-MiXeR implicates biologically specific gene sets. Nat. Genet. 56, 1310–1318 (2024).

84. Burgess, S., Davies, N. M. & Thompson, S. G. Bias due to participant overlap in two-sample Mendelian randomization. Genet. Epidemiol. 40, 597–608 (2016).

85. Minelli, C. et al. The use of two-sample methods for Mendelian randomization analyses on single large datasets. Int. J. Epidemiol. 50, 1651–1659 (2021).

86. Hemani, G. et al. The MR-Base platform supports systematic causal inference across the human phenome. Elife 7, (2018).

87. Hemani, G., Tilling, K. & Davey Smith, G. Correction: Orienting the causal relationship between imprecisely measured traits using GWAS summary data. PLoS Genet. 13, e1007149 (2017).

