## Supplementary information for "The genetic architecture of MRI derived human cervical spinal cord morphology reveals sensory-motor axis and biomarkers of neurological and systemic diseases"

Supplementary Materials

Section 1: Descriptive summary of spinal cord phenotypes	1-2

Supplementary figure 1-2: Phenotype summary	1-2

Section 2: Phenotyping robustness and reproducibility evaluation	3-4

Supplementary figure 3 Repeatability evaluation	3

Supplementary figure 4 Evaluation of method robustness	4

Section 3: Genome-wide association (GWA) results in the European ancestry	5-10

Supplementary figure 5-8: Manhattan and Q-Q plots	5-8

Supplementary figure 9: Loci summary for 12 spinal cord metrics	9

Supplementary figure 10: MAF vs. effect size for significant variants	10

Section 4: GWAS covariate selection	11

Supplementary figure 11 Summary of GWAS covariates selection	11

Section 5: GWA results (Excluded height as a covariate)	12-15

Supplementary figure 12-15: Manhattan and Q-Q plots (without correcting for height)	12-15

Section 6: Sex-specific GWA results	16-21

Supplementary figure 16-19: Miami plots	16-19

Supplementary figure 20: Sex-specific SNP heritability correlation	20

Supplementary figure 21: Significant SNP-by-sex interaction effects	21

Section 7: GWA results in non-European ancestries	22-24

Supplementary figure 22-24: Effect size comparison: European vs. non-European cohorts	22-24

Section 8: Gene-based findings	25

Supplementary figure 25: Gene-based findings	25

Section 9: Correlations between spinal cord phenotypes and brain structures	26

Supplementary figure 26: Correlations between spinal cord shape metrics and brain volumes	26

Section 10: Mendelian randomization	27

Supplementary figure 27: Mendelian randomization	27

Section 11: Spinal cord image quality control (QC)	28-31

Supplementary figure 28: Image QC overview	28

Supplementary figure 29-30: Manual QC on vertebral labels	29-30

Supplementary figure 31: Additional removal of noise between processing steps	31

References	32

Section 1: Descriptive summary of spinal cord phenotypes

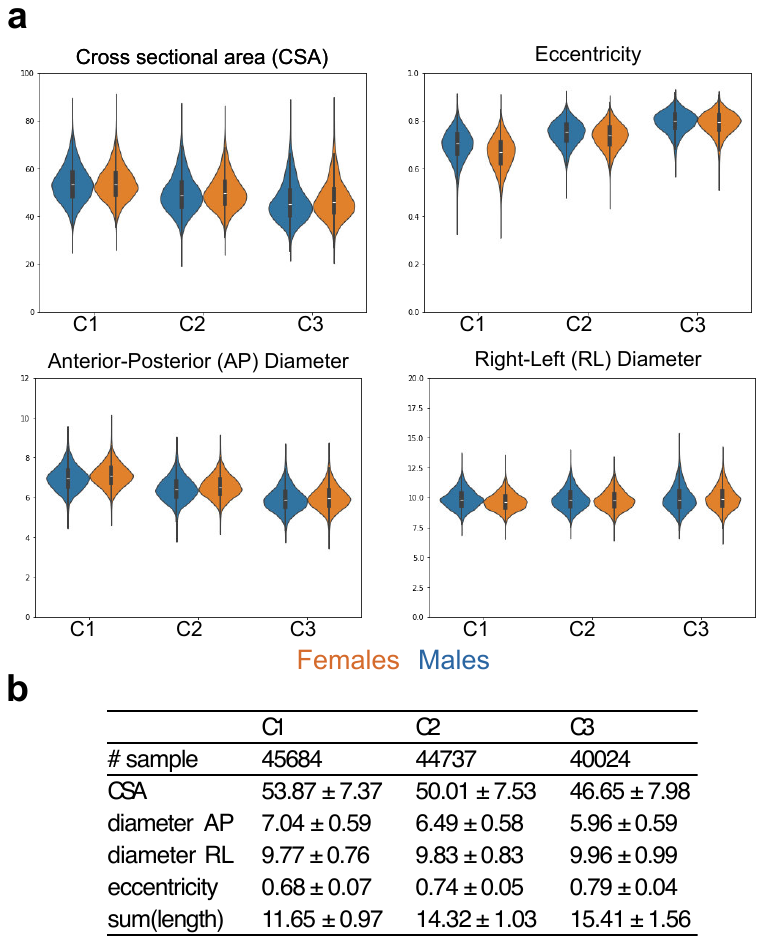

Supplementary Figure 1**: (a)** Violin plots of spinal cord phenotypes after imaging QC at each vertebral level, stratified by sexes (orange: female; blue: male). (**b**) The corresponding sample size, mean and standard deviation of each spinal cord phenotype across C1-C3 in units of mm for diameters and mm^2^ for cross-sectional area (CSA).

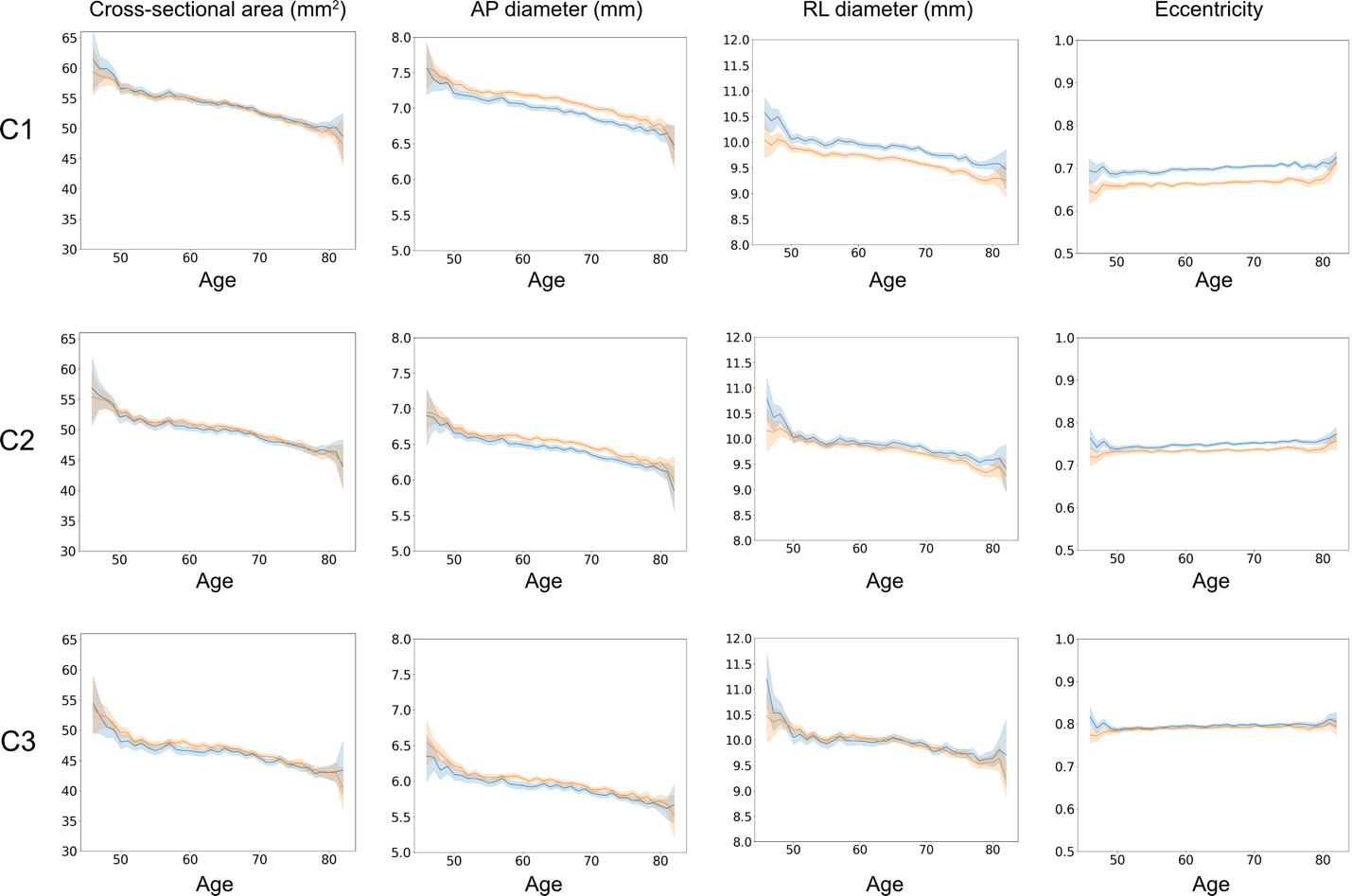

Supplementary Figure 2**:** Line plot showing the mean spinal cord phenotype across age stratified by sex (orange: female; blue: male). The x-axis represents age (years), and the y-axis indicates the average value of the phenotype. Shaded regions denote the 95% confidence interval (CI) of the mean estimate at each age from 45 to 83. AP, anterior-posterior; RL, right –left.

### Section 2: Phenotyping robustness and reproducibility evaluation

**2.1 Repeatability evaluation**

To ensure cross-scan consistency, we conducted a repeatability evaluation using data from participants who underwent repeat imaging. Only data from the first scan session was used in the primary GWAS analysis. A small subset of participants underwent a second scan 1-7 years after their initial session, with most repeat scans were acquired 2-3 years after the initial scan.

Following the same QC procedure, we obtained 4374, 4270, and 3290 spinal cord IDPs for C1, C2, and C3 respectively for reproducibility analysis. The average age at the first scan was 61.5 ± 7.5 years, and 64.2 ± 7.3 years at the second scan. Image quality was slightly lower in the repeat scans, resulting in a higher failure rate during QC especially at the C3 level. This is potentially due to age-related imaging artifacts in older participants.

We assessed scan-to-scan consistency using Spearman’s rank correlation coefficient, stratifying results by the time interval between the two scans (x-axis) (Supplementary Figure 3). We observed age-related degeneration in spinal cord cross-sectional area and diameters, as indicated by decreasing correlations with increasing scan intervals. In contrast, eccentricity measurements showed consistently high correlations regardless of the age gap, suggesting greater stability of this phenotype over time.

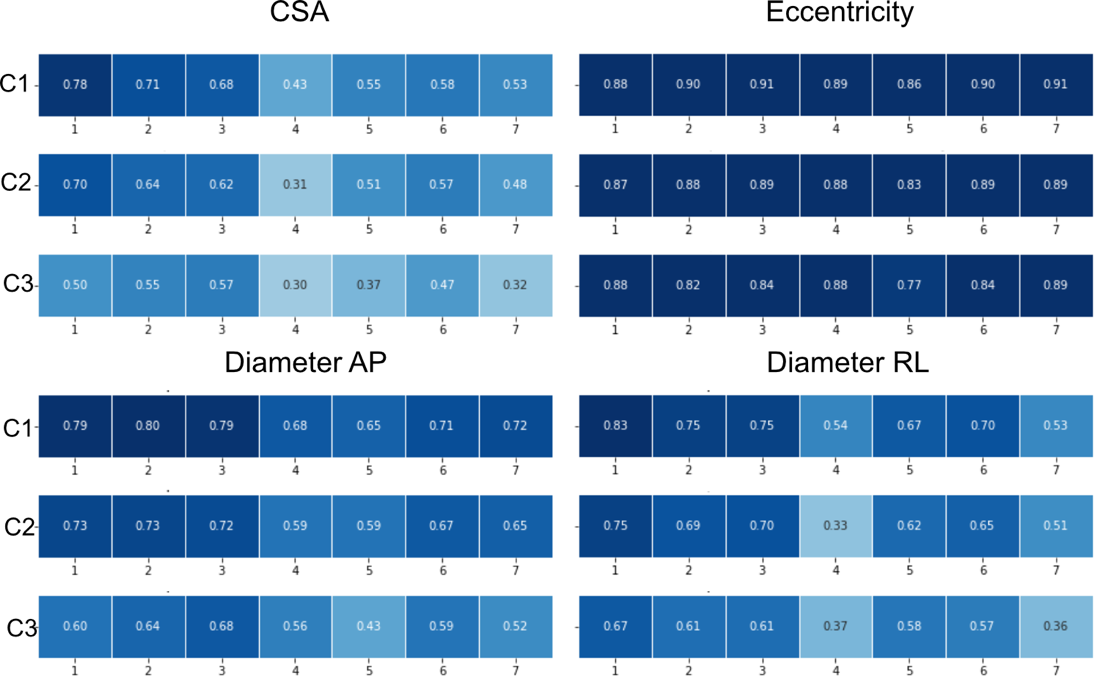

Supplementary Figure 3**.** Heatmaps of the Spearman’s rank correlation coefficients between the first and second scan for each spinal cord phenotype. Correlation values are plotted against the between scans intervals (years).

**2.2 Evaluation of method robustness: atlas-based vs. native-space analysis**

To assess the robustness of spinal cord metrics, we compared two processing pipelines: native-space and atlas-based analysis. While the current study used the native-space approach, the atlas-based method was evaluated as an alternative to ensure the spinal cord shape metrics can be reliably extracted across processing pipelines. Both approaches began with segmentation and vertebral labelling of C1–C3. In the native-space method, metrics were extracted directly from the labelled segmentation (seg_labeled.nii). In contrast, the atlas-based pipeline included an additional registration step to using the PAM50 atlas^2^, with metrics derived from the warped atlas labels (PAM50_levels.nii.gz). For implementation details and exact code, please refer to <https://github.com/art2mri/Enigma-SC>.

We focused on C2 and C3 phenotypes for method comparison because the native-space analysis made substantial improvements to C1 labelling, make the atlas-based method an inadequate comparator. Besides the differences in C1 labelling and registration errors, we found strong consistency between native-space and atlas-based methods at the C2 and C3 levels (Supplementary Figure 4). Across eight phenotypes, including cross-sectional area, anterior-posterior and right-left diameters, and eccentricity, the Spearman’s correlation coefficients between the two methods yielded R² values ranging from 0.97 to 0.99. These results demonstrate excellent robustness of the derived spinal cord metrics at the C2 and C3 levels.

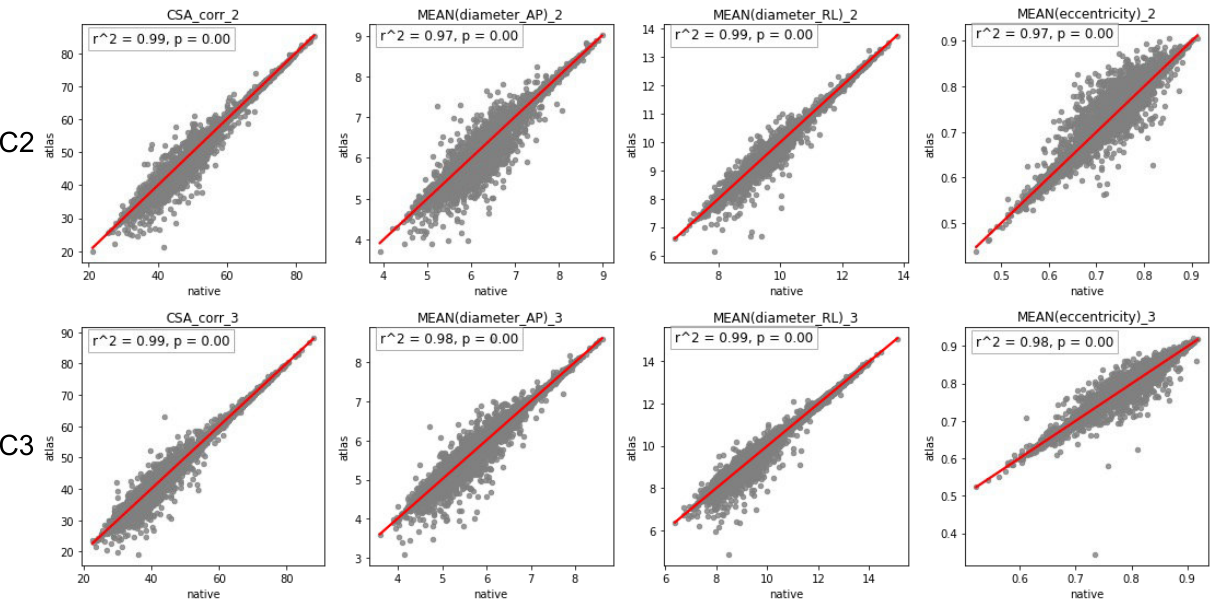

Supplementary Figure 4. Comparison of spinal cord metrics at C2 and C3 between native-space and atlas-based analyses. Spearman’s correlations are shown for cross-sectional area, anterior-posterior diameter, right-left diameter, and eccentricity. Each scatter plot displays the R² and *p*-value, confirming high agreement (R² = 0.97–0.99) across all eight phenotypes.

### Section 3: Genome-wide association (GWA) results in the European ancestry

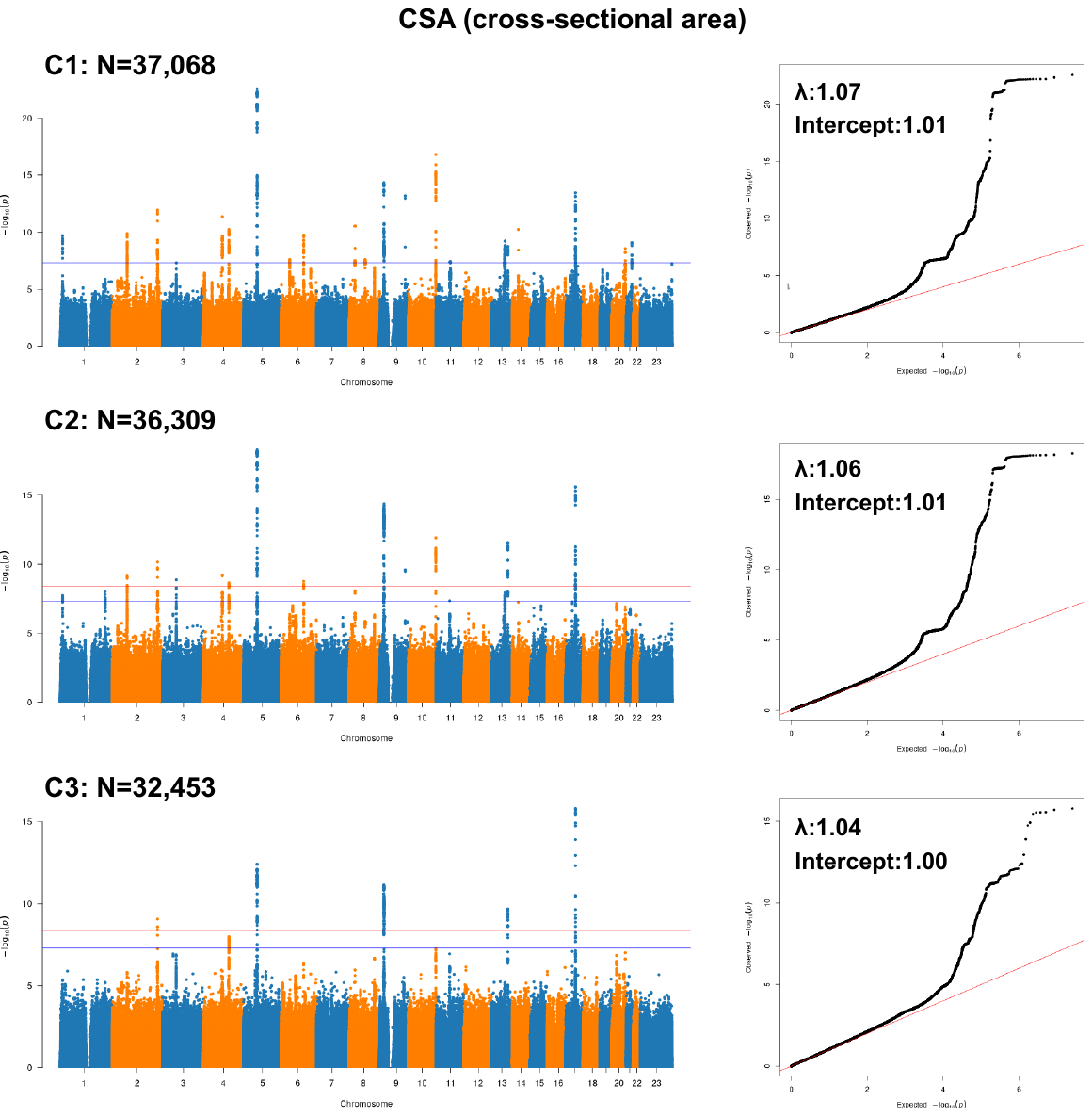

Supplementary Figure 5**.** Manhattan and quantile–quantile (Q–Q) plots for cross-sectional area at the C1, C2, and C3 vertebral levels in the European sample. In Manhattan plots, each point represents a single genetic variant according to its genomic position (x-axis) and −log_10_P (two-sided P values). The blue horizontal line indicates the conventional genome-wide significance threshold (P < 5 × 10⁻⁸), while the red line marks the Bonferroni-corrected threshold used in this study (P < 4.2 × 10⁻⁹), accounting for 12 phenotypes. In the Q–Q plots, the red diagonal line represents the expected null distribution. Genomic inflation factors (λ) and LD Score regression intercepts are labelled within each plot.

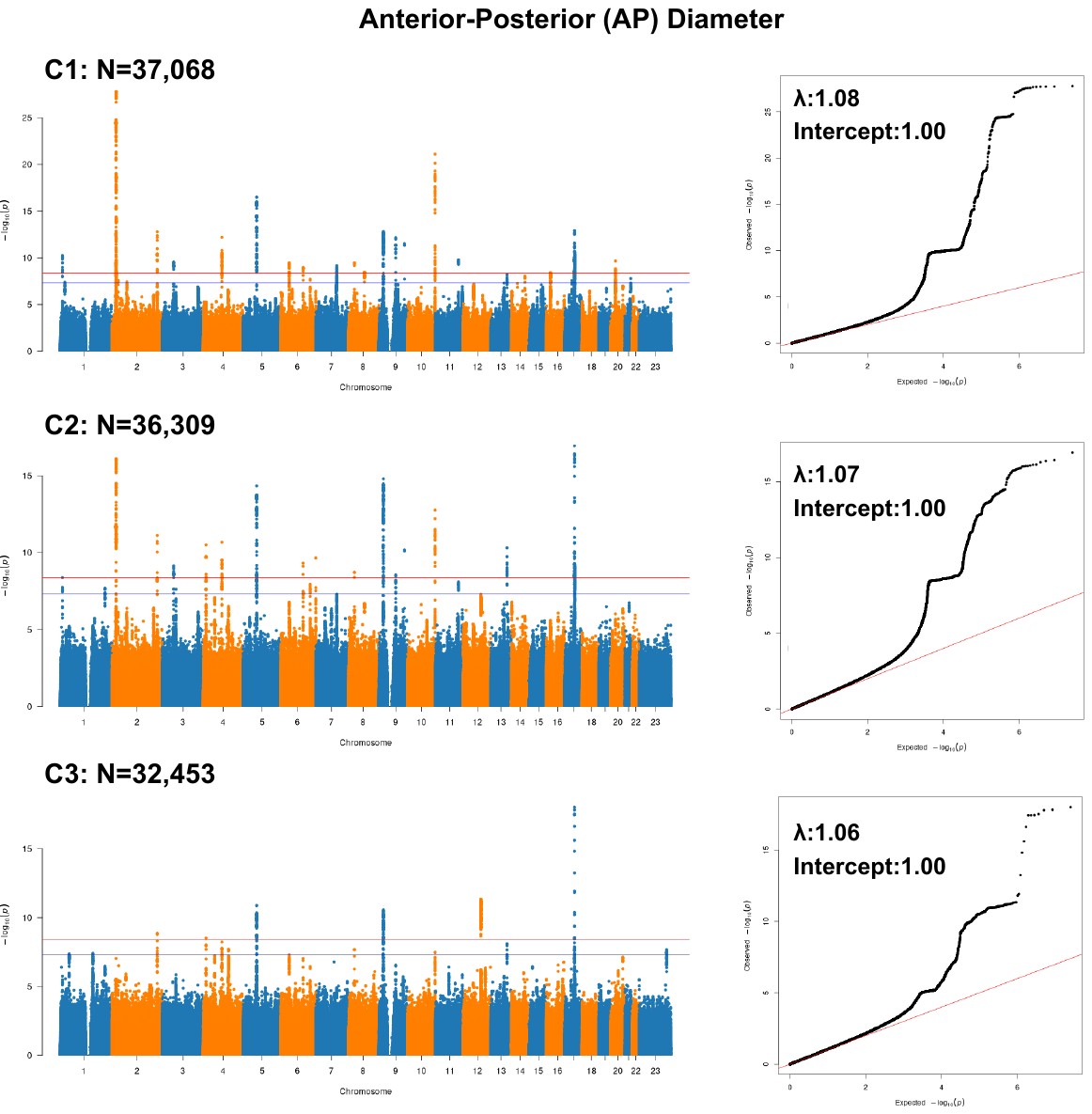

Supplementary Figure 6**.** Manhattan and quantile–quantile (Q–Q) plots for anterior-posterior diameter at the C1, C2, and C3 vertebral levels in the European sample. In Manhattan plots, each point represents a single genetic variant according to its genomic position (x-axis) and −log_10_P (two-sided P values). The blue horizontal line indicates the conventional genome-wide significance threshold (P < 5 × 10⁻⁸), while the red line marks the Bonferroni-corrected threshold used in this study (P < 4.2 × 10⁻⁹), accounting for 12 phenotypes. In the Q–Q plots, the red diagonal line represents the expected null distribution. Genomic inflation factors (λ) and LD Score regression intercepts are labelled within each plot.

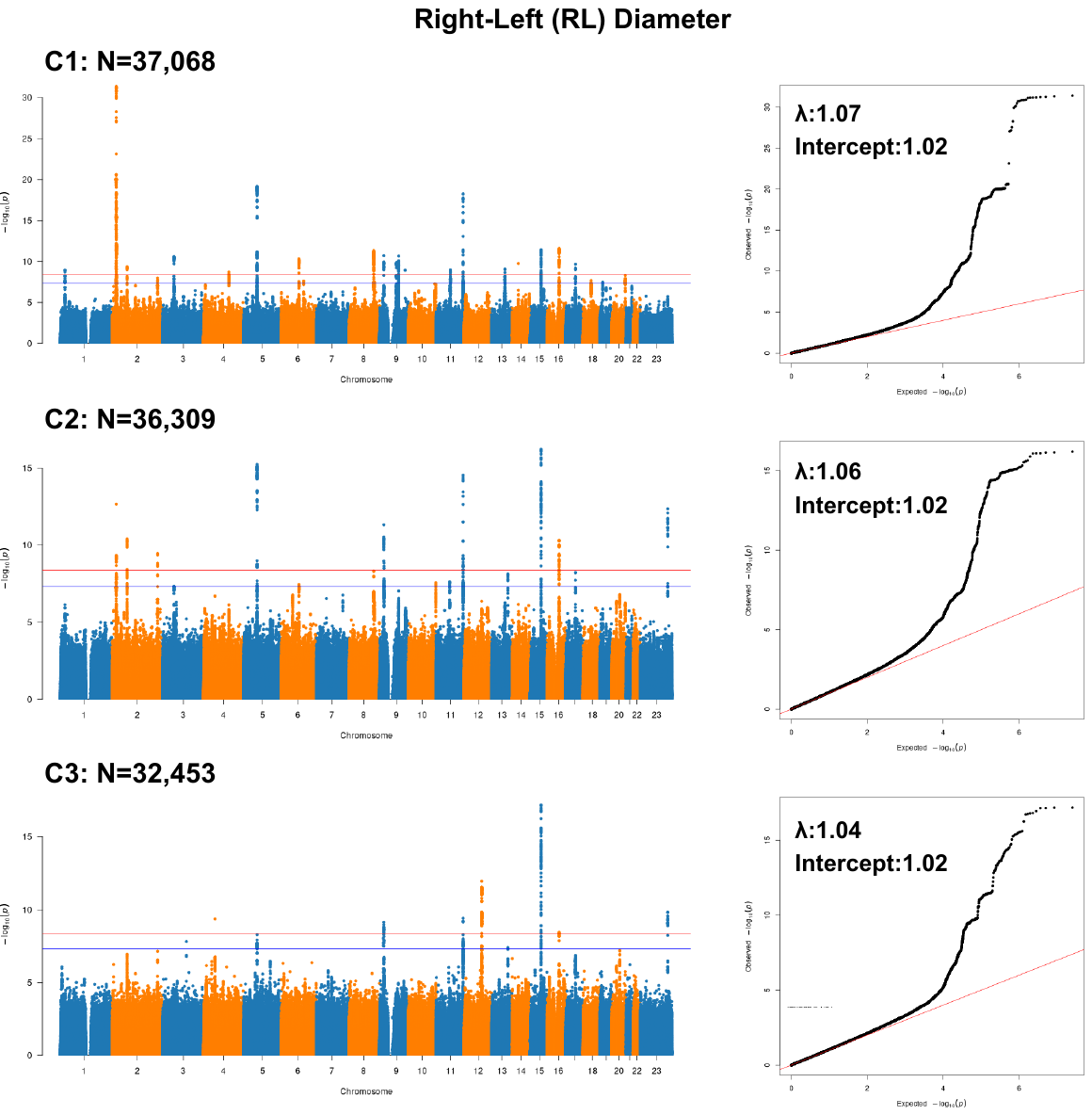

Supplementary Figure 7**.** Manhattan and quantile–quantile (Q–Q) plots for right-left diameter at the C1, C2, and C3 vertebral levels in the European sample. In Manhattan plots, each point represents a single genetic variant according to its genomic position (x-axis) and −log_10_P (two-sided P values). The blue horizontal line indicates the conventional genome-wide significance threshold (P < 5 × 10⁻⁸), while the red line marks the Bonferroni-corrected threshold used in this study (P < 4.2 × 10⁻⁹), accounting for 12 phenotypes. In the Q–Q plots, the red diagonal line represents the expected null distribution. Genomic inflation factors (λ) and LD Score regression intercepts are labelled within each plot.

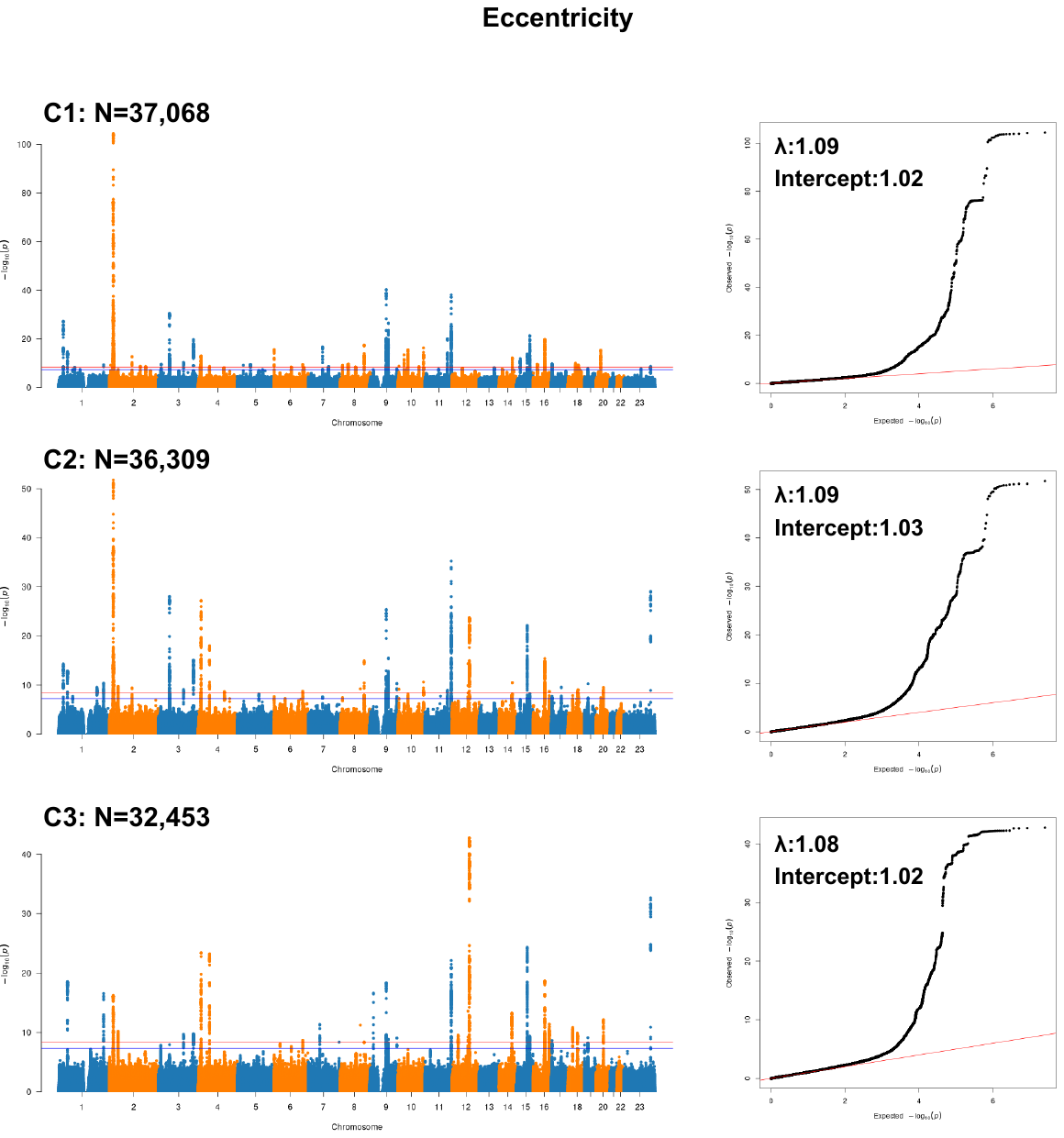

Supplementary Figure 8**.** Manhattan and quantile–quantile (Q–Q) plots for **eccentricity** at the C1, C2, and C3 vertebral levels in the European sample. In Manhattan plots, each point represents a single genetic variant according to its genomic position (x-axis) and −log_10_P (two-sided P values). The blue horizontal line indicates the conventional genome-wide significance threshold (P < 5 × 10⁻⁸), while the red line marks the Bonferroni-corrected threshold used in this study (P < 4.2 × 10⁻⁹), accounting for 12 phenotypes. In the Q–Q plots, the red diagonal line represents the expected null distribution. Genomic inflation factors (λ) and LD Score regression intercepts are labelled within each plot.

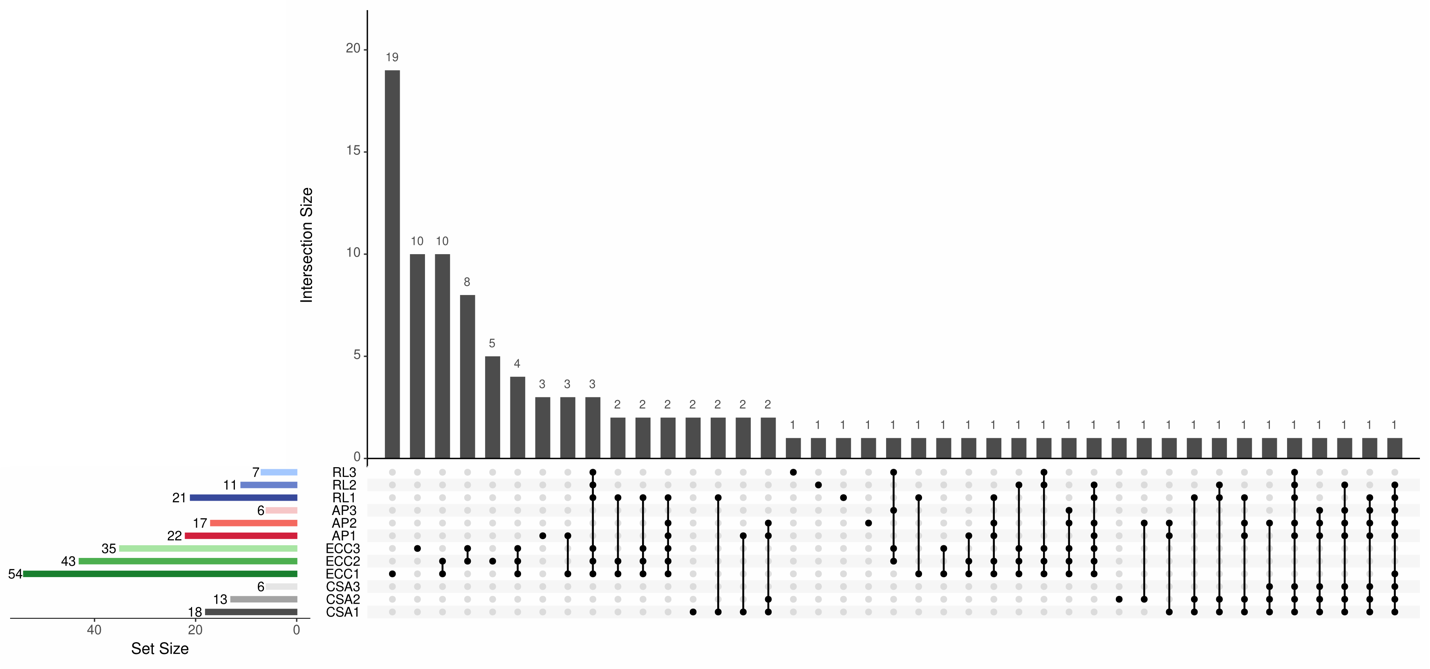

Supplementary Figure 9**.** **Summary of loci identified for 12 spinal cord shape metrics.** Loci were defined by merging independent SNPs with P < 4.17 × 10⁻⁹ that were within 250 kilobases of each other and had pairwise linkage disequilibrium R² > 0.1.

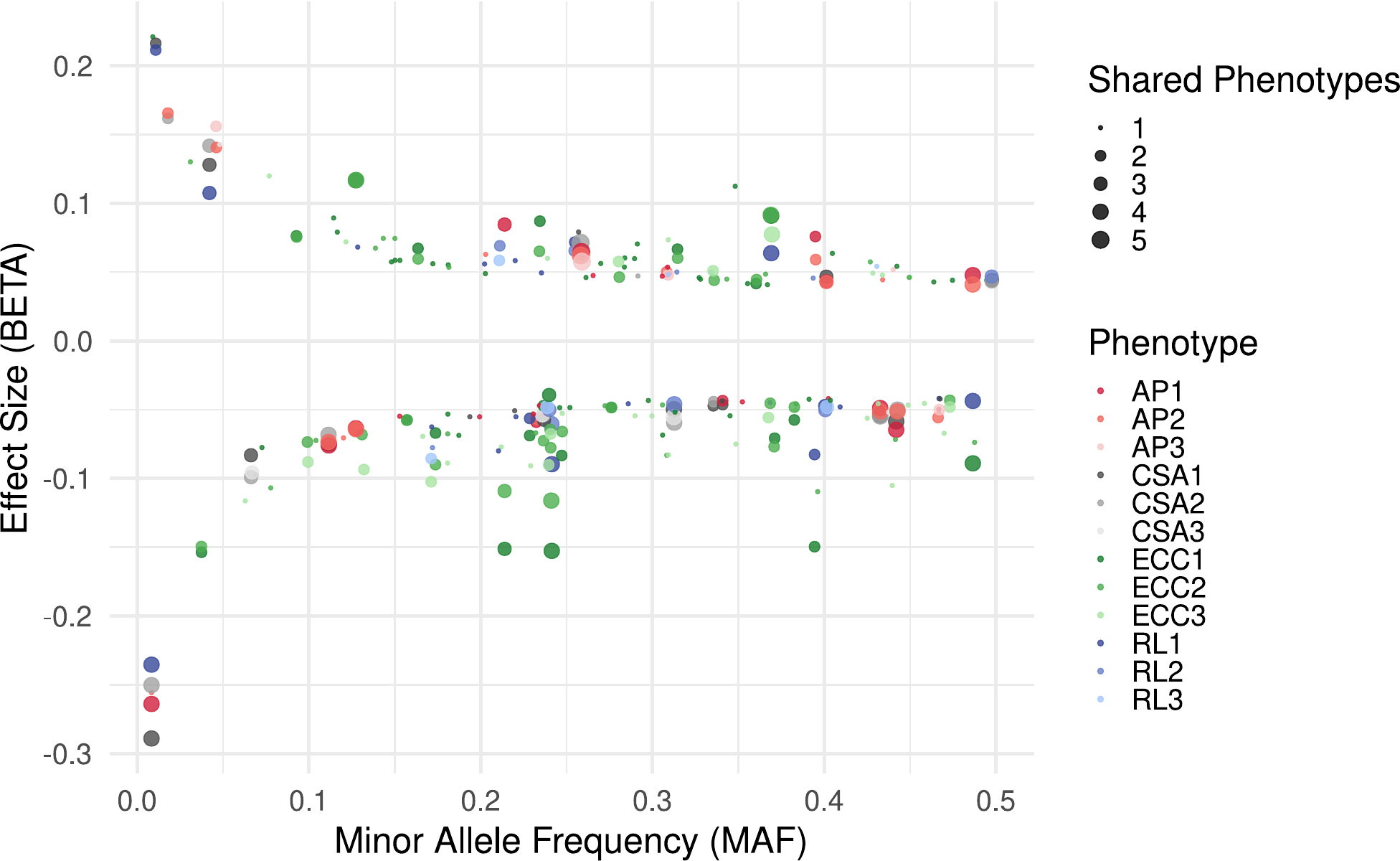

Supplementary Figure 10**. Minor allele frequency (MAF) versus effect size (BETA) for genome-wide significant variants.** Each point represents a variant that reached genome-wide significance (*P* < 4.2 × 10⁻⁹) in the GWAS. Shapes indicate whether SNPs are shared across multiple phenotypes, colors represent the four spinal cord shape metrics, and shading reflects the vertebral level (C1 to C3).

### Section 4: GWAS covariate selection using stepwise regression

To validate the choice of covariates, we performed stepwise regression guided by principal component analysis (PCA). PCA was conducted on the four selected spinal cord phenotypes (cross-sectional area (CSA), eccentricity, anterior-posterior diameter, right-leftdiameter) separately for each vertebral level (C1–C3). Across all levels, the first two principal components (PC1 and PC2) accounted for over 99% of the variance. PC1 was primarily driven by CSA and diameter measures, while PC2 was mainly influenced by eccentricity and diameter measures. We tested sex, age, height, intracranial volume (ICV), imaging site, and T1-weighted signal-to-noise ratio (SNR) as candidate covariates in stepwise regression models and selected the best-fitting model based on the lowest Akaike Information Criterion (AIC). Results indicated that all tested covariates, except for imaging site, were significant contributors. However, imaging site was still included as a routine covariate to account for potential batch effects in GWAS.

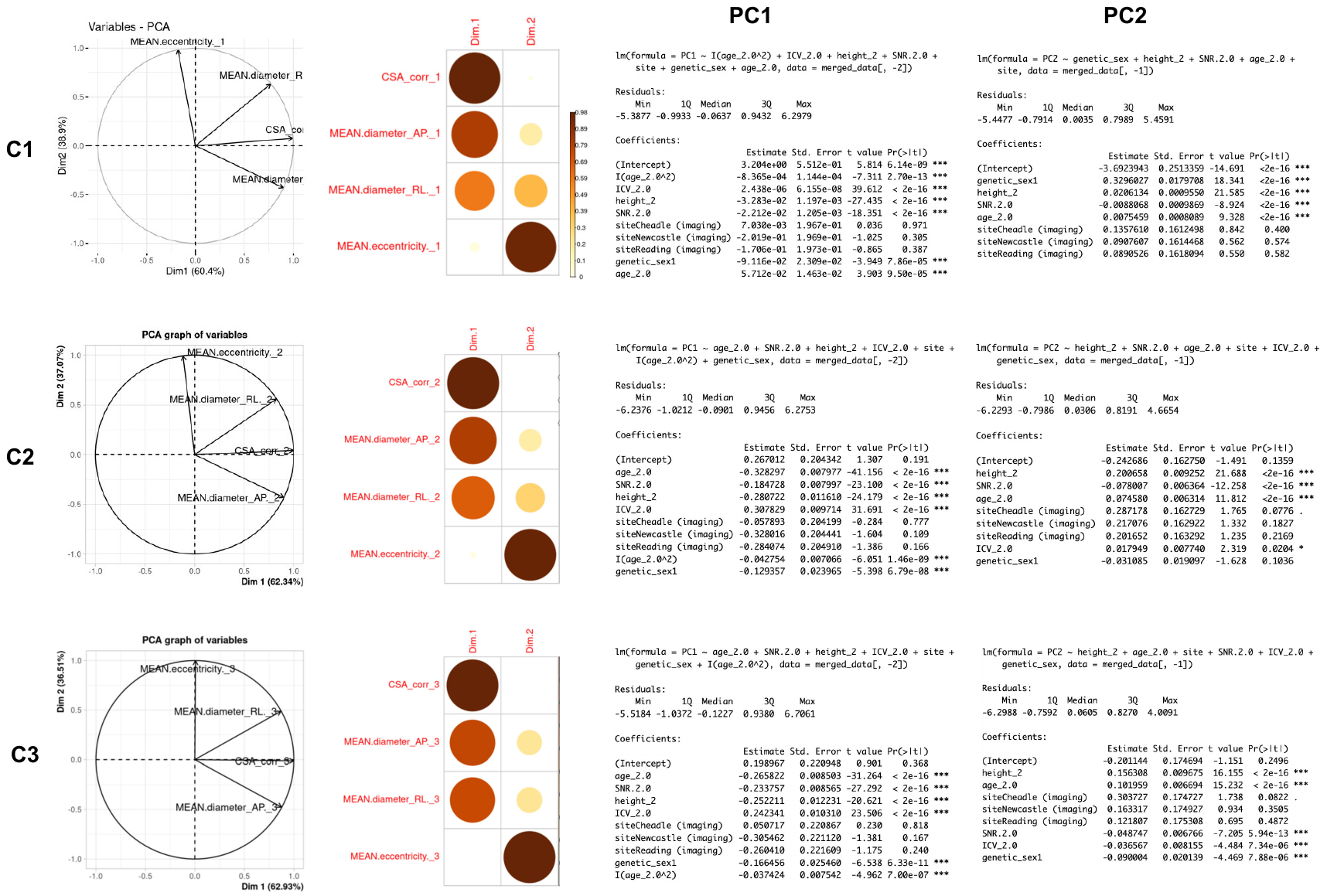

Supplementary Figure 11: Principal component analysis (PCA) of four spinal cord phenotypes (CSA, eccentricity, anterior-posterior diameter, right-left diameter) at each vertebral level (C1–C3). The plots show the correlations of PC1 and PC2 with each phenotype. Summary of covariates selected through stepwise regression based on models with the lowest AIC.

### Section 5: GWA results in the European ancestry (Excluded height as a covariate)

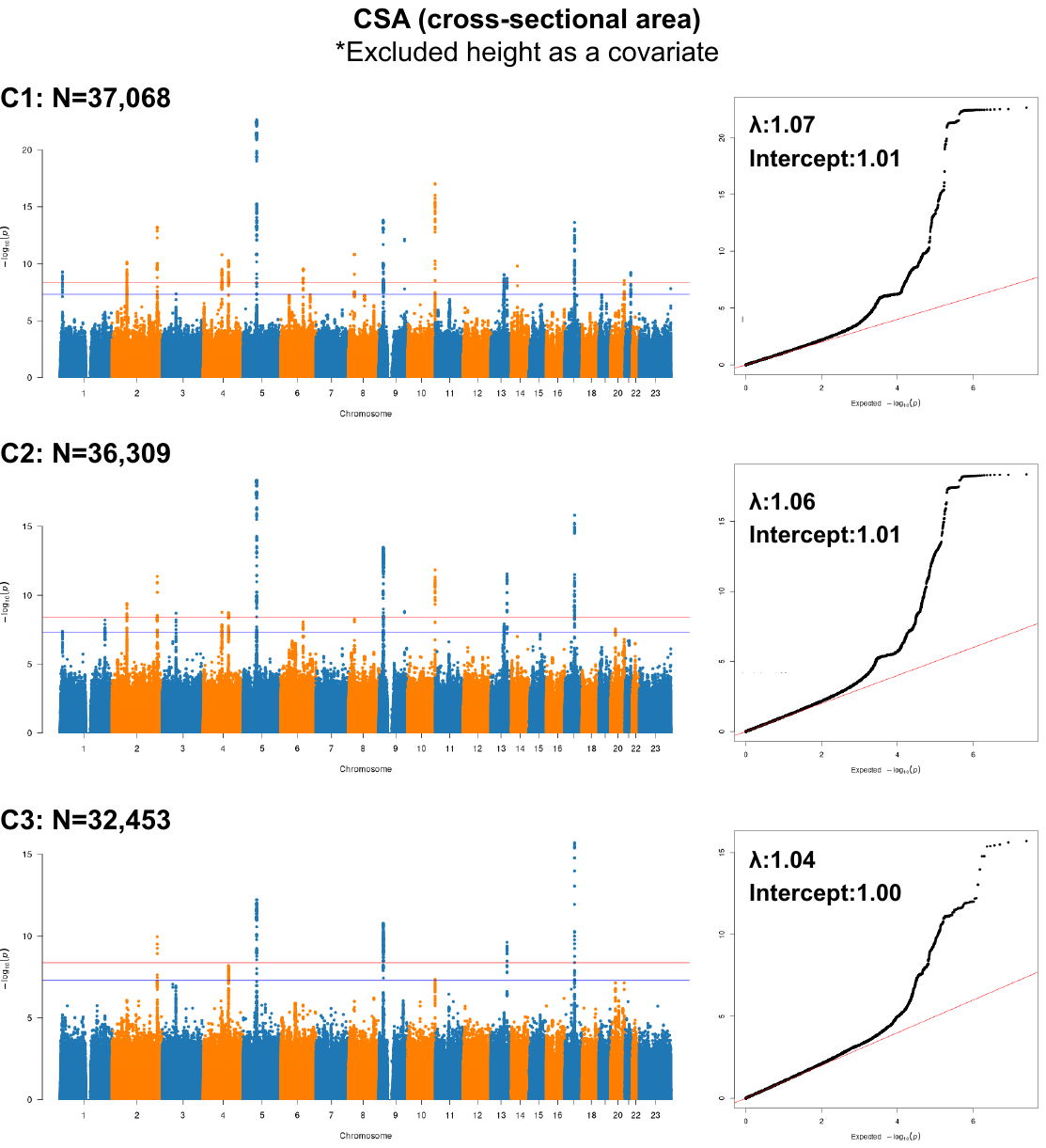

Supplementary Figure 12**.** Manhattan and quantile–quantile (Q–Q) plots for cross-sectional area at the C1, C2, and C3 vertebral levels in the European sample, without height as a covariate. In Manhattan plots, each point represents a single genetic variant according to its genomic position (x-axis) and −log_10_P (two-sided P values). The blue horizontal line indicates the conventional genome-wide significance threshold (P < 5 × 10⁻⁸), while the red line marks the Bonferroni-corrected threshold used in this study (P < 4.2 × 10⁻⁹), accounting for 12 phenotypes. In the Q–Q plots, the red diagonal line represents the expected null distribution. Genomic inflation factors (λ) and LD Score regression intercepts are labelled within each plot.

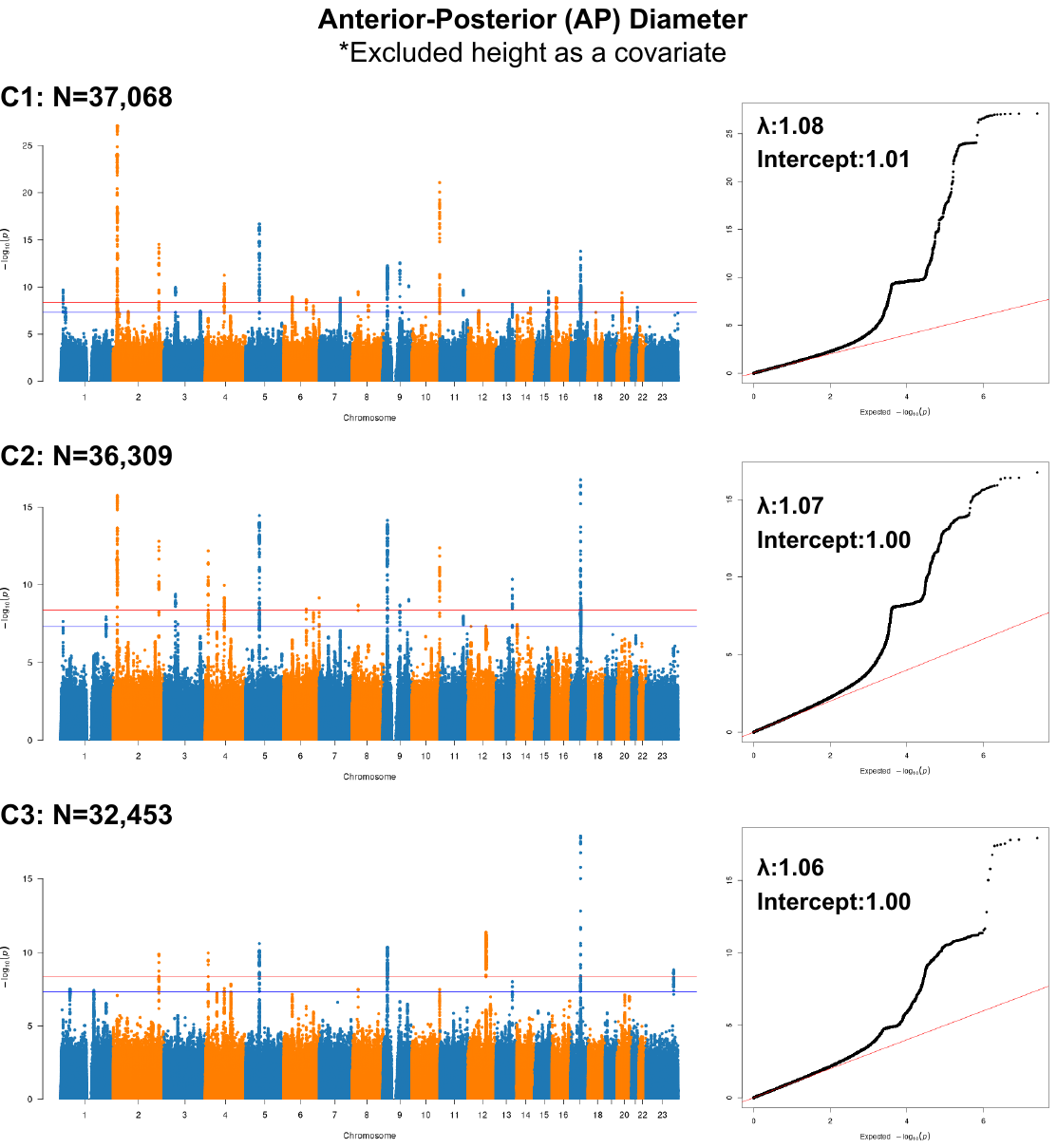

Supplementary Figure 13**.** Manhattan and quantile–quantile (Q–Q) plots for anterior-posterior diameter at the C1, C2, and C3 vertebral levels in the European sample, without height as a covariate. In Manhattan plots, each point represents a single genetic variant according to its genomic position (x-axis) and −log_10_P (two-sided P values). The blue horizontal line indicates the conventional genome-wide significance threshold (P < 5 × 10⁻⁸), while the red line marks the Bonferroni-corrected threshold used in this study (P < 4.2 × 10⁻⁹), accounting for 12 phenotypes. In the Q–Q plots, the red diagonal line represents the expected null distribution. Genomic inflation factors (λ) and LD Score regression intercepts are labelled within each plot.

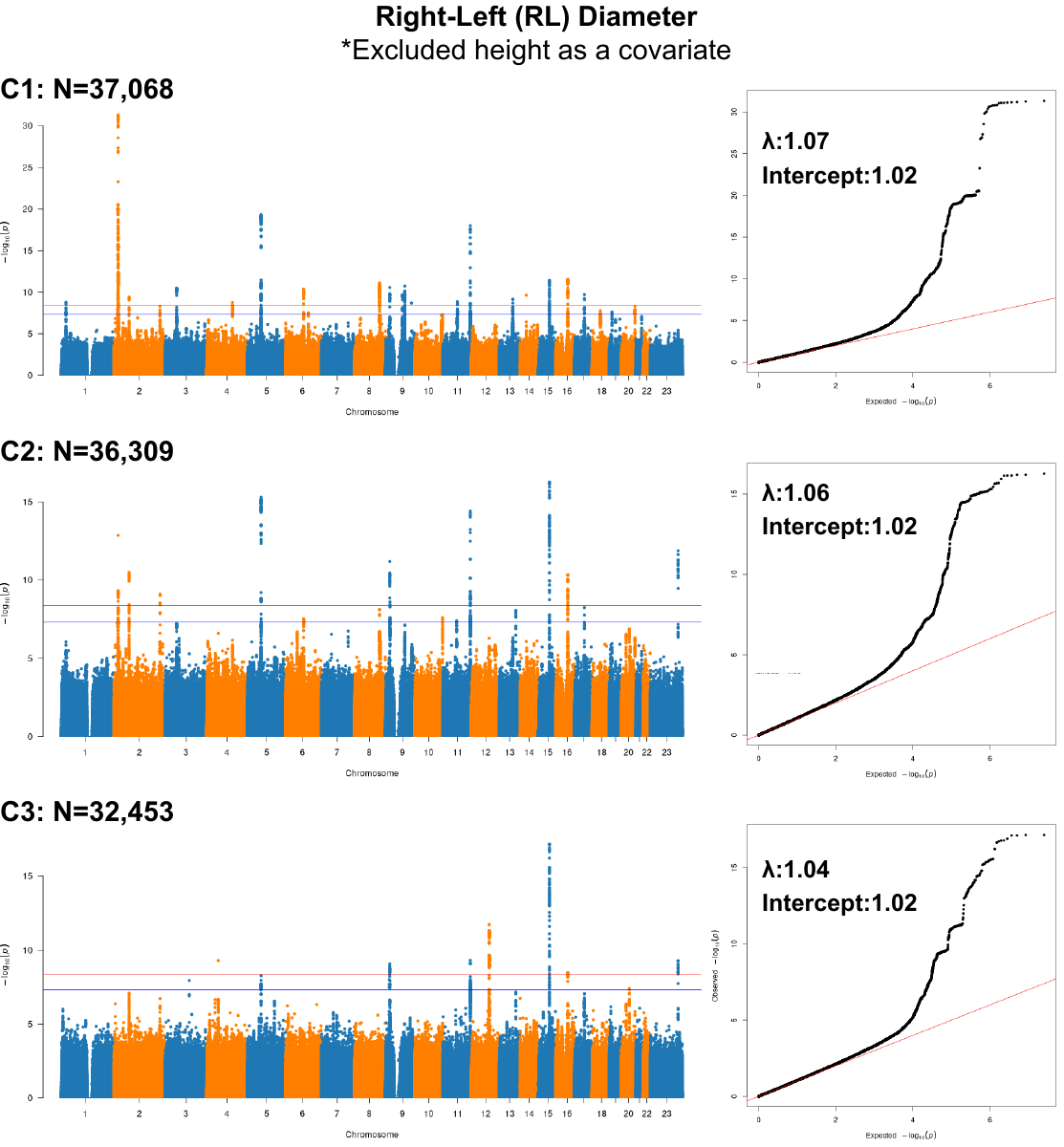

Supplementary Figure 14**.** Manhattan and quantile–quantile (Q–Q) plots for right-left diameter at the C1, C2, and C3 vertebral levels in the European sample, without height as a covariate. In Manhattan plots, each point represents a single genetic variant according to its genomic position (x-axis) and −log_10_P (two-sided P values). The blue horizontal line indicates the conventional genome-wide significance threshold (P < 5 × 10⁻⁸), while the red line marks the Bonferroni-corrected threshold used in this study (P < 4.2 × 10⁻⁹), accounting for 12 phenotypes. In the Q–Q plots, the red diagonal line represents the expected null distribution. Genomic inflation factors (λ) and LD Score regression intercepts are labelled within each plot.

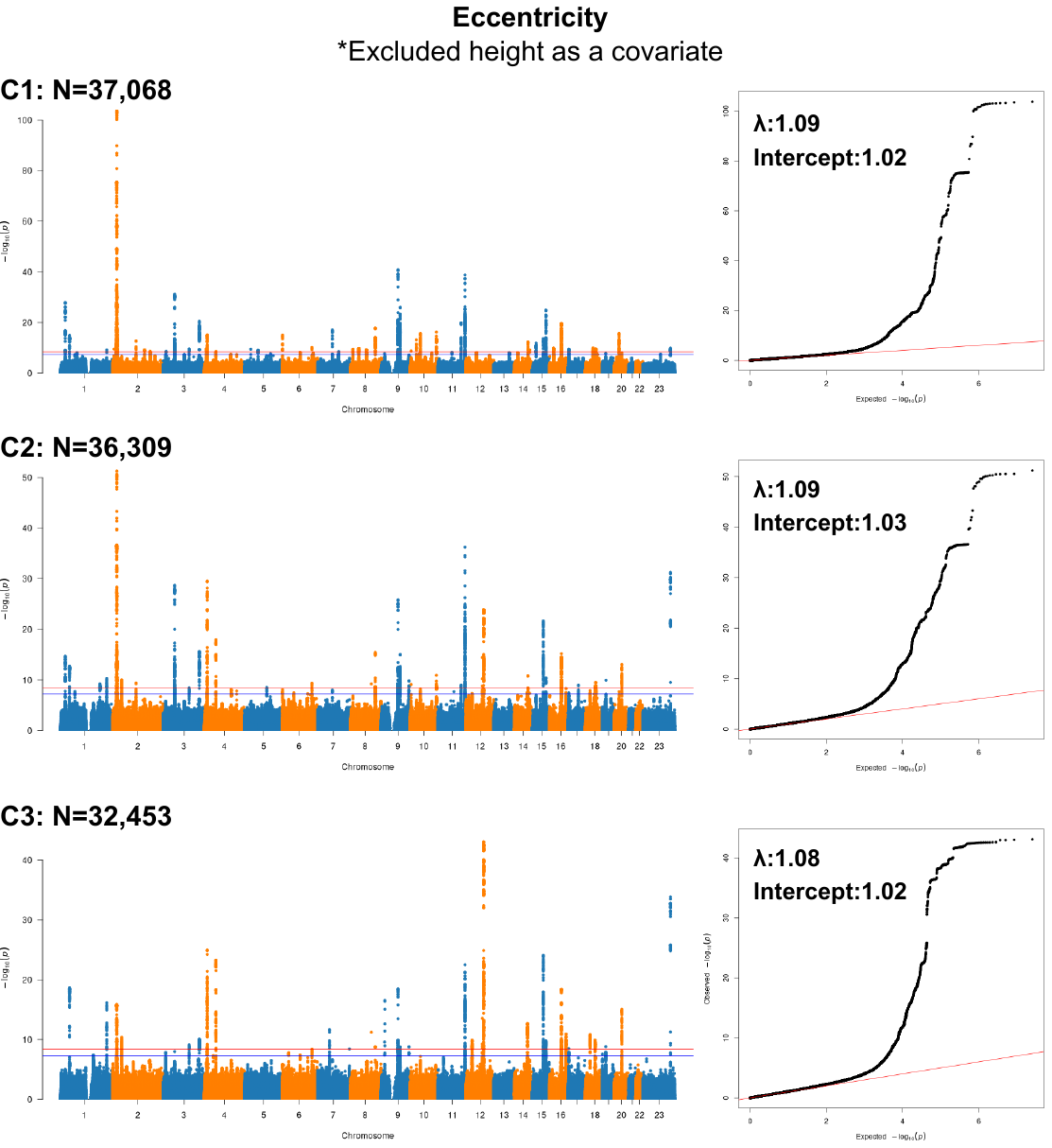

Supplementary Figure 15**.** Manhattan and quantile–quantile (Q–Q) plots for eccentricity at the C1, C2, and C3 vertebral levels in the European sample, without height as a covariate. In Manhattan plots, each point represents a single genetic variant according to its genomic position (x-axis) and −log_10_P (two-sided P values). The blue horizontal line indicates the conventional genome-wide significance threshold (P < 5 × 10⁻⁸), while the red line marks the Bonferroni-corrected threshold used in this study (P < 4.2 × 10⁻⁹), accounting for 12 phenotypes. In the Q–Q plots, the red diagonal line represents the expected null distribution. Genomic inflation factors (λ) and LD Score regression intercepts are labelled within each plot.

**Section 6: Sex-specific GWAS results**

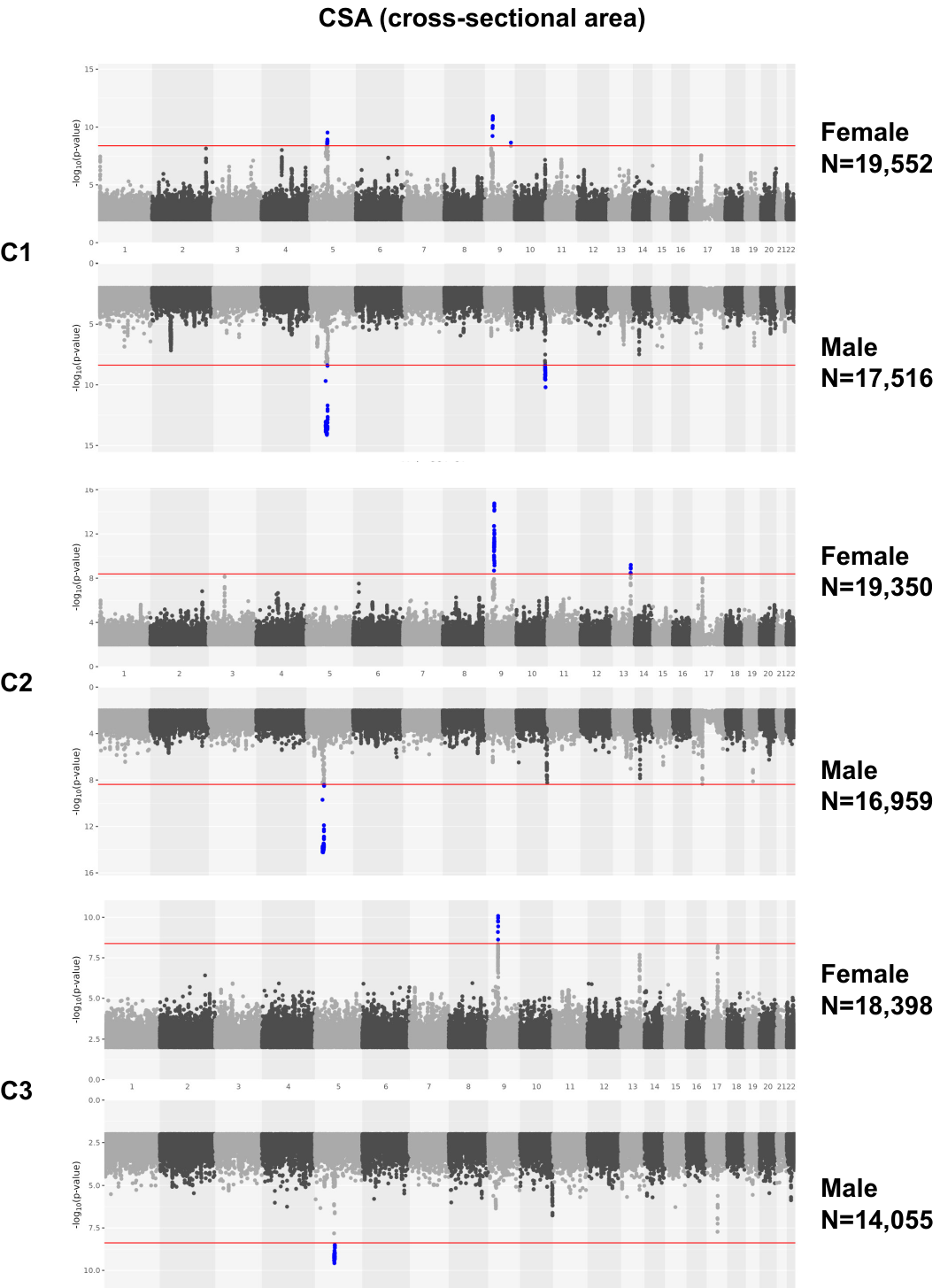

Supplementary Figure 16. Miami plot of sex-stratified GWAS for **cross-sectional area**. Results for females are shown in the upper panel and for males in the lower panel. The genome-wide significance threshold (P < 4.2 × 10⁻⁹) is indicated by the red horizontal line. Genome-wide significant variants are highlighted in blue. SNPs with –log₁₀(P) < 2 are not displayed.

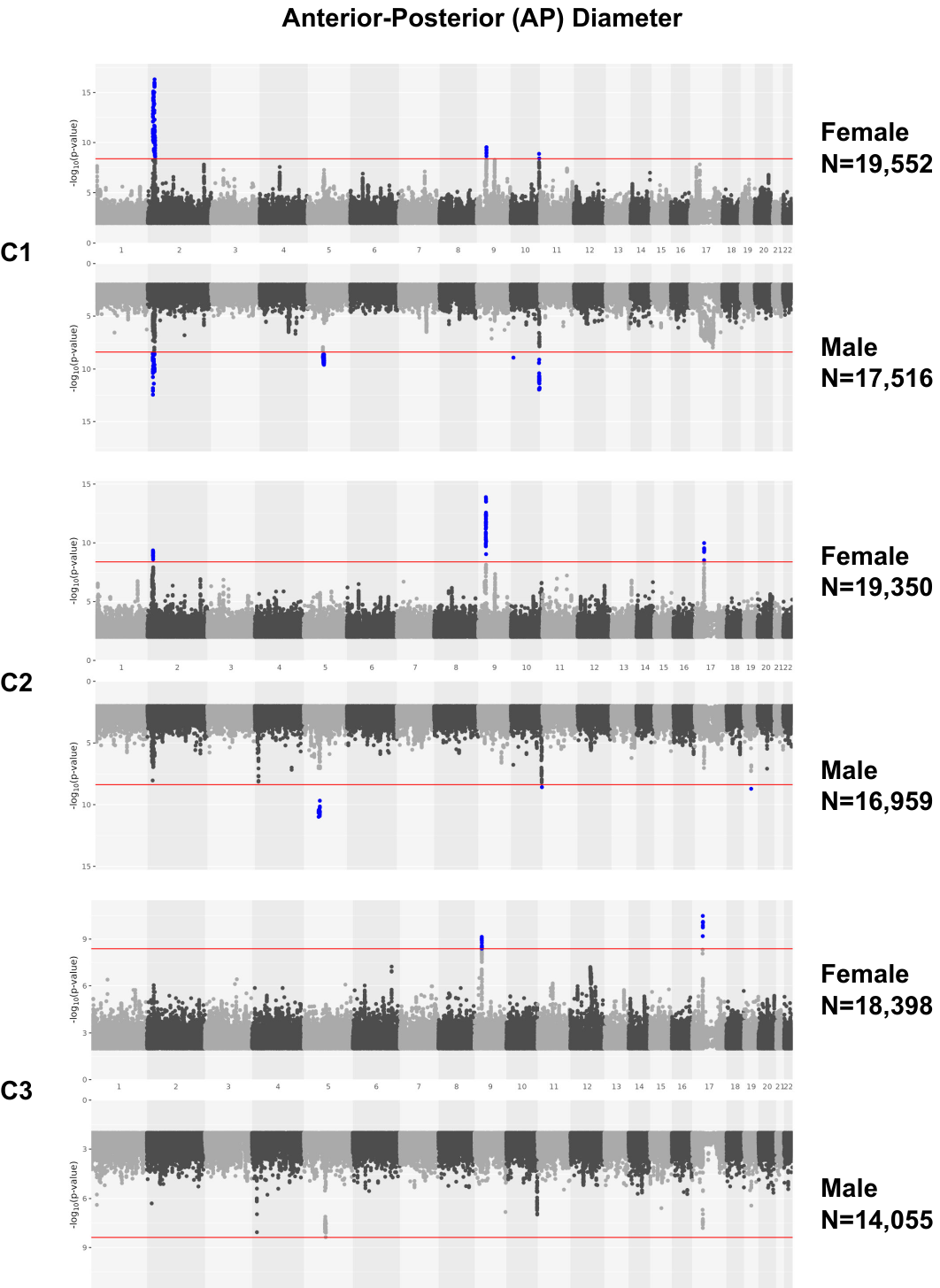

Supplementary Figure 17**.** Miami plot of sex-stratified GWAS for anterior-posterior diameter. Results for females are shown in the upper panel and for males in the lower panel. The genome-wide significance threshold (P < 4.2 × 10⁻⁹) is indicated by the red horizontal line. Genome-wide significant variants are highlighted in blue. SNPs with –log₁₀(P) < 2 are not displayed.

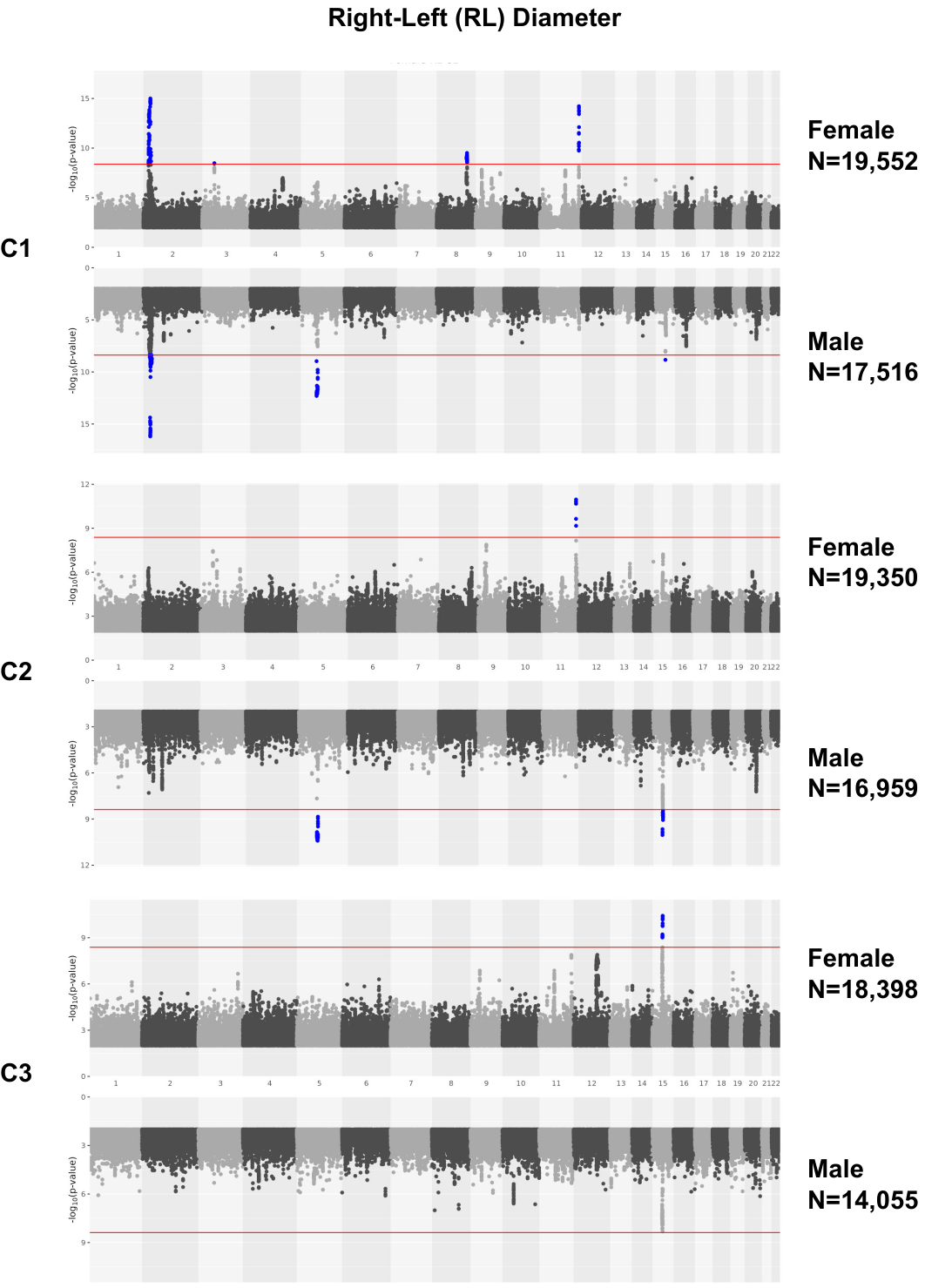

Supplementary Figure 18**.** Miami plot of sex-stratified GWAS for right-left diameter. Results for females are shown in the upper panel and for males in the lower panel. The genome-wide significance threshold (P < 4.2 × 10⁻⁹) is indicated by the red horizontal line. Genome-wide significant variants are highlighted in blue. SNPs with –log₁₀(P) < 2 are not displayed.

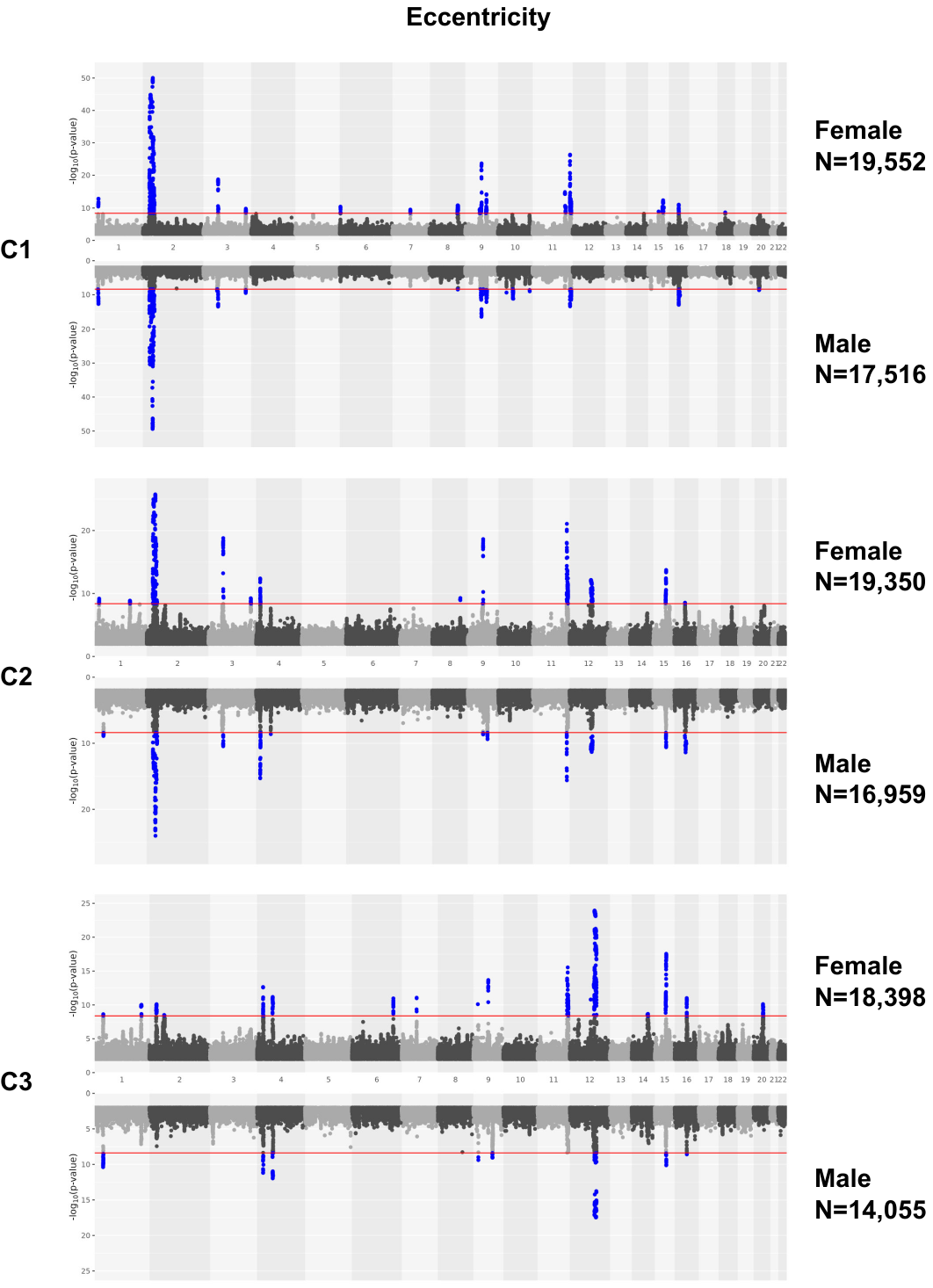

Supplementary Figure 19**.** Miami plot of sex-stratified GWAS for eccentricity. Results for females are shown in the upper panel and for males in the lower panel. The genome-wide significance threshold (P < 4.2 × 10⁻⁹) is indicated by the red horizontal line. Genome-wide significant variants are highlighted in blue. SNPs with –log₁₀(P) < 2 are not displayed.

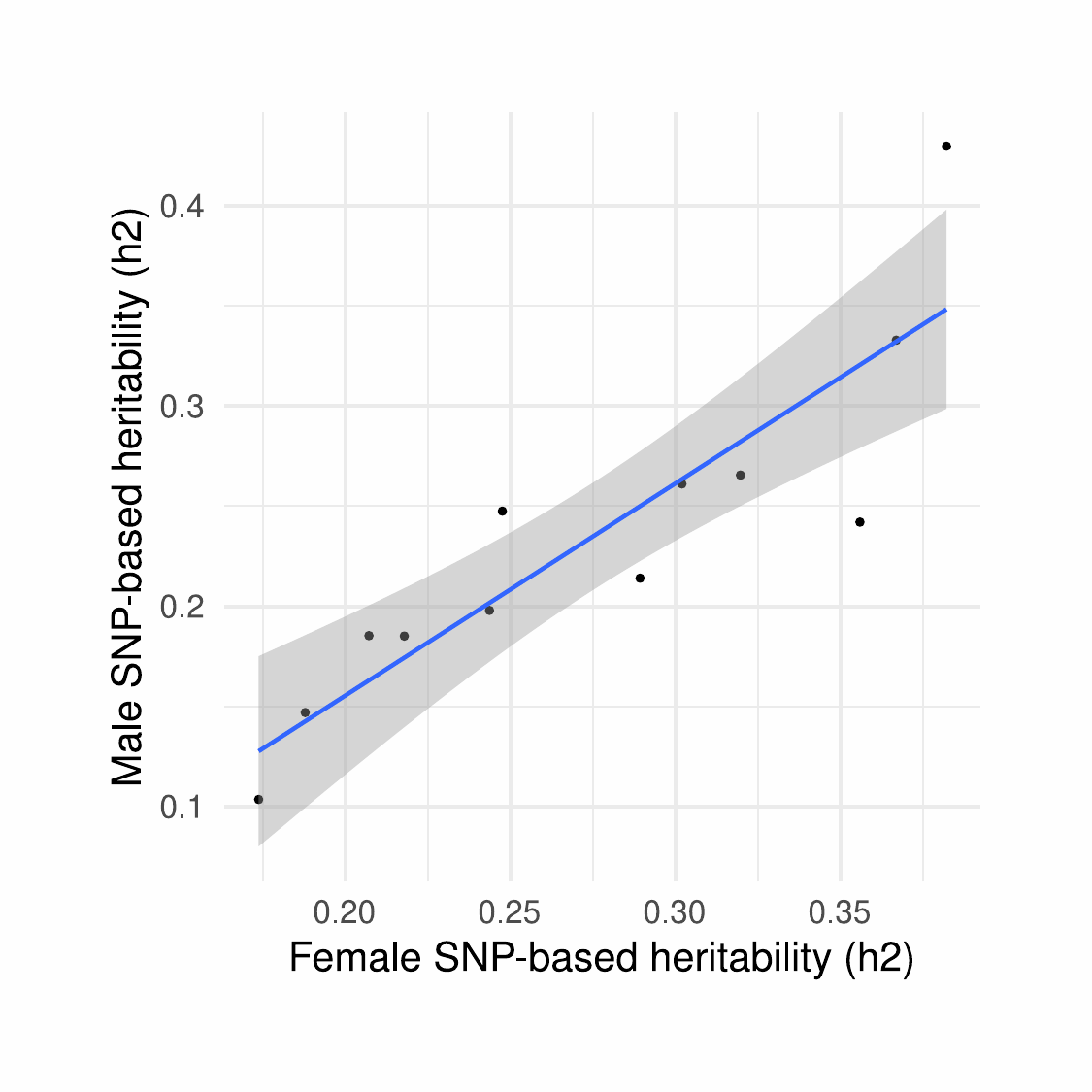

Supplementary Figure 20**. Correlation of sex-specific SNP-based heritability estimates.** The scatter plot shows the correlation between male- and female-specific SNP-based heritability estimates across 12 spinal cord shape metrics. Each point represents one phenotype.

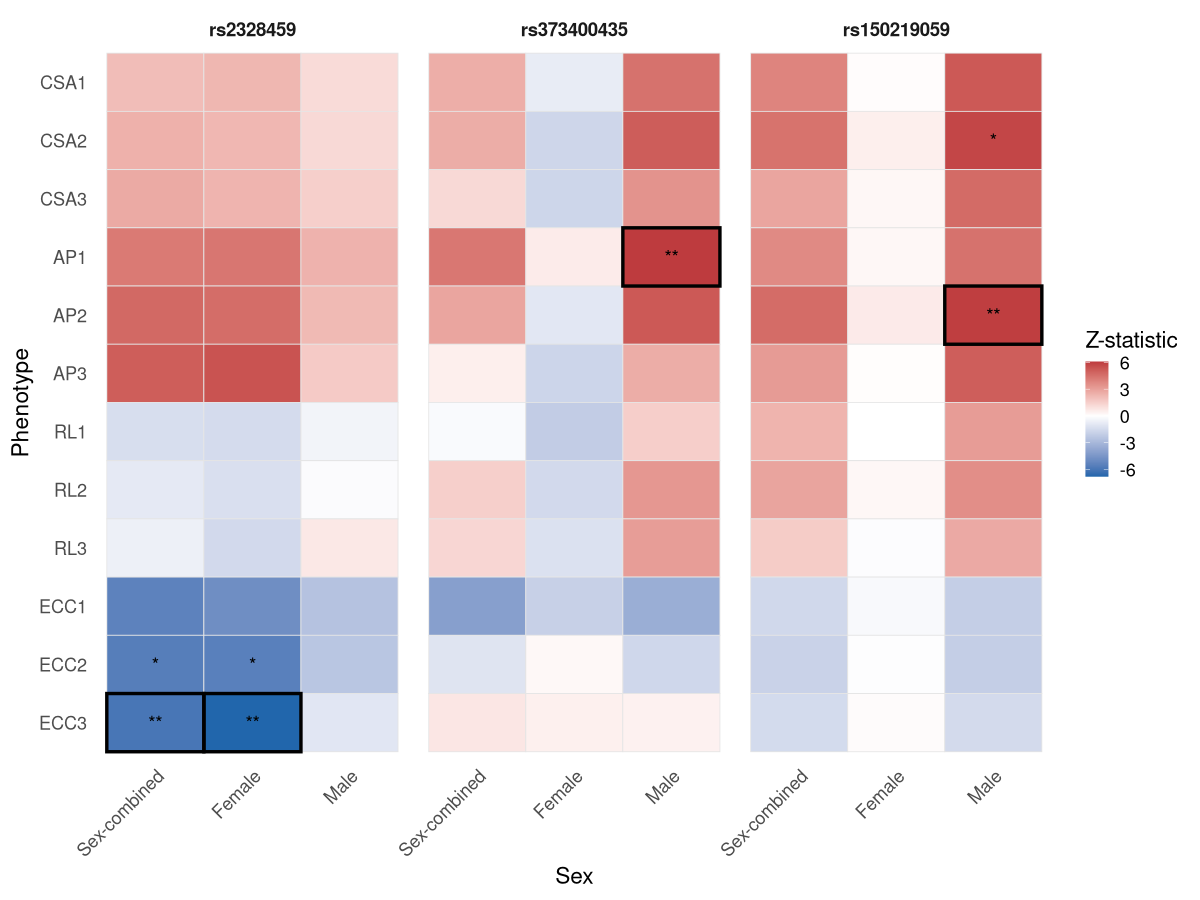

Supplementary Figure 21**.** **Heatmap showing three SNPs (rs2328459, rs373400435, rs150219059) with significant SNP-by-sex interaction effects.** Colour represents the z-statistic (beta/standard error) from sex-combined and sex-specific GWAS. A single asterisk (*) indicates P < 5 × 10⁻⁸, and a double asterisk (**) indicates P < 4.2 × 10⁻⁹.

### Section 7: Association in the non-European ancestries

To evaluate the transferability of association signals across ancestries, we assessed whether the independent significant SNPs identified in the European-ancestry GWAS of spinal cord shape metrics were also associated in non-European individuals from the UK Biobank. These analyses were conducted in three cohorts: Central/South Asian (n = 442-404), AFR (n = 222-205), and EAS (EAS; n = 207-193) participants across the C1 to C3 vertebral levels. Sample sizes slightly varied across levels due to progressively lower image quality from C1 to C3.

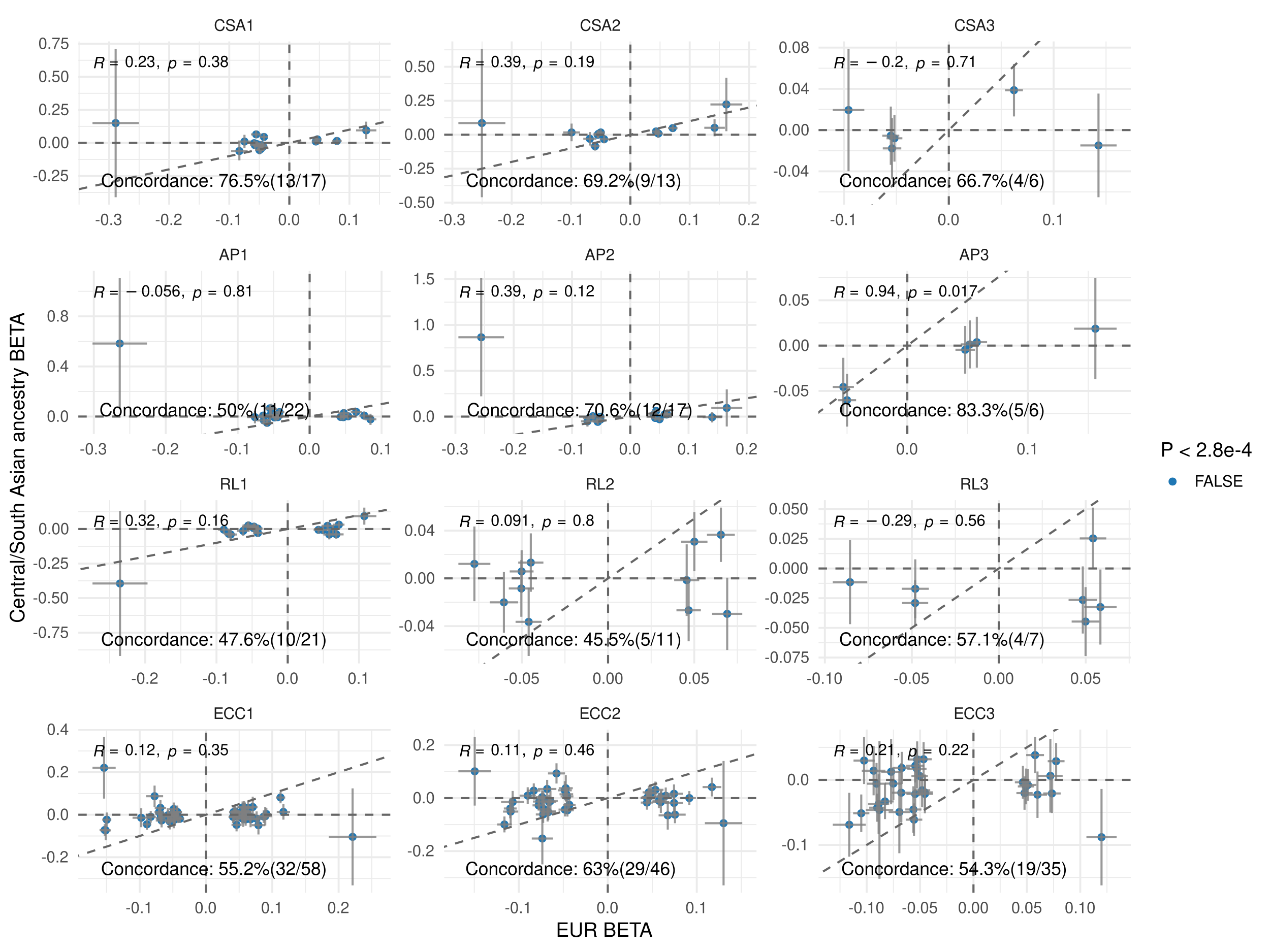

Supplementary Figure 22: Comparison of effect sizes between the European (EUR) and Central/South Asian cohorts. The plot includes 178 genome-wide significant independent SNPs (one SNP was unavailable in the CSA cohort) identified in the EUR GWAS. Spearman’s rank correlation coefficient and directional concordance are indicated. A two-sided sign test assessed whether SNP effect directions were concordant beyond chance. Each point represents the per-allele effect size, with error bars showing the 95% confidence interval. No variants reached Bonferroni-corrected significance (P < 0.00028, correcting for 179 tests).

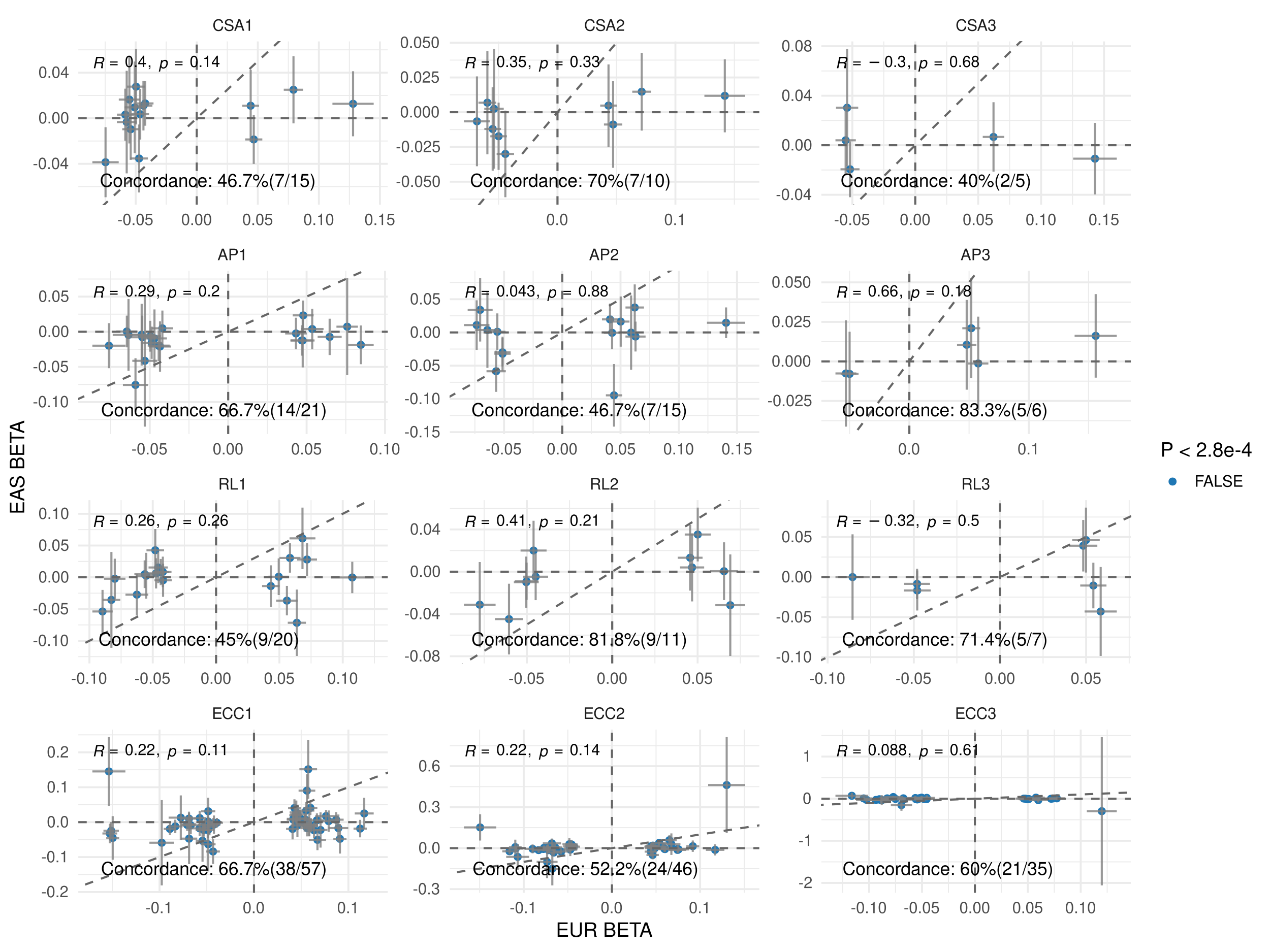

Supplementary Figure 23: Comparison of effect sizes between the European (EUR) and East Asian (EAS) cohorts. The plot includes 173 genome-wide significant independent SNPs (six SNPs were unavailable in the EAS cohort) identified in the EUR GWAS. Spearman’s rank correlation coefficient and directional concordance are indicated. A two-sided sign test assessed whether SNP effect directions were concordant beyond chance. Each point represents the per-allele effect size, with error bars showing the 95% confidence interval. No variants reached Bonferroni-corrected significance (P < 0.00028, correcting for 179 tests).

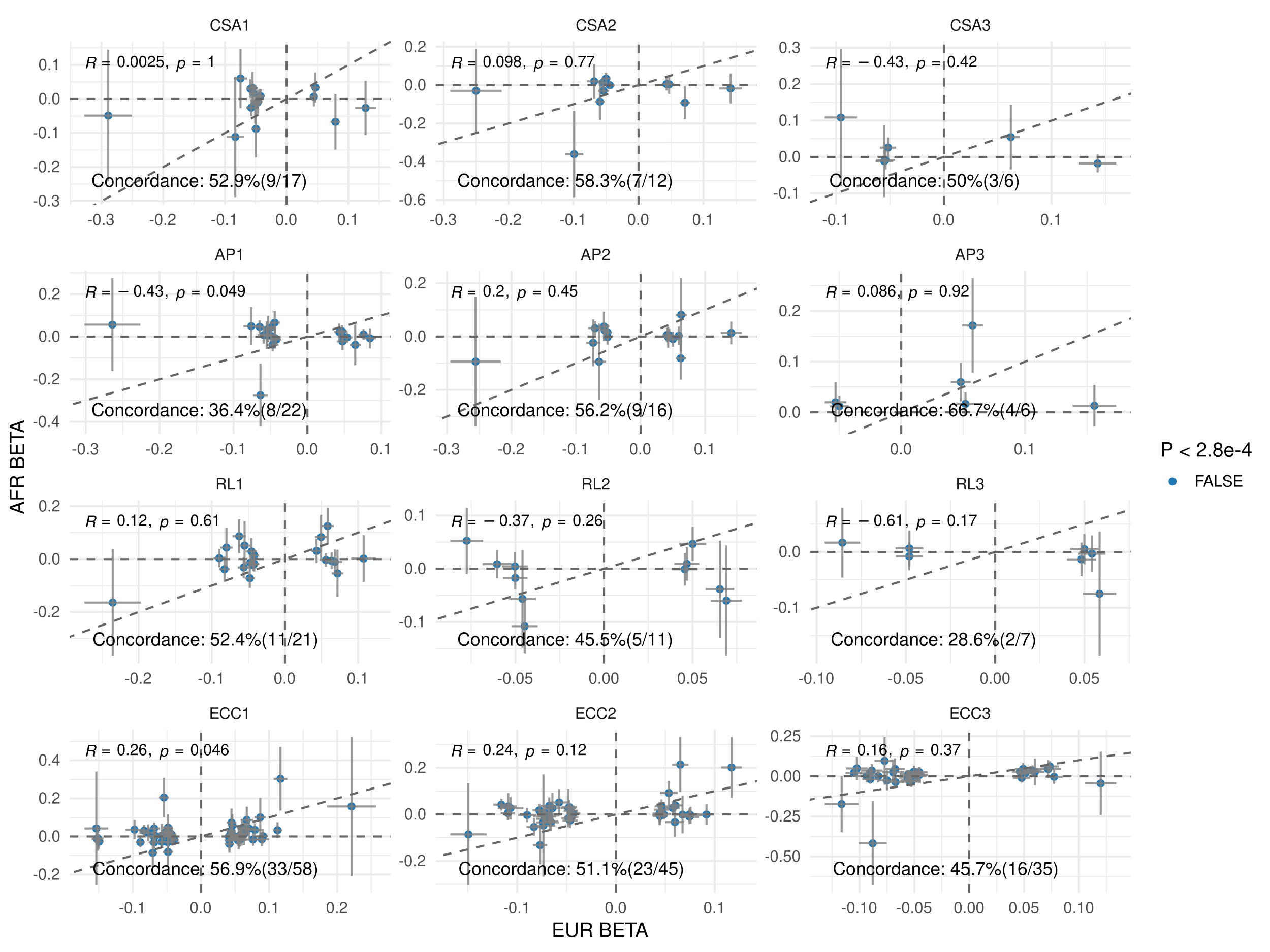

Supplementary Figure 24: Comparison of effect sizes between the European (EUR) and African (AFR) cohorts. The plot includes 176 genome-wide significant independent SNPs (three SNPs were unavailable in the AFR cohort) identified in the EUR GWAS. Spearman’s rank correlation coefficient and directional concordance are indicated. A two-sided sign test assessed whether SNP effect directions were concordant beyond chance. Each point represents the per-allele effect size, with error bars showing the 95% confidence interval. No variants reached Bonferroni-corrected significance (P < 0.00028, correcting for 179 tests).

### Section 8: Gene-based findings

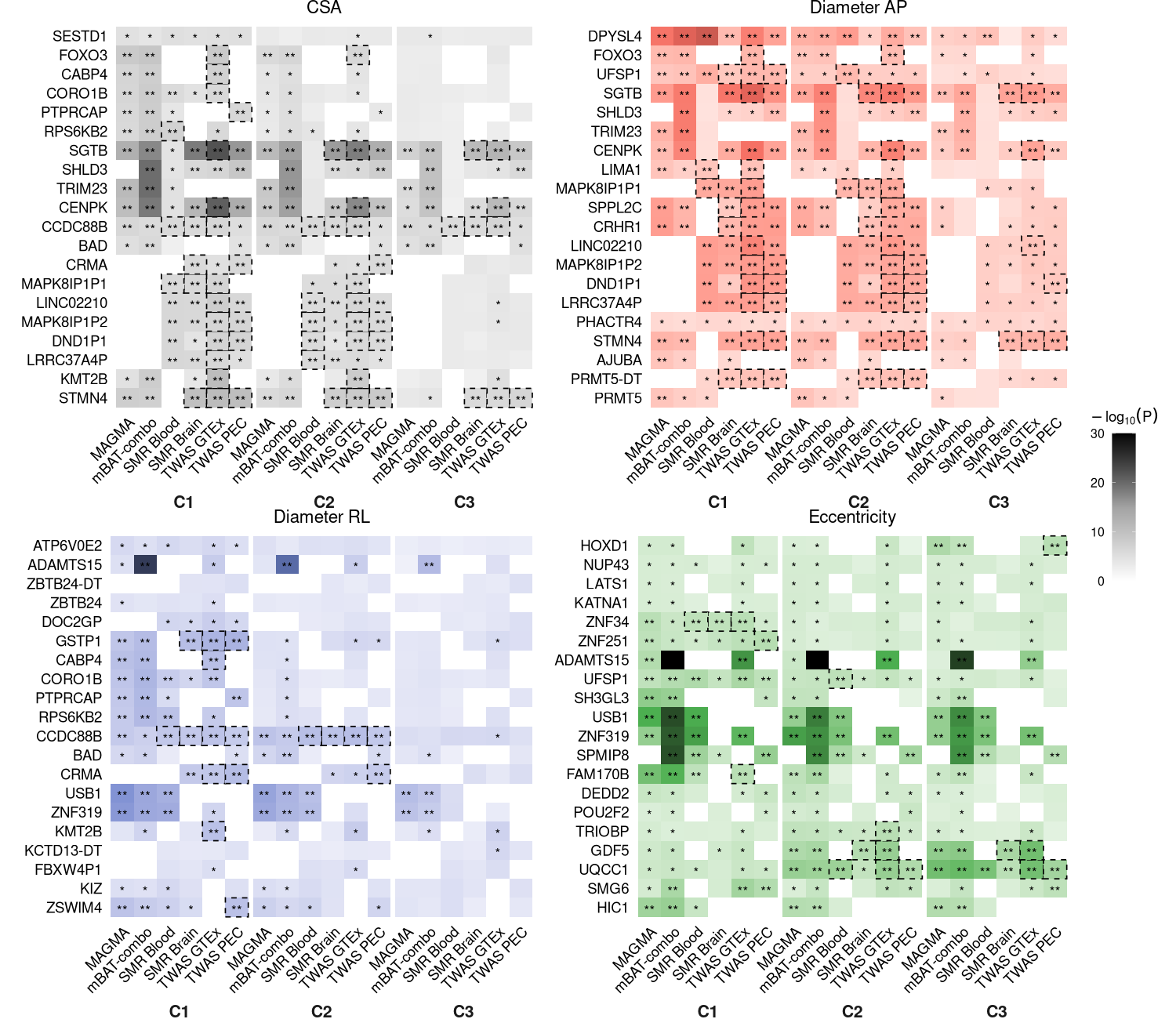

Supplementary Figure 25: **Top 20 prioritised gene-based associations for each spinal cord morphology phenotype.** For each combination of spinal cord vertebra (C1, C2, C3) and spinal cord morphology phenotype (CSA, AP diameter, RL diameter, and eccentricity), we extracted gene-level association results from six gene-based analysis methods: MAGMA, mBAT, SMR (BrainMeta and eQTLgen), and TWAS (GTEx and PsychENCODE). For each method and verbetra-morphology feature pair, we identified the gene with the minimum p-value. From these results, a set of unique gene identifiers was extracted to summarize the most significant associations across all analytic approaches of each phenotype. The genes are ordered by the number of times they were significant after Bonferroni correction across all verbetra-morphology feature pair combinations. * FDR < 0.05; ** Bonferroni P < 0.05; dashed boxes represent significant gene-based associations (Bonferroni P < 0.05) with evidence the same causal variant underlies the GWAS and eQTL association using the HEIDI test in SMR (HEIDI P > 0.05) or colocalisation in TWAS (COLOC PP.H4 > 0.8).

### Section 9: Genetic and phenotypic correlations between spinal cord phenotypes and other brain structures

To assess phenotypic correlations between spinal cord and brain structures, we selected a range of brain regions available in the UK Biobank imaging dataset. Specifically, we included brainstem subregion volumes (Freesurfer subsegmentation, Category 191), subcortical structure volumes (Freesurfer ASEG, Category 190), and cortical region volumes defined by the Brodmann Area Maps (Freesurfer BA exvivo, Category 195). These regions were chosen to capture structural variation across multiple levels of the central nervous system. Genetic correlations were calculated from summary statistics from previous GWAS studies listed in Supplementary table 25.

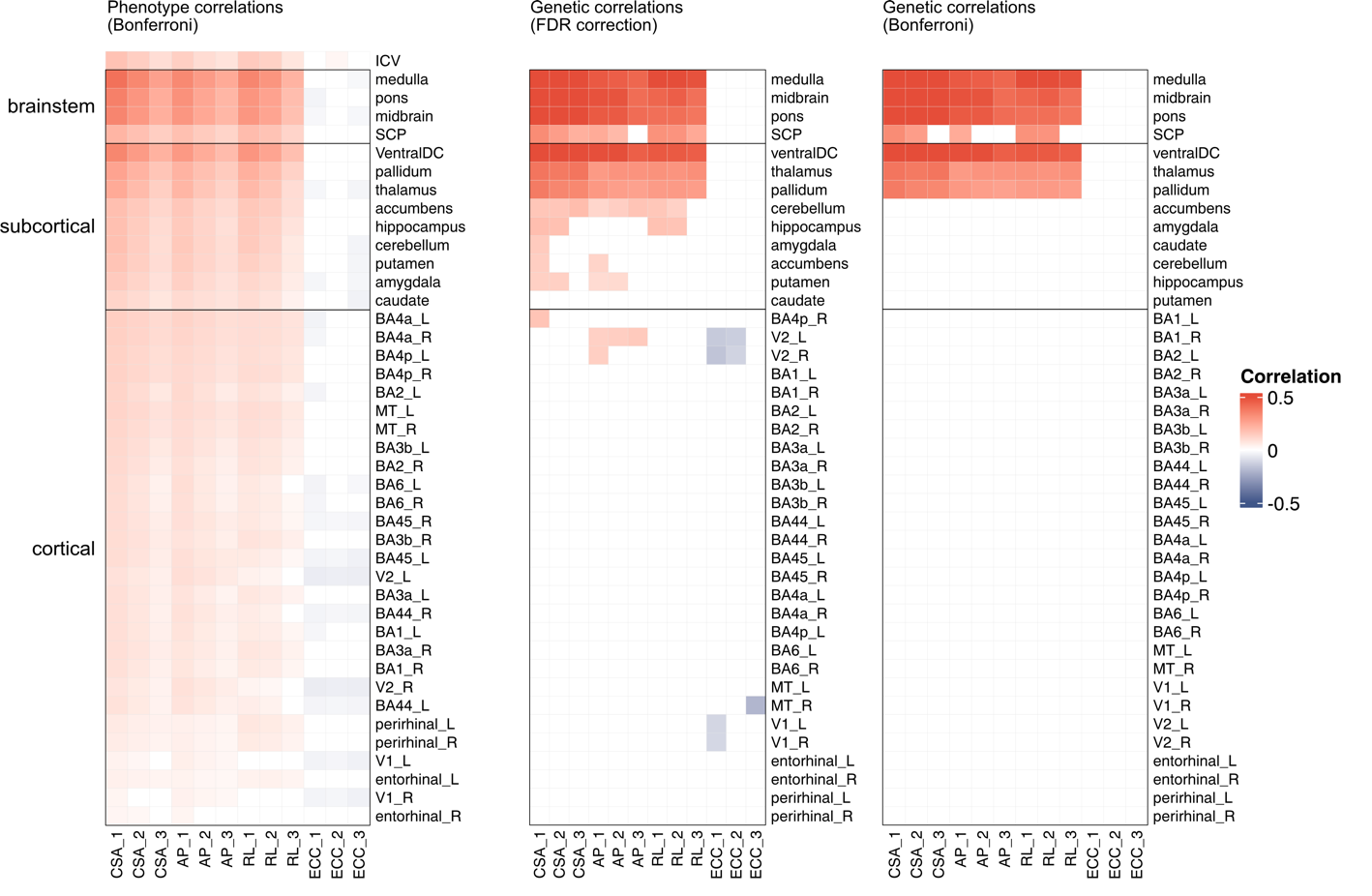

Supplementary Figure 26: Heatmaps showing correlations between spinal cord shape metrics and brain volumes. (a) Phenotypic correlations between spinal cord cross-sectional area (CSA), anterior-posterior (AP) diameter, right–left (RL) diameter, eccentricity, and brainstem, subcortical, and cortical brain volumes, including the intracranial volume (ICV). ICV was only included in the phenotypic correlations as it was used as a covariate in the GWAS. Phenotypic correlations (significant after Bonferroni correction for multiple comparisons, p < 0.05/504). (b) Genetic correlations (FDR-corrected significance threshold < 0.05). (c) Genetic correlations (significant after Bonferroni correction for multiple comparisons, p < 0.05/492).

### Section 10: Mendelian randomization

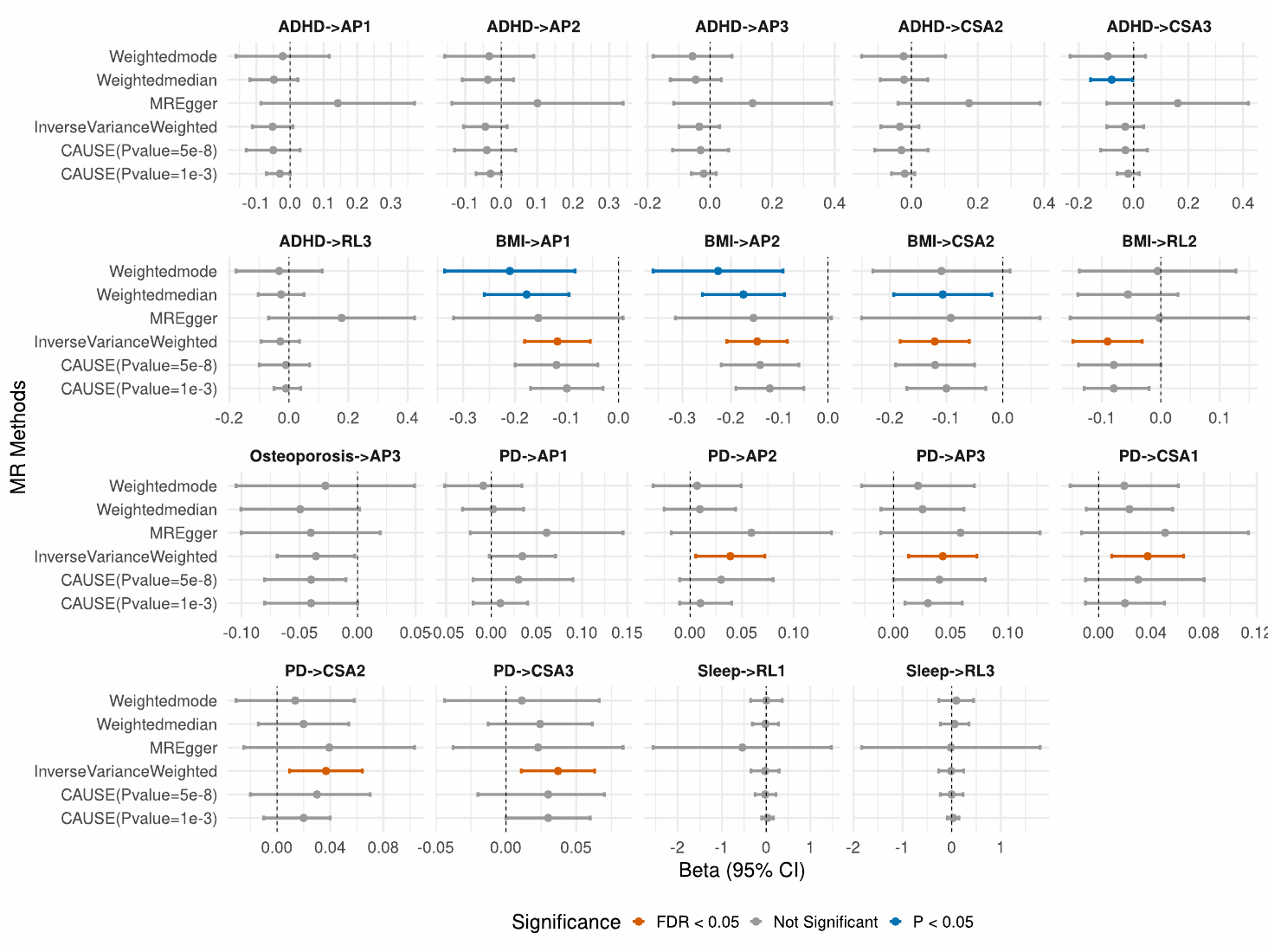

Supplementary Figure 27**: Forward Mendelian randomization (MR) analysis assessing the causal effect of spinal cord phenotypes on various traits.** The x-axis represents effect size estimates (Beta) with 95% confidence intervals (CI); the y-axis represents MR methods. Colored by statistical significance: nominal (P < 0.05) - orange, not significant - grey, or FDR < 0.05 - blue, based on the inverse-variance weighted method.

### Section 11: Spinal cord image quality control

**11.1 Image QC overview**

A key QC challenge is the inconsistent image quality across the C1 to C3 regions in the same image, primarily due to signal drop-off near the edges of the scan. To address this, we implemented a novel and rigorous QC procedure. First, T1-weighted images with poor overall quality and severe artifacts (flagged as unusable by the standard UK Biobank QC protocol^1^), were pre-excluded. Second, we applied vertebral level–specific manual QC to remove spinal cord labels with visibly poor quality. All vertebral labels were manually inspected as a conservative approach, given that these phenotypes had not been previously derived at scale. Third, we excluded additional data points showing large residuals between processing steps. This step accounts for instances where vertebral labelling was reliable, but errors still occurred during phenotype extraction from the segmentation mask.

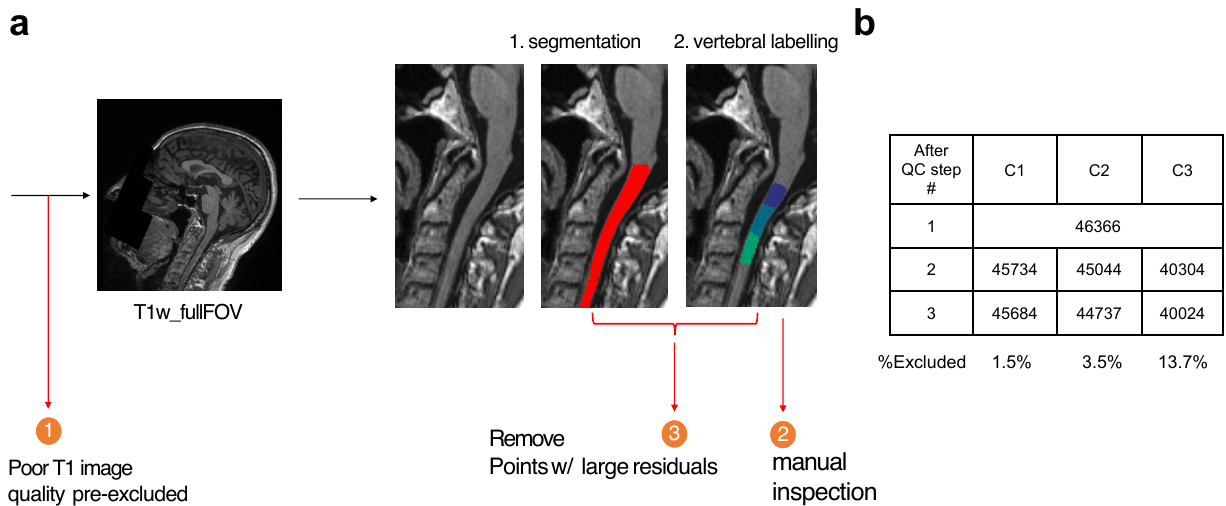

Supplementary Figure 28: summary of spinal cord image QC procedure. (a) first images with poor overall quality were pre-excluded from processing the spinal cord pipeline. Then vertebral labels were manually inspected using the manual QC procedure (figure below). Lastly, points with large residuals between processing steps were removed. (b) a summary of sample sizes after each QC step. Reproduced by kind permission of UK Biobank ©.

**11.2 Manual QC on vertebral labels**

The manual QC process followed a systematic three-step approach. First, we generated QC snapshots using SCT’s sct_qc command and screened them using a custom-built toolbox that allowed rapid identification of poor-quality or mislabelled vertebral regions. Multiple regions could be marked as QC failures. Representative examples of failed QC cases are shown in Supplementary figure 29. These failed QC cases are due to inaccurate vertebral level labelling due to poor image quality or mislabels. Second, any uncertain cases were reviewed in 3D, since the standard QC images only show mid-sagittal slices. Finally, we visualized failed cases in the context of the full phenotype distribution among every 1000 samples. For any unlabelled outliers, we conducted further manual inspection to sure these points are not produced by image artifacts.

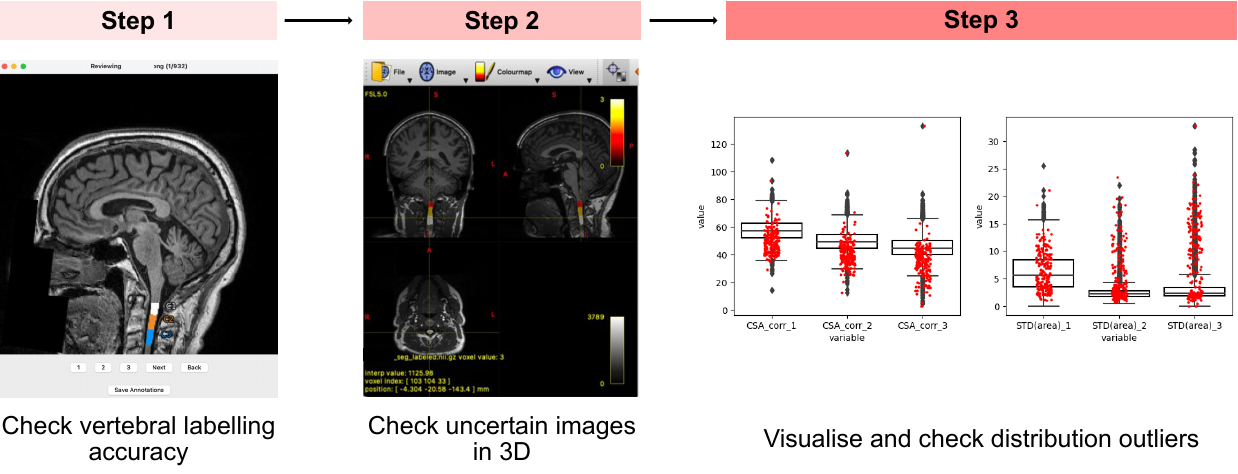

Supplementary Figure 29: The manual QC on vertebral labels follow a systematic 3-step. Step 1: visual inspection on vertebral labels generated using sct_qc. Step 2: check uncertain images in 3D. Step 3: visualise failed QC cases in distribution and check any unlabelled outliers. Reproduced by kind permission of UK Biobank ©.

­
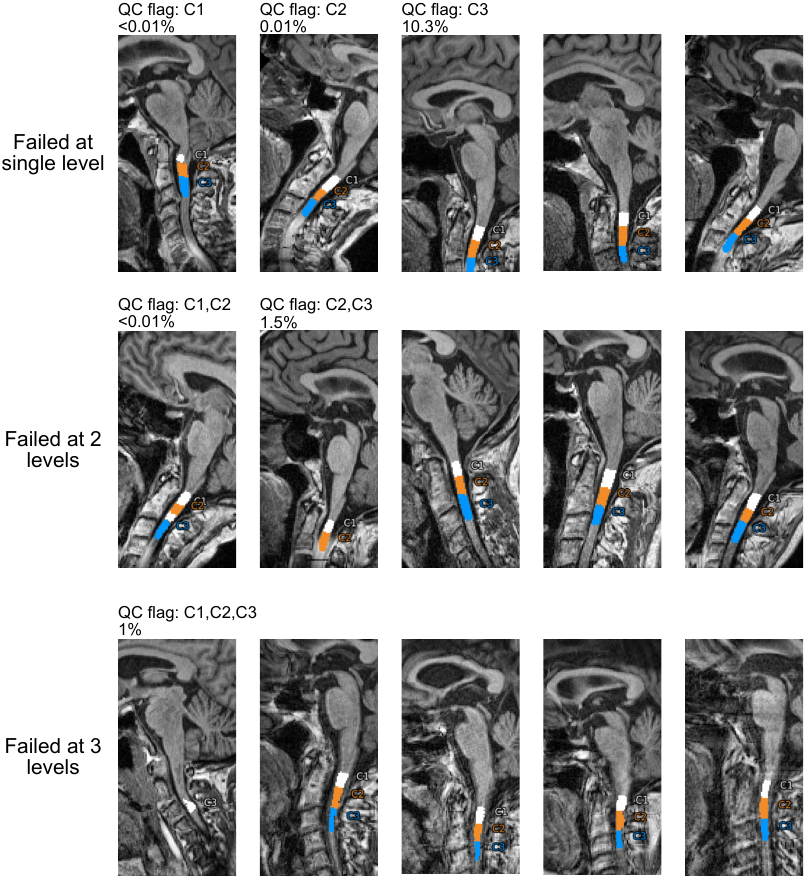
­­

Supplementary Figure 30: Examples of different QC failure scenarios, including cases with failure at a single vertebral level and those with failures across multiple levels, along with their corresponding proportions in the dataset.­ Reproduced by kind permission of UK Biobank ©.

### 11.3 Additional removal of noise between processing steps

To remove any points with inconsistent values between processing steps, we compared shape metrics derived from the segmentation mask and those from the vertebral label image. Residuals between the two were calculated, and any point exceeding 5 standard deviations was excluded. These discrepancies were typically due to inaccuracies in the segmentation mask, which were corrected during vertebral labelling, as the model was trained to preserve the tubular structure of the spinal cord. This served as the final QC step for fine cleaning. Only a small percentage of noisy points were removed, with the majority showing high agreement. Although the metrics derived from the vertebral label image more closely resemble the results of manual QC, we did not use them for downstream analysis because they are not based on true axial slices and do not align with the image plane.

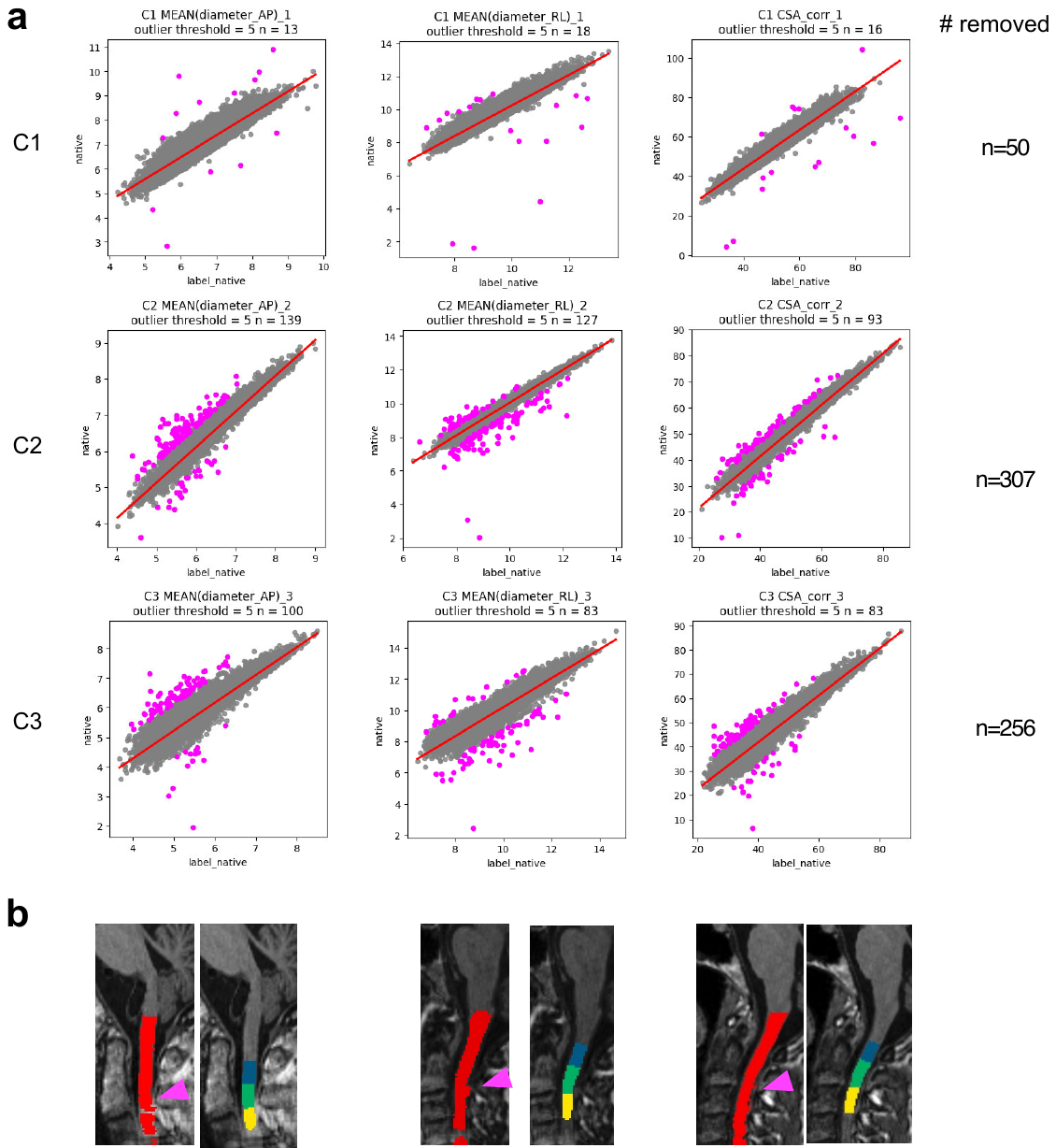

Supplementary Figure 31**:** (a) Scatterplot showing residuals between shape metrics derived from segmentation masks and vertebral labels. Points exceeding 5 standard deviations were excluded (pink). (b) Example images of excluded points, with arrows highlighting segmentation errors (left) corrected during vertebral labelling (right). Reproduced by kind permission of UK Biobank ©.

**References**

1.	Alfaro-Almagro, F. *et al.* Image processing and Quality Control for the first 10,000 brain imaging datasets from UK Biobank. *Neuroimage* **166**, 400–424 (2018).

2.	De Leener, B. *et al.* PAM50: Unbiased multimodal template of the brainstem and spinal cord aligned with the ICBM152 space. *Neuroimage* **165**, 170–179 (2018).
